# Quarantine and testing strategies to ameliorate transmission due to travel during the COVID-19 pandemic: a modelling study

**DOI:** 10.1101/2021.04.25.21256082

**Authors:** Chad R. Wells, Abhishek Pandey, Meagan C. Fitzpatrick, William S. Crystal, Burton H. Singer, Seyed M. Moghadas, Alison P. Galvani, Jeffrey P. Townsend

## Abstract

**Background:** Numerous countries imposed strict travel restrictions, contributing to the large socioeconomic burden during the COVID-19 pandemic. The long quarantines that apply to contacts of cases may be excessive for travel policy.

**Methods:** We developed an approach to evaluate imminent countrywide COVID-19 infections after 0–14-day quarantine and testing. We identified the minimum travel quarantine duration such that the infection rate within the destination country did not increase compared to a travel ban, defining this minimum quarantine as “sufficient.”

**Findings:** We present a generalised analytical framework and a specific case study of the epidemic situation on November 21, 2021, for application to 26 European countries. For most origin-destination country pairs, a three-day or shorter quarantine with RT-PCR or antigen testing on exit suffices. Adaptation to the European Union traffic-light risk stratification provided a simplified policy tool. Our analytical approach provides guidance for travel policy during all phases of pandemic diseases.

**Interpretation:** For nearly half of origin-destination country pairs analysed, travel can be permitted in the absence of quarantine and testing. For the majority of pairs requiring controls, a short quarantine with testing could be as effective as a complete travel ban. The estimated travel quarantine durations are substantially shorter than those specified for traced contacts.

**Funding:** EasyJet (JPT and APG), the Elihu endowment (JPT), the Burnett and Stender families’ endowment (APG), the Notsew Orm Sands Foundation (JPT and APG), the National Institutes of Health (MCF), Canadian Institutes of Health Research (SMM) and Natural Sciences and Engineering Research Council of Canada EIDM-MfPH (SMM).

**Research in context:** *Evidence before this study:* Evidence from early in the pandemic indicates that border closures at the epicentre slowed global dissemination of COVID-19. As community transmission became established in many nations, studies have suggested that the benefit of strict border closures in mitigating the transmission of disease from travellers diminished. Research for community settings has shown that testing later during quarantine, rather than upon entry into quarantine, can substantially shorten the duration of quarantine needed to reduce post-quarantine transmission. In particular for international air travellers, a 14-day quarantine can effectively be shortened to five or seven days. The number of infectious COVID-19 cases that escape from these quarantines depends on the prevalence of disease in the country the traveller originated as well as the travel volume into the country.

*Added value of this study:* We developed a framework to identify quarantine and testing strategies that enable travel from specific origins without increasing their infection rates per capita within destinations. No prior study has evaluated the appropriate duration of quarantine necessary to prevent any rise in infection rates per capita in the destination countries as a result of travel. By accounting for prevalence, daily incidence, vaccine coverage, immunity, age demographics, and travel flow between countries, we quantified the contribution of travel towards within-country the imminent infections in the destination country under different quarantine and testing strategies. For travel between 26 European countries, our results for the pandemic situation observed on November 21, 2021 demonstrate that there are often less burdensome quarantine and testing strategies that can serve as effective alternatives to strict border closure. Specifically, these estimated sufficient quarantine durations are especially dependent on COVID-19 prevalence and immunity within the two countries. We also found that asymmetry in the travel flow, just not the volume of travel flow, can also influence the estimated sufficient quarantine durations. Using data on variants of concern, including Omicron, we found that the adequacy of a border control strategy to limit variant spread depends strongly on the geographical distribution of the variant. While our results pertain to European countries, we also provide an interactive spreadsheet that can be used to determine appropriate quarantine durations between any two countries. Moreover, our framework can also be applied at any spatial or population scale within which movement restrictions could feasibly be implemented.

*Implications of all available evidence:* Travel quarantine and testing strategies can effectively mitigate importation and onward transmission within a country. Identifying sufficient strategies can allow countries to permit travel to and from other countries, without risking a short-term increase in infection rates. As long as the community transmission is occurring, the long-term epidemic trend within the destination country is more apt to be determined by other disease control measures, e.g., contact tracing, vaccination, and non-pharmaceutical interventions. Together, travel quarantine and other related control measures can mitigate the risk of transmission between countries, limiting the threat of variants of concern.

## Introduction

Quarantine of travellers to prevent the importation of disease has been a cornerstone of efforts to prevent infectious disease since at least the fourteenth century. ^1^ Ongoing efforts to limit the importation and global dissemination of COVID-19 cases have included the imposition of strict national border control measures, typically mandating an extensive quarantine.^2^ These restrictions were aimed to slow or mitigate the spread of SARS-CoV-2 to heretofore unafflicted nations.^3^ However, border closures at the epicentre only minimally delayed global spread. ^4^ Moreover, border restrictions and extended quarantines have had diminishing effects as the pandemic has progressed. ^5–7^ While regions that implemented international travel controls early experienced a delay in the time of the initial peak—compared with regions that waited ^8^—imported cases have had little impact on the epidemic trajectory where community transmission was well established. ^9^ Furthermore, these interventions carry a large socioeconomic burden. A policy that enables safe travel between countries and shortens travel quarantine without increasing the country-specific infection rate would aid in the rejuvenation of the international economy.

Quarantine duration has traditionally been set with the objective of ensuring prevention of post-quarantine transmission with or without testing. ^10^ Such a goal is reasonable for those entering a country experiencing zero or near-zero prevalence of disease, ^11^ or for a traced contact with a high chance of being infected. For example, the World Health Organization recommends a 14-day quarantine of exposed individuals to mitigate onward transmission from COVID-19 cases who have extended incubation periods. ^12,13^ However, travellers are typically far less likely to have been infected than traced contacts; if disease prevalence in their origin country is not substantially elevated in comparison to their destination, then their risk of transmitting is presumably equivalent to residents of the destination country who are following the same country-specific public health precautions. Because international travel is generally considered a desirable activity for diverse socioeconomic reasons, it would be reasonable for border controls in these global pandemic scenarios to have the objective not of preventing any transmission of pandemic disease by travellers—which, after all, is typically exchangeable in impact with transmission by residents—but instead ensuring that travel does not cause a net increase in disease compared to border closure or complete ban of travel. Because sufficient quarantine and testing at the border can decrease within-country post-quarantine transmission to arbitrarily low levels, ^10,14–16^ and infected individuals departing the country diminish within-country transmission, such a balance should be determinable.

Questions regarding the efficacy of travel quarantine and testing have recently been brought to the fore by the emergence of variants of concern of SARS CoV-2 (VOCs). Some VOCs are more transmissible than previous strains, ^17^ and there is uncertainty regarding the extent of protection provided by natural and vaccine acquired immunity against each VOC. ^18–20^ VOC emergence has spurred the renewal of border closures, long quarantines, and extensive COVID-19 testing to prevent VOCs from establishing in unaffected countries. ^5^ Impeding the establishment of VOCs with an appropriate travel quarantine strategy could provide time for vaccine coverage to be scaled up, limiting overall transmission. ^21^ However, it is unclear what minimal travel quarantine and testing strategies are sufficient to reduce the seeding of new VOC cases. Therefore, knowledge of the potential suppressive effects of travel quarantine and testing on prevalence of known VOCs is important to limiting the resurgence of the disease, and should be considered in the specification of country-specific and individualised travel quarantine strategies.

Here we integrate analytical approaches for the calculation of post-quarantine transmission ^10^ with a model of country-specific imminent infections (i.e., the infections that will be generated from currently infected individuals) to develop a generalised analytical framework to identify travel quarantine and testing strategies such that infections will not increase in the destination country when compared to a strategy of complete border closure. To quantify the minimum sufficient duration of travel quarantine among each origin-destination pair of 26 countries in Europe, we incorporate the travel flow between countries and country-specific age structure, while accounting for natural immunity, vaccine coverage and disease burden observed on November 21, 2021 across Europe. We also generalised the analysis to provide quarantine and testing strategies based on European Union (EU) traffic-light stratification of COVID-19 risk instead of country-specific strategies.

## Methods

### Sufficient travel quarantine

To identify travel quarantine and testing strategies that do not elevate a country’s case burden when travel is resumed with another country, we evaluated the extent of transmission in a country when travel is allowed, compared to the transmission occurring under a travel ban (**Fig. 1**). Our measure of transmission was the daily new within-country imminent infections (generated over infected individuals’ remaining disease time course). When considering travel between countries, we assume long-term stays such that prevalence in travellers rises to the prevalence in the country visited by the time of return. The assumption of long-term stays instead of short-term stays simplifies the calculations of imminent infections when allowing travel between two countries, and provides a conservative estimate of the minimum sufficient quarantine duration (**Supplementary material: Sufficient travel quarantine**). We do not specifically model inflight transmission in our calculations, as the risk of inflight transmission with appropriate precautions is low. ^2,22^

**Figure 1.**
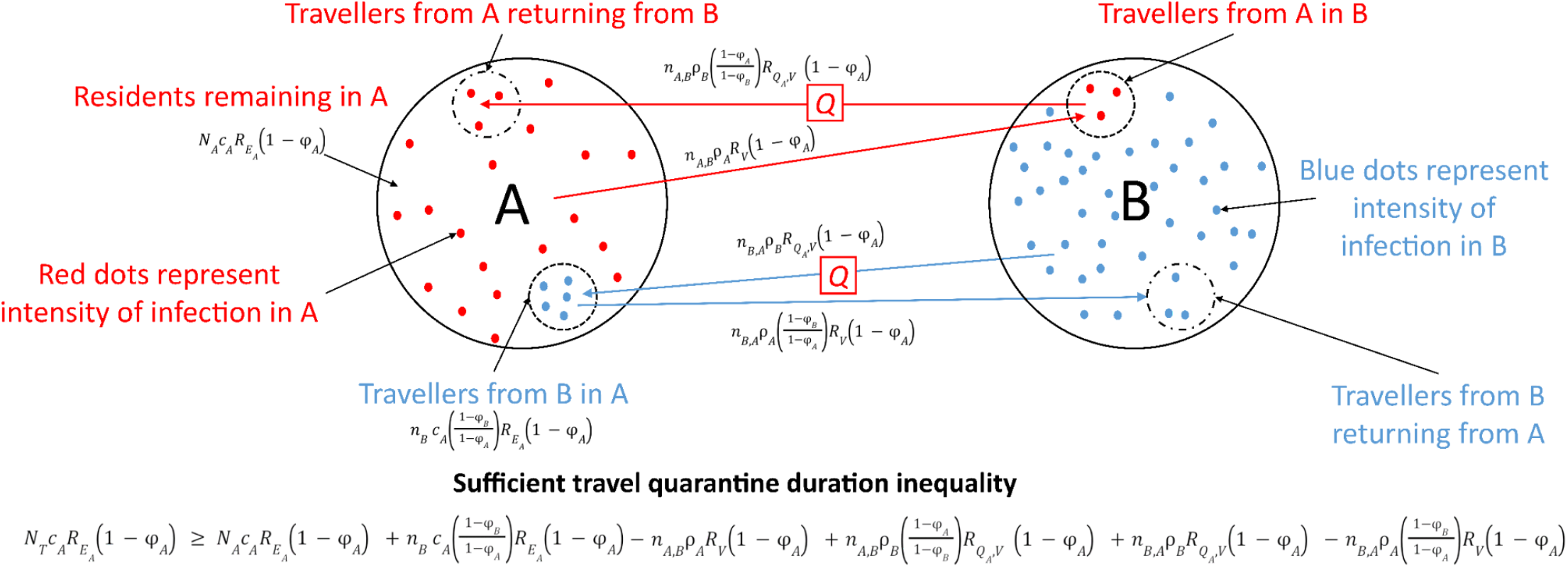
Model schematic diagram for daily new within-country imminent infections and travel quarantine. With travel between country A (red) and country B (blue), the travel quarantine specified by country A is dependent on the intensity of infection in both country A and B, the number of residents leaving country A for country B, the number of travellers entering country A from country B, the number of residents returning to country A, and the number of travellers from country B returning to country B. In the most general instantiation of our model, the minimum duration of travel quarantine is dependent on the number of daily travellers from country A to country B, *n*_*AB*_, the number of daily travellers from country B to country A, *n*_*BA*_, the country-specific prevalence of non-isolated infections *ρ*, country-specific immunity *φ*, the number of travellers from country B abroad in country A, *n*_*B*_, the number of citizens of A who are non-travellers and not quarantined, *N*_*A*_, daily incidence per capita, *c*. The amount of remaining transmission (*R*_*E*_, *R*_*V*_, *R*_*Q,V*_) is calculable based on virus- or variant-specific properties and the temporal infectivity and test sensitivity ^10^.

### No travel

Daily new within-country imminent infection in country A (destination country) with no travel between country A and country B is 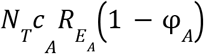, where *N*_*T*_ is the total population of country A, *c*_*A*_ is the average daily incidence per capita (over the last two weeks) within country 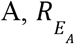 is the effective reproduction number accounting for self-isolation upon symptom onset, and *φ*_*A*_ is the percent of country A that is immune. The term *φ*_*A*_ can be decomposed into the fraction who are currently infected plus the fraction that have either recovered from infection and remain immune or have been vaccinated and remain immune.

### Travel between countries A and B

Country-specific imminent infection in country A with long-term travel is composed of non-travellers and travellers. The contribution to daily new within-country imminent infection of non-travellers is 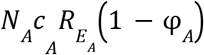, where *N*_*A*_ is the number of non-traveling and non-quarantined country-A residents. At steady-state with respect to travel, the contribution to daily new within-country imminent infection of returning travellers is 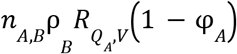, where *n*_*A,B*_ is the daily number of country-A residents returning from a long-term visit to country B, ρ_*B*_ is the prevalence of non-isolated infections in country B, and 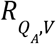 is the average ‘left-over’ imminent infection in country A of an infected visitor who underwent the quarantine-and-testing regimen *Q*_*A*_. Similarly, daily new within-country imminent infections are increased by arrival of infected travellers from country B, whose contribution is 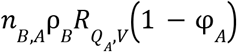, where *n*_*B,A*_ is the daily number of long-term visitors from country B to country A. Country-specific imminent infection in country A is also increased by travellers from country B who become infected in country A; their contribution towards daily new within-country imminent infection is 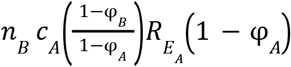, where *n*_*B*_ is number of residents of country B abroad in country A, and 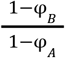 is the infection risk ratio for individuals who are not immune to infection at frequency 1 − *φ*_*B*_ while in country A with its frequency of susceptibility 1 − *φ*_*A*_.

Simultaneously, the departure of infected residents decreases daily new within-country imminent infection by 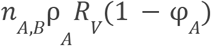, where *R*_*V*_ is the average ‘left-over’ imminent infection of an infected resident of country A leaving at an unknown time in their infection period. The departure of infected individuals from country B also decreases daily new within-country imminent infection by 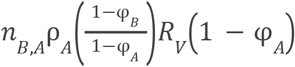. Therefore, permitting the population in country A to perform long-term travel to and return from country B, as well as permitting long-term travel from country B to country A, is no worse than border closure in terms of daily new within-country imminent infection when

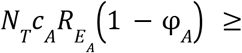

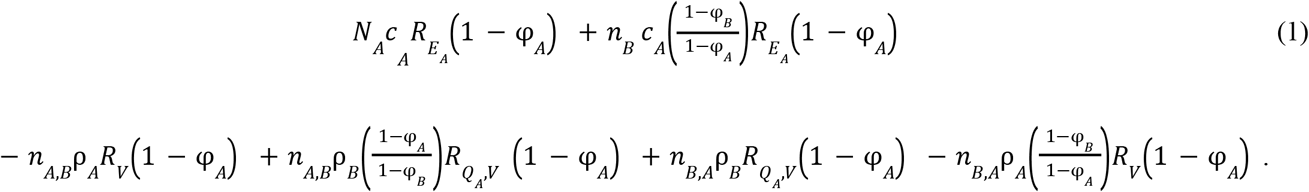

For a specified testing strategy, the minimum quarantine duration that satisfies this condition is defined as a sufficient travel quarantine.

### Effective reproduction numbers

To determine the effective reproduction numbers (**Supplementary material: Effective reproduction number**), we first constructed an average temporal infectivity profile for an infected individual ^23,24^ using a reproduction number of 3 and an incubation period of 5·72 days (**Supplementary material: Infectivity curve** and **Fig. S9**). ^10,25–27^ Accounting for the proportion of infections that remain asymptomatic after the incubation period, ^28^ we calculate an effective reproduction number for non-travellers using the average infectivity profile while assuming individuals self-isolate upon symptom onset. For travellers, we also discount the periods in the average temporal infectivity profile when they can not transmit (e.g. isolation, quarantine or travel abroad).

### Travel quarantine strategies

For each origin-destination country pair, we evaluated a travel quarantine strategy of 0–14 days quarantine duration with a RT-PCR test conducted on exit. If quarantine durations up to 14 days were not sufficient, then we indicated a travel ban. A delay of 24 h was assumed in obtaining results for an RT-PCR test (e.g. an exit RT-PCR test is conducted 24 h before exit from quarantine); we specified no delay in obtaining antigen test results. We considered three alternative quarantine strategies: i) no testing, ii) antigen testing on exit from quarantine, and iii) antigen testing on entry to and exit from quarantine.

### Diagnostic sensitivity

To determine the temporal diagnostic sensitivity of the RT-PCR assay over the course of disease, we fit a time-dependent function (represented by a log-Normal probability density function) to the results from serial testing, ^29^ with the sensitivity of the rapid antigen test also dependent on the percent positive agreement of the BD Veritor rapid antigen test (**Supplementary material: Temporal diagnostic sensitivity** and **Fig. S8**).^30^

### Country-paired analysis

We applied our modelling framework to 26 European countries, using country-specific vaccine coverage, ^31–34^ natural immunity, estimated daily incidence, ^35^ prevalence, age demographics, ^36^ and 2019 country pairwise travel flow (**Table S1**). To calculate the level of vaccine-acquired immunity, we used age- and dose-specific effectiveness in the reduction of documented infection, ^37^ specifying age- and country-specific vaccination coverage. ^38^ Natural immunity is inferred from the estimated number of cumulative infections in the country,^39^ which accounts for under-reporting of cases. We specify prevalence to be infections that are active and not isolated based on estimated daily incidence. To quantify country’s pairwise travel flow data, annual arrivals for all forms of paid accommodation was utilised for the majority of countries (**Table S1**). Using this annual data, we calculated the average number of daily travellers between the countries. The use of arrivals for all forms of paid accommodation reflects the multitude of travel routes (e.g., automobile, train, or aeroplane) into a country and does not limit it to just airline travel.

### Generalised approach based on European Union traffic-light system

We also conducted a generalised implementation of our modelling framework aligned to the EU traffic-light system to contrast the results with our country-specific analysis. Unlike the country-specific analysis that is highly parameterized, we applied our approach for the EU traffic-light model based on differences in incidence and prevalence alone, otherwise specifying the average traits of European countries (**Supplementary material: Generalised approach using EU traffic-light system**). The EU traffic-light system for COVID-19 travel restrictions stratifies epidemic status of a country based on caseload over the preceding two weeks: i) < 25 cases (Green); ii) 25–150 cases (Amber); iii) 150–500 cases (Red); and iv) > 500 cases (Dark Red), per 100,000 residents. We considered four categories of countries stratified by their risk status: 25/100,000 (Green status), 150/100,000 (Amber status), 500/100,000 (Red status), and 1,000/100,000 (Dark Red). Assuming symmetric travel flow for the resulting 16 combinations, we calculated the minimum sufficient travel quarantine for each origin-destination pair.

### Variants of concern

In our base case scenario, we evaluated the goal of eliminating the impact of travel on all within-country imminent infections without distinguishing among genetic variants or their characteristics. To quantify quarantine durations that can prevent variants of concern from seeding or increasing the extent of its transmission in a destination country, we evaluated travel quarantine strategies based on i) daily new within-country imminent infections emanating from a particular VOC and ii) daily new within-country imminent infections from multiple variants. We determined the minimum sufficient quarantine when considering multiple VOC by calculating the minimum duration of travel quarantine that satisfies the daily new within-country imminent infection inequality for all variants considered (**Fig. 1**). We considered two known and prevalent VOCs, Delta G/478K.V1 and Omicron B.1.1.529+BA. ^40^ We characterised the VOCs with increased transmission relative to the general transmission, 100% cross-immunity, and no reduction in vaccine effectiveness. Using the estimate that the Delta G/478K.V1 VOC is 60% more transmissible within households than the Alpha 202012/01 GRY VOC, ^41,42^ we assumed that the Delta G/478K.V1 VOC is 191·2% more transmissible relative to general transmission. Based on early estimates that the odds of household transmission of Omicron B.1.1.529+BA relative to Delta G/478K.V1 is 3.2,^43^ we assumed that Omicron B.1.1.529+BA is 413·1% more transmissible relative to general transmission (i.e., 76·2 % more transmissible than Delta).

### Sensitivity and scenario analysis

By assessing the change in sufficient quarantine durations as a result of the perturbations in each parameter, we identified how the parameters influence our estimates. To conduct this one-way sensitivity analysis, we calculated sufficient quarantines after iteratively altering the value of each parameter by one standard deviation towards the median from the 26 countries, while retaining all other parameters fixed (**Supplementary material: Sensitivity analysis**).

To evaluate the impact of the different assumptions on the duration of the minimum sufficient quarantine, we conducted scenario analyses that computed the sufficient quarantine durations associated with 1) alternative durations of the incubation period, 2) reduced adherence to quarantine or self-isolation upon symptom onset, 3) serial dates of analysis, and 4) an alternative measure of travel flow (airline passenger data). For different values of the incubation period, we examined a longer reported incubation period of 8·29 days ^44^ and an extended incubation period of 11·66 days (based on the 95th percentile of the reported distribution). ^45^ When varying the adherence to self-isolation, we only vary it for individuals who are not in quarantine, as isolation is mandatory in quarantine. Lack of adherence to quarantine was parameterized as complete; transmission for those not adhering to quarantine was specified as equivalent to the scenario of a zero-day quarantine with no test. To perform a scenario analysis examining the differences in outcome across serial dates of analysis, we quantified the minimum duration of quarantine at weekly intervals between October 3, 2021 and November 21, 2021 and then compared these durations to those estimated for October 3, 2021. To calculate imminent infections based on an alternative source of travel flow data, annual air passenger transport between countries was used. ^46^

## Results

### Country-specific quarantines

Evaluation using our full model with all parameters determined by the epidemic situation observed on November 21, 2021 yielded sufficient quarantine durations—when an RT-PCR test is conducted on exit from quarantine—to enable travel without increasing within-country transmission (**Fig. 2**). From the estimated country-specific travel quarantine strategies that are individualised for origin and destination pairs, we found that travel can be allowed among most pairings without increasing the within-country imminent infections in the destination country (**Fig 2, Fig S1**). Countries with lower prevalences of disease tended to require more stringent regimens of quarantine and testing (**Fig. S2**). For example, as of November 21, Spain was classified in Red EU travel status and exhibited the lowest case burden per capita among the 26 European countries studied. We found that quarantine durations ranging between 0–4 days would result in fewer imminent infections in Spain than closed borders for most of the origin countries, with a travel ban from the UK (**Fig. 2A**). Malta was also in Red status with a minimum sufficient quarantine duration between 2–10 days, with a travel ban from Germany and Poland (**Fig. 2B, Fig. S1**). Cyprus, Portugal, the UK, Greece, and Austria—each with Dark Red EU travel status—exhibited minimum sufficient quarantine durations ranging from 0–2 days to 0–12 days with the implementation of travel bans (**Fig. 2C–G**). A Dark Red EU travel status country such as Hungary—with high prevalence—can require no quarantine (**Fig. 2H**). In general, we found that the recommended duration of travel quarantine increases with the ratio of the prevalence in the origin country to the destination country (**Fig. 2I**).

**Figure 2.**
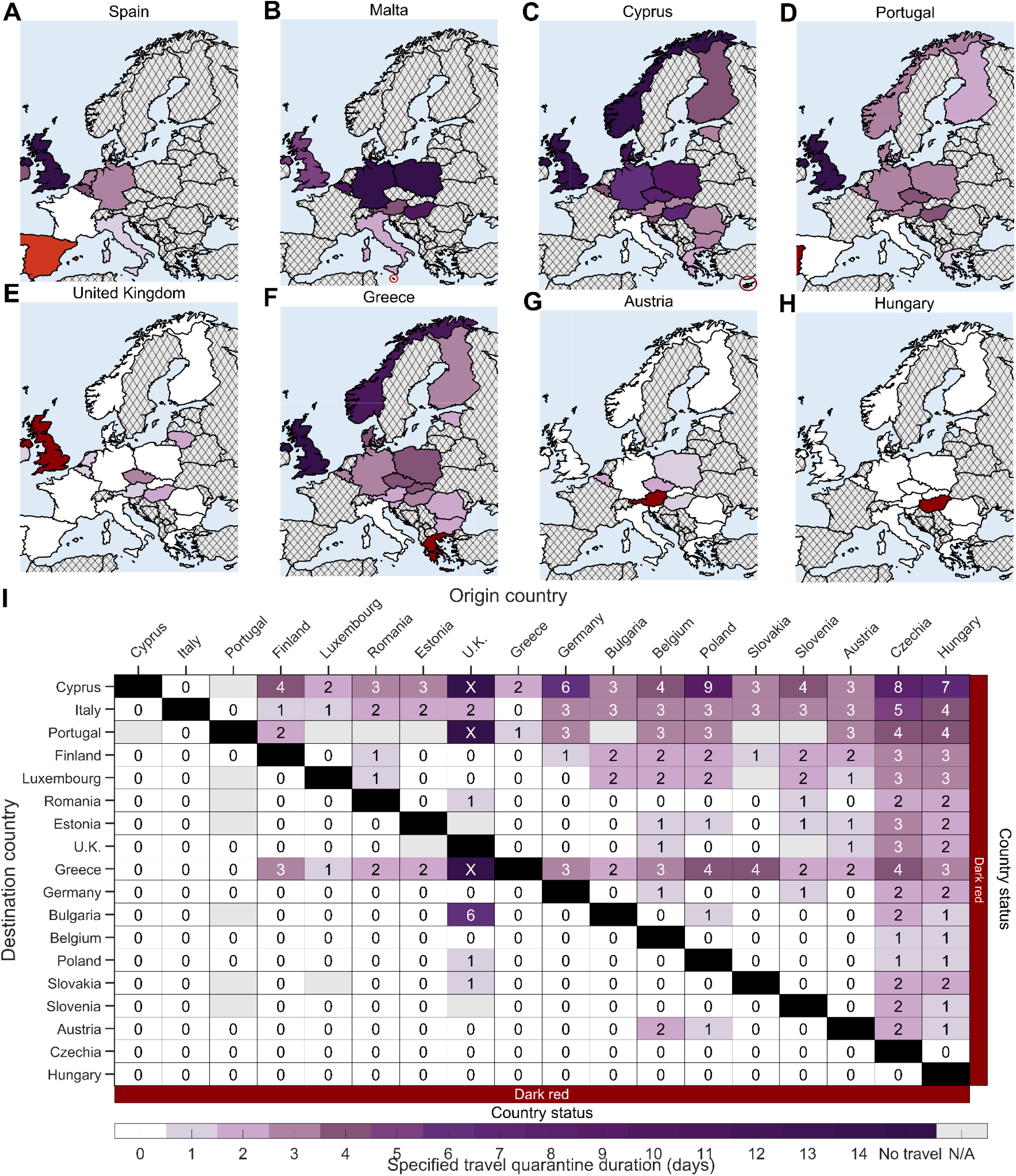
The estimated minimum duration of travel quarantine for specified origin-destination country pairs that reduces within-country imminent infections to be equivalent to border closure for the pandemic as of November 21. Specifying age-dependent vaccine effectiveness and proportion of asymptomatic infections, as well as country-specific demographics, incidence, prevalence of non-isolated infections, vaccine coverage, natural immunity and travel flow, we determine the minimum sufficient duration of travel quarantine with an RT-PCR test on exit from quarantine (colour gradient) that should be stated by the destination country for individuals arriving from the origin country based on data for November 21, 2021. The countries are ranked based on their estimated incidence per 100,000 over the last two weeks (November 8 to November 21) and stratified based on the European Union country classification system: Green, < 25 cases per 100,000; Amber, 25–150 cases per 100,000; Red, 150–500 cases per 100,000; and Dark Red, > 500 cases per 100,000. We consider travel quarantine durations of zero-days (white) to no travel (dark purple with “X”, i.e., sufficient travel quarantine would exceed 14 days) for (**A**) Spain, (**B**) Malta, (**C**) Cyprus, (**D**) Portugal, (**E**) the United Kingdom, (**F**) Greece, (**G**) Austria, (**H**) Hungary, and (**I**) 18 of 26 countries analysed (*cf*. **Fig. S1**). Within-country travel quarantine is not evaluated in the analysis (black). Travel flow data was not available for all origin-destination country pairs (grey). For quarantine durations of 1 day or longer, there was a 24-h delay in obtaining the RT-PCR test result. For a zero-day travel quarantine, the RT-PCR test was conducted 24 h before travel.

In particular, our analysis for November 21 illustrates that the minimum sufficient quarantine duration for destination countries with lower prevalence (Red status) had a median of four days, whereas in destination countries with high prevalence (Dark Red status) the median estimated quarantine duration was zero days. Compared to November 21, there was more diversity in the EU status among countries on August 8 (**Fig. S32**). Specifically, the estimated case burden of COVID-19 on August 8 indicates that there was one country in Green status, three in Amber, 13 in Red, and nine in Dark Red compared to the situation on November 21 when none of the 26 countries were Green or Amber status. For August 8, the minimum sufficient quarantine duration for destination countries with low prevalence (i.e., Green status) had a median of four days, moderate prevalence (i.e. Amber status) two days, whereas in destination countries with high prevalence (Dark Red status) the median estimated quarantine duration was zero days. Similarly, the estimated duration of travel quarantine required for travellers from origin countries with high prevalence (Dark Red status; median one days) was longer than that for travellers departing from origin countries with low prevalence (Green status; median zero days).

In addition to prevalence of disease, travel volume has a notable impact on quarantine and testing strategies that are sufficient to enable travel without increasing within-country transmission (**Fig. S2**). For instance, a comparison of Dark Red status Norway and Greece reveals that sufficient quarantines to visit Greece would need to be longer due to differences in typical travel asymmetries in and out of the country (**Fig. 2D; Fig. S1**). For a few destination countries (such as Spain, Malta, Cyprus, Portugal and Greece), we found that even a travel quarantine duration of 14 days would be insufficient to keep within-country imminent infections equivalent to or lower than that achieved by a complete travel ban (**Fig. 2I; Fig. S1**). This result can arise when there is substantial asymmetry in the number of travellers abroad. For example, travel asymmetry between the UKto Greece is high; consequently, a ban on travel for visitors from the UK would be required to ensure transmission within Greece is not increased. A large number of travellers in the country and fewer residents abroad leads to serious challenges in minimising quarantine and testing strategies to maintain travel at normal levels. These results indicate that a multitude of factors can simultaneously influence the importation of infection, impacting the minimum sufficient quarantine durations. Sensitivity analysis of the minimum sufficient quarantine durations for each parameter revealed that the natural immunity and number of daily travellers in both the destination and origin countries; daily incidence per capita and disease prevalence in the destination country; as well as vaccine-elicited immunity in the origin country exhibited substantial impact on the quarantine duration (**Fig. S2**). The number of travellers abroad in the destination and origin country, vaccine-elicited immunity in the destination country, and disease prevalence in the origin country had moderate impact, while age demographics and population sizes had negligible effects on the duration of travel quarantine (**Fig. S2**). As the pandemic situation evolves in each country, the changes in population immunity and disease prevalence will impact the minimum sufficient quarantine (**Fig. S1 vs Fig S32**). The estimates are fairly stable on a monthly time scale; the minimum sufficient quarantine durations calculated as of October 3 would not lead to imminent infection greater than border closure for 68·9% of country pairings through to November 21 (**Fig. S26**). To maintain a relevant minimum sufficient quarantine for a country, access to an open source interactive spreadsheet has been made available (**Supplementary File**).

### Variants of concern

Considering variants of concern, we quantified the sufficient durations of travel quarantine when an RT-PCR test is conducted on exit from quarantine with the goal of no net increase in the incidence of Delta G/478K.V1 ^42^ and Omicron B.1.1.529+BA for estimates of their circulation within the destination and origin countries. ^40^

As of November 21, the highly contagious Delta G/478K.V1 variant was widespread across much of Europe. Therefore, among countries with surveillance enabling estimation of the number of cases attributed to the variant, sufficient quarantine and testing is similar for these variants to that determined for general transmission without stratification by variants (**Fig. 2I** vs. **Fig. 3A**). In contrast, the emerging Omicron B.1.1.529+BA variant and all other variants excluding Delta G/478K.V1 and Omicron B.1.1.529+BA were at relatively lower frequency in most European countries, with much greater variance in prevalence. Consequently, sufficient quarantine durations would be longer for these variants than that determined in a general analysis of COVID-19 transmission (**Fig. 3B–C** vs. **Fig. 2I** and **Fig. 3A**). Combining multiple variants of concern leads to sufficient quarantines for each origin-destination pair determined by the maximum of those deemed sufficient for each variant. Consequently, incorporating additional variants of concern into the goal of assuring no additional infections due to travel leads to potentially longer quarantines and significant travel restrictions (**Fig. 3D** vs **Fig. 2I** and **Fig. 3A–C** and **Fig. S3** vs **Fig. S1**). Sufficient durations of travel quarantine increased substantially when making policy on the basis of the multiple variants of concern compared to evaluation that does not distinguish between variants (median of 3 days vs median of zero days; 32·2% travel ban vs 1·5% travel ban; **Fig. S3** vs **Fig. S1**).

**Figure 3.**
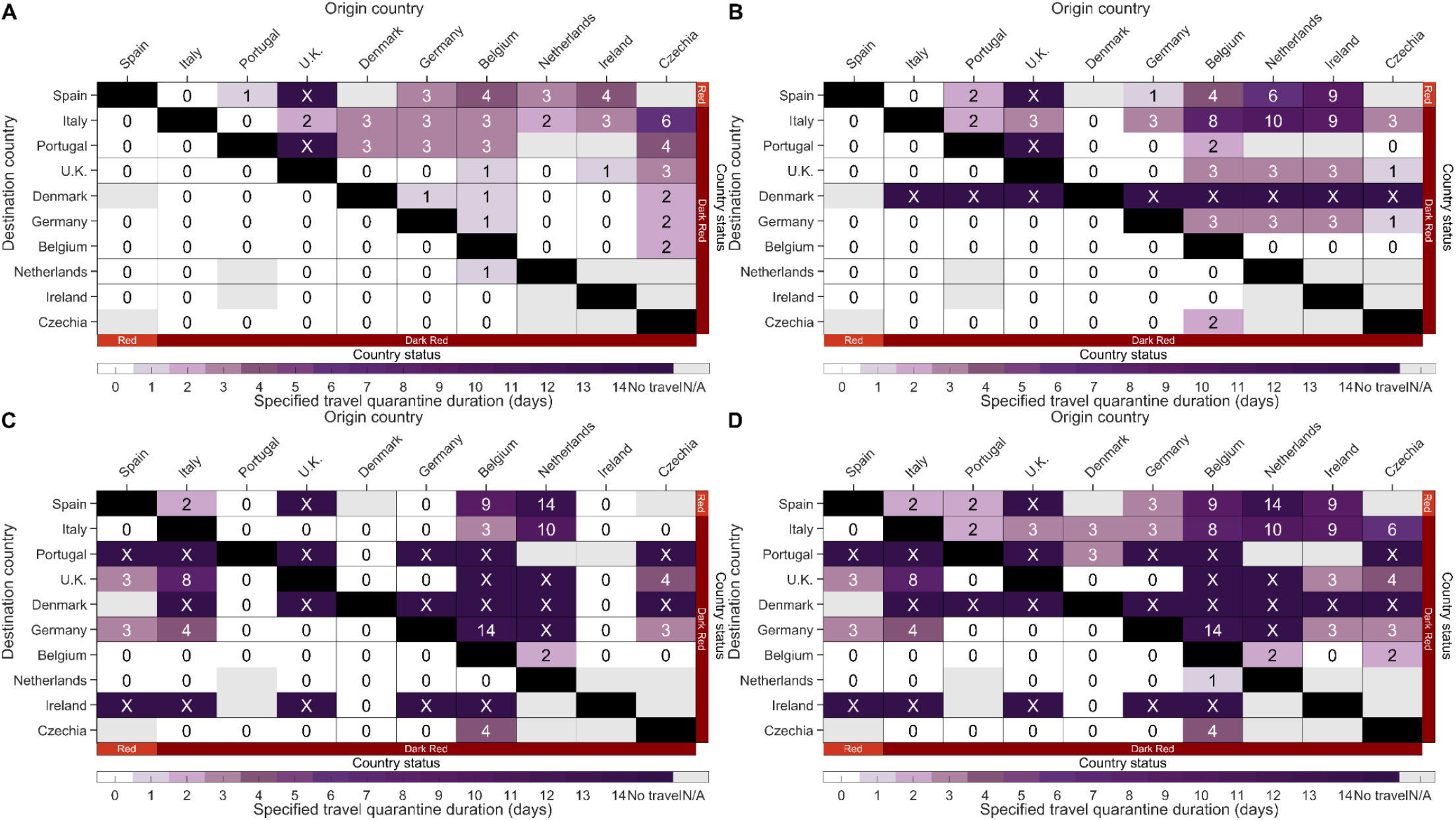
The estimated minimum duration of travel quarantine for specified origin-destination country pairs that reduces imminent infections to be equivalent to banning travel when considering variants of concern for the pandemic as of November 21, 2021. Specifying age-dependent vaccine effectiveness and proportion of asymptomatic infections, as well as country-specific demographics, incidence, prevalence of non-isolated infections, percentage of variants of concern, vaccine coverage, natural immunity and travel flow, we determine the minimum sufficient duration of travel quarantine with an RT-PCR test on exit from quarantine (colour gradient) that should be stated by the destination country for individuals arriving from the origin country when considering (**A**) transmission of the variant of concern Delta G/478K.V1, (**B**) transmission of the variant of concern Omicron B.1.1.529+BA, (**C**) transmission of the variants except Delta G/478K.V1 and Alpha 202012/01 GRY, and (**D**) general transmission and transmission of the variants of concern Delta G/478K.V1 and Omicron B.1.1.529+BA based on data for November 21, 2021. We consider travel quarantine durations of zero-days (white) to no travel (dark purple, i.e., specified quarantine can exceed 14 days). Within-country travel quarantine is not evaluated in the analysis (black). Travel flow data was not available for all country pairs (grey). The countries are ranked based on their estimated incidence per 100,000 over the last two weeks (November 8 to November 21) and stratified based on the European Union country classification system: Green, < 25 cases per 100,000; Amber, 25 to 150 cases per 100,000; Red, 150–500 cases per 100,000; and Dark Red, > 500 cases per 100,000. For travel quarantine durations of 1 day or longer, there was a 24-h delay in obtaining the RT-PCR test result. For a zero-day travel quarantine, the RT-PCR test was conducted 24 h before travel.

### Travel quarantine based on European Union traffic-light system

We calculated sufficient quarantines based on the input used by the EU COVID risk classification system, ^47^ specifying population sizes, pre-existing immunity, travel duration, and travel flow as the average of obtained data on the countries analysed and age-specific vaccine coverage observed within Europe. We found that for origin countries at equal or lower COVID-19 status than the destination country, a zero-day travel quarantine with RT-PCR test is equivalent to or better than travel ban (**Fig. 4**). The specified duration of travel quarantine extends as the ratio of the two-week case count per capita in the origin country to the destination country increases (**Fig. 4**).

**Figure 4.**
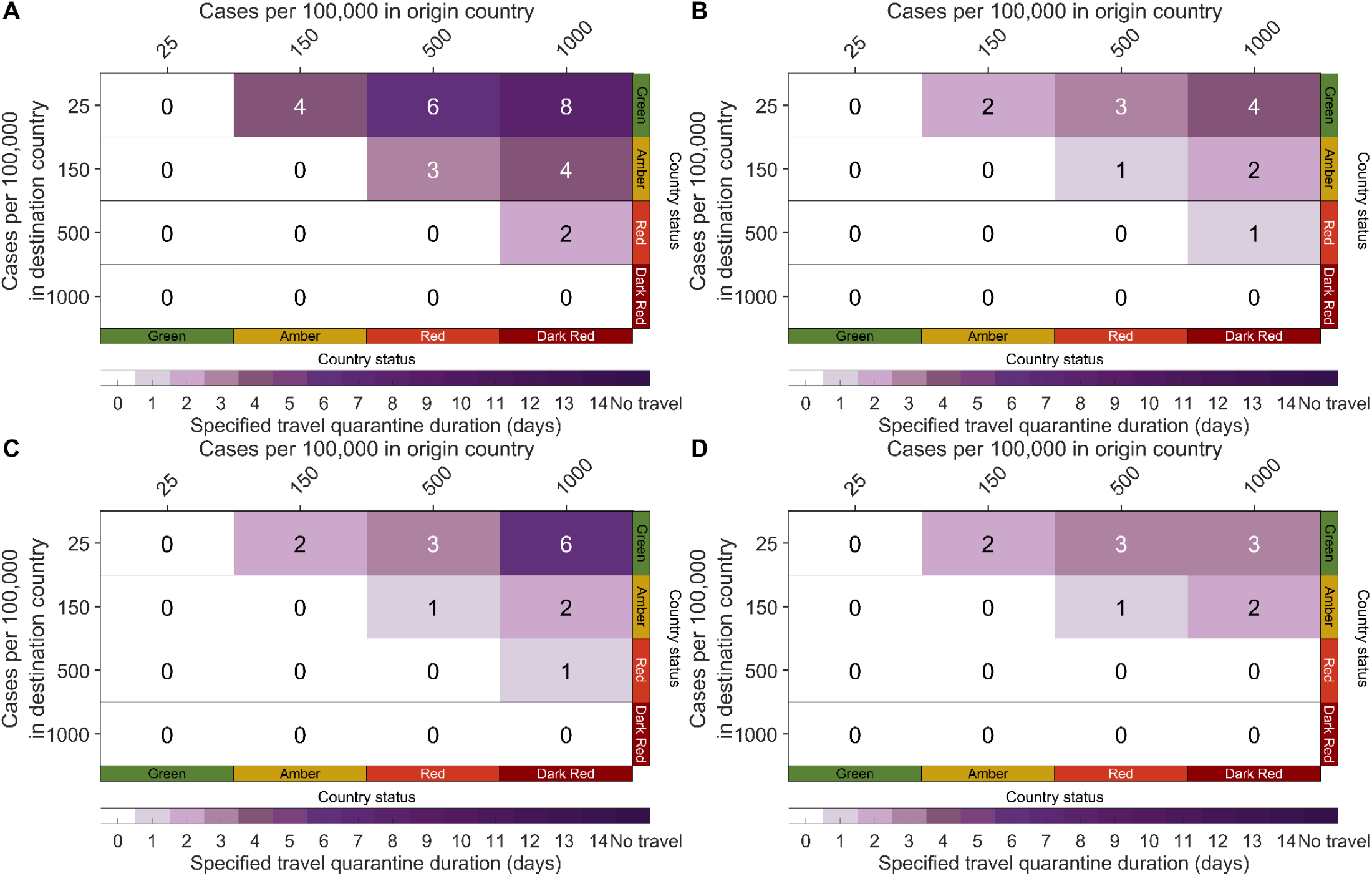
The estimated minimum duration of travel quarantine for origin-destination country pairs sufficient to prevent additional within-country imminent infections due to travel, using prevalence associated with European Union traffic-light categorization of COVID-19 risk. The minimum sufficient durations of travel quarantine (colour gradient) are calculated including (**A**) no testing, (**B**) an RT-PCR test on exit from quarantine, (**C**) a rapid antigen test on exit from quarantine, and (**D**) a rapid antigen test on both entry to and exit from quarantine, specifying age-dependent vaccine effectiveness and proportion of asymptomatic infections, average European age structure, 42% vaccine-acquired immunity, and 32% natural immunity, for origin countries whose EU traffic-light status is Green (25 cases per 100,000), Amber (150 cases per 100,000), Red (500 cases per 100,000), or Dark Red (1,000 cases per 100,000). For travel quarantine durations of one day or longer, there was a 24-h delay in obtaining the RT-PCR test result and no delay in obtaining the rapid antigen test. For a zero-day travel quarantine, the RT-PCR test was conducted 24-h before travel.

Because testing approaches and quarantine durations vary across different countries, we compared sufficient travel quarantine durations under alternative strategies of no testing, a RT-PCR test on exit, a rapid antigen test on exit, and a rapid antigen test on both entry and exit. Regardless of the testing approach, discrete tier categorization as opposed to quantitative calculation led to sufficiency of zero-day travel quarantine in any origin-destination pair for which the destination EU traffic-light status was equivalent or worse than the origin EU traffic-light status (**Fig. 4**; cf. **Fig. 2G**). Among origin countries with greater point prevalence than the destination country, the median minimum sufficient quarantine duration with no test was four days, with a range of two to eight days (**Fig. 4A**). With an RT-PCR test on exit from quarantine, the median duration of travel quarantine was two days, and durations ranged from one to four days (**Fig. 4B**). With the exception of a Green destination and Dark-Red origin country, switching to a less-sensitive but inexpensive and logistically flexible rapid antigen test yields results identical to those derived using an RT-PCR test on exit (**Fig. 4C** vs. **Fig. 4B**). Performing a rapid antigen test on entry to quarantine in addition to exit from quarantine allowed for a day shorter quarantine duration when the destination country is Green and the status of the origin country is Dark Red (**Fig. 4D** vs. **Fig. 4B**). Furthermore, quarantine for a destination country in Red and origin country in Dark Red can be eliminated (**Fig. 4D** vs. **Fig. 4B**). We also quantified the effects of these alternative testing strategies in the context of our richly parameterized country-pair analysis (**Fig. S4–S6**), and found that these trends were largely consistent with the results obtained from a tier-based analysis (**Fig. S7** and **Fig. S33**). Specifically, when the origin country was assigned lower-risk status than the destination country, the durations of quarantine for travellers from the origin country were equal to the median for equivalent country pairs in the tier-based analysis (**Fig. S7** and **Fig. S33**). When the origin country was assigned higher-risk status, the tier-based analysis exhibited the same increasing trend of quarantine duration with increasing EU risk status as the country-pair analysis. The greatest discrepancy was associated with the strategy of no test for a Redstatus destination and a Dark Red status origin for the epidemic status as of November 21 (**Fig. S7B**), where the median of quarantine duration from the country-pair analysis was six days longer than that determined from the tier-based analysis.

### Scenario analysis

We quantified the impact of longer incubation periods of 8·29 days and 11·66 days on the sufficient quarantine durations compared to the baseline incubation period of 5·72 days. In the country-pair analysis, the sufficient quarantine durations for an RT-PCR test on exit from quarantine did not change for 62·9% of the origin-destination pairs using an 8·29-day incubation period (**Fig. S1** vs. **Fig. S10**), and 54·4% remained unaltered for an 11·66-day incubation period (**Fig. S1** vs. **Fig. S15**). Among the pairs where the quarantine duration changed, the median quarantine duration for the incubation period of 8·29 days was one day longer than the median quarantine for the incubation period of 5·72 days, while median quarantine duration for the 11·66-day incubation period was three days longer. For the latter incubation period, 55·4% of country pairs could implement a sufficient 0-day quarantine with an RT-PCR test on exit, compared to 65·0% for the baseline incubation period of 5·72 days. With the EU traffic-light analysis, the minimum sufficient quarantine decreased at most two days when considering an incubation period of 8·29 days for all testing strategies (**Fig. 4** vs. **Fig. S14**). For the longer incubation period of 11·66 days, the minimum sufficient quarantine increased at most seven days (**Fig. 4** vs. **Fig. S19**).

In our baseline analysis, we performed a policy evaluation based on 100% adherence to self-isolation upon symptom onset and within quarantine. If adherence to self-isolation were to decrease from 100% to as low as 25%, we found that 14·5% of the estimated quarantine durations would change in the country-pair analysis with an RT-PCR test on exit from quarantine (**Fig. S20–S22**). As adherence to self-isolation diminishes, there is a greater chance that a country would have to enforce border closure. With an RT-PCR test on exit from quarantine, the proportion of origin-destination pairs that require no travel increases from 1·6% to 15·0% when self-isolation adherence declines from 100% to 75% (**Fig. S23**). At 25% adherence to self-isolation, 37·1% of the country pairs would require a travel ban—majority of which are imposed on origin countries with higher case burden over the last two weeks than the destination country (**Fig. S25**).

The average daily travel flow was informed by annual arrivals for all forms of paid accommodation. In an alternative scenario, we used annual air passenger transport between countries to measure average daily travel flow between countries. Among all country-pairings common to both data sets, travel flow informed by air passengers resulted in no quarantine for 49·5% and border closure for no country pairings compared to 64·9% and 1·7% respectively in the baseline analysis (**Fig. S27** vs **Fig. S1**). Comparing the quarantine durations of the two analyses, 49·8% of quarantine durations differed (**Fig. S27** vs **Fig. S1**). Among those that differed, we found that more stringent quarantines were suggested under travel flow informed by the number of air passengers (median of two day longer quarantine compared to baseline). Under the EU-tier based analysis, there was no change in the sufficient quarantine durations when using the air passenger transport between countries (**Fig. 4 vs Fig. S31**).

## Discussion

Here we have identified strategies of travel quarantine and testing that prevents within-country imminent transmission beyond what would still occur under a travel ban scenario. We conducted our analysis for a snapshot in time based on the epidemic situation observed on November 21, 2021 in 26 European countries. We demonstrated that quarantines for European destinations can be informed by country-specific prevalence, daily incidence, vaccine coverage, immunity, age-demographics, and travel flow from the country of origin. Our analysis for this specified time of the epidemic indicated that for nearly half of these European origin-destination country pairs, no quarantine or test is necessary to prevent increased within-country imminent transmission from travellers, and for the majority, a test with no quarantine would be sufficient. For many other country pairs, a travel quarantine of a few days combined with testing on exit would suffice. The duration of travel quarantine is influenced mainly by the relative prevalence of disease between the origin and destination countries, immunity in both countries, and the asymmetry of travel. With a goal of preventing the introduction of variants of concern, quarantine and testing strategies that are sufficient for the general case will usually help to prevent the rise of imminent VOC transmission within-country as well. The strategies diverge when variants of concern are infrequent and prevalence is heterogeneous on the international scale.

The incubation period of a disease often determines the standard quarantine durations. However, quarantine durations quantified from our analysis are generally shorter than those implemented in most countries. These short but non-trivial quarantine durations arise from a multitude of factors. Firstly, unlike standard quarantines that aim for no post-quarantine transmission, our framework only requires quarantines where travel does not elevate infection rates in the destination country relative to border closure. Also, infected travellers are more likely to enter quarantine later in infection compared to traced contacts who enter during the early stages postinfection. Other external factors, such as prevalence, travel flow, and immunity, also influence these quarantine durations. For example, the duration of standard quarantines can be insufficient to suppress within-country transmission at times. Therefore, accounting for the duration of the incubation period as well as external factors is essential for determining the appropriate travel quarantine strategy.

The sufficient travel quarantines for European countries presented here are informed by the epidemic scenario on November 21. The framework presented here can be used to guide short-term policy recommendations over the course of the epidemic by reevaluating quarantine durations based on changes in the epidemic scenario. Heterogeneity in the sufficient quarantine durations determined for country pairs across Europe demonstrates the importance of accounting for country-specific characteristics, such as asymmetry in travel. These estimates could, in principle, be updated regularly using our country-pair model based on the evolving epidemic situations in each country (**Supplementary File**). However, implementing travel quarantine and testing policies that are specific to many input parameters, dependent on country of origin as well as destination—and that change dynamically as waves of the pandemic strike and recede—is logistically challenging. Indeed, some countries have specified a single quarantine duration for any traveller entering their country. ^48,49^ A single policy may be unduly restrictive—or in some cases may not be restrictive enough. An intermediate approach is to simplify input parameters and discretize country classification in accordance with existing frameworks, such as the incidence-based EU traffic-light system. By considering European averages for other parameters, we demonstrated the feasibility of mapping our more highly parameterized approach to the EU traffic-light system.

The agreement between results of our highly parameterized country-pair analysis and those we obtained from mapping to the prevalence-only EU traffic-light categories is imperfect. For example, Greece (Dark Red EU-status) requires a travel ban for travel from the UK (Dark Red EU-status) in the country-pair model, whereas no quarantine is indicated for the EU traffic-light model. The EU traffic-light model relies only on differences in incidence. Expanding beyond the EU traffic-light model to include country-specific vaccination coverage is possible, but at the expense of overwhelming policy makers with numerous scenarios and complex implementation. However, any individual country can still improve the accuracy of the tier-based model by categorising the rest of Europe in the EU tier-system and parameterizing the model (e.g., travel flow, immunity, and demographics) specific to their country as well as European averages for origin countries. Therefore, our results from the EU traffic-light model provide guidance regarding a simplified approach that aligns with current practises and illustrates easy communication of appropriate travel quarantine duration. Together, the two models provide insight as to an effective quarantine duration for travel within Europe, and provide analytical justification for decision-making that can otherwise be politicised rather than evidence-based.

The sufficient quarantine durations are influenced by prevalence and immunity in the two countries, as well as the asymmetry in travel flow, consistent with previous studies highlighting importation risks are determined by multiple simultaneous factors. ^15^ There are uncertainties associated with travel as well as the epidemiological state of a country that may alter the length of sufficient quarantine duration. Where disease prevalences and immunity levels are unknown or poorly monitored, countries may adopt a precautionary principle and take a restrictive approach to border control. Because travel volumes during the pandemic are uncertain, we used the annual number of arrivals to paid accommodations in 2019 to inform a fixed daily travel flow, which underestimates the overall travel flow across country borders during 2019. However, we expect that it would still be greater than the reduced travel flow during the pandemic. Thus, our estimates for the sufficient travel quarantine can be considered conservative relative to the extent of travel during the pandemic. While we present our results based on average daily travel flow, seasonal fluctuations in travel could distort these average relationships, affecting imminent transmissions as seen during 2020 summer travel in Europe. ^50^ Our sensitivity analysis indicates that a disproportionate increase of travellers into a destination country requires longer quarantines, while quarantine restrictions can be lessened when residents are more willing to travel. Therefore, re-evaluation of the sufficient quarantine to account for changes in travel flow between countries may be necessary to ensure continued safe travel.

Our analyses focused on within-country imminent infection in comparison to travel ban, justifying the imposition of quarantine and testing strategies only insofar as they reduce within-country transmission more than a travel ban policy. However, whether the sufficient quarantine and testing strategies identified here are worthwhile from a public health or cost-effectiveness perspective requires additional consideration. For instance, high rates of hospitalisation can overwhelm the healthcare infrastructure in a country. Stringent disease control measures were found to slow the local disease progression, ^8^ delaying the peak in incidence and potentially preventing a crash of the local health system. Thus, policymakers may consider applying travel quarantines to ensure that the hospitalisation rate remains manageable. Hospitalisation rates in the destination country are likely not influenced by traveller origin or even by quarantine duration because of the relatively small numbers of travellers when compared with widespread infection among the resident population during a fully emerged pandemic. ^51^ In this circumstance, travel quarantines will have an almost trivial impact on within-country imminent infections and a proportionately small contribution toward hospitalisation rate. For countries looking to adopt a zero-COVID policy, the minimum sufficient quarantines determined from this framework are inadequate because they allow for a level of transmission similar to that estimated under a travel ban and do not halt the importation of cases. A balanced consideration of quarantine duration should come both from an understanding of the relative impact of travel on infection and on the public health consequences of infections.

With the implementation and relaxation of different disease-control efforts—along with differences in contact patterns and age demographics—the reproduction number will differ across countries, and constantly change over the course of the pandemic.^52^ The minimum sufficient quarantine duration is estimated under the specification that the implicit contact patterns were similar between residents and non-residents. This homogeneity impacts the number of infections occurring in and produced from travellers. Accounting for heterogeneity in the implicit contact patterns between travellers and residents can increase or decrease the minimum sufficient quarantine duration. Indeed, the number of infections that will occur among the residents of the destination country is influenced by the reproduction number. However, our estimates of minimum sufficient travel quarantine is independent of any changes in the reproduction number, as this number factors out of the calculation (**Supplementary material: Sufficient travel quarantine**). Therefore, we chose to use a homogeneous reproduction number in the absence of self-isolation and vaccination for all countries. Our choice to use the basic reproduction number (and not the effective reproduction number) is conservative as it does not account for interventions.

Our analysis focuses on the European region, where SARS-CoV-2 infection is already widespread. Globally, there are only a few instances where the virus has been forestalled at a national ^35^ or geographic border. ^53^ Over the course of the pandemic, for instance, New Zealand has effectively maintained a low number of COVID-19 cases. ^35^ Travel to New Zealand was restricted to citizens of that country, who were only allowed entry upon a negative test prior to departure, followed by a highly effective 14-day quarantine with two negative tests.^10,54^ New Zealand aims to maintain closed borders with most of the globe until there is sufficient vaccine protection. ^54,55^ This strategy is supported by our analysis for a country with very low or zero daily incidence and prevalence.

Similarly, our approach would recommend border closure by many countries early in the pandemic before global spread of infection—a closure that did not happen for a variety of geopolitical reasons, but also because of technological and supply limitations that prevented rapid global testing. Even after several months into the pandemic, infections may still be under-reported due to limited health resources and a high proportion of asymptomatic infections. ^56–59^ In part for this reason, the EU traffic-light system elevates their assessment of risk for countries that have high rates of test positivity. Disproportionate under-reporting of infections in the origin country relative to the destination would limit the informativeness of our analyses using only reported infections. To overcome this limitation, an analytical layer could be added that adjusts reported prevalence by test positivity. ^59,60^

As the pandemic progresses, quarantine decisions may shift completely toward prevention of one or multiple variants of concern that are more transmissible or can escape natural and vaccine-mediated immunity, ^17^ that may justify imposition of highly restrictive quarantine and testing strategies, or even border closure. Just as in the early COVID-19 pandemic, ^61^ a strong implementation of travel restrictions could forestall specific variants of concern at the border. However, detection of such variants can be substantially delayed relative to their introduction or establishment in a country,^62,63^ which can lessen the effectiveness of border closure. Moreover, early detection of low-frequency emerging variants is unlikely—even by an aggressive surveillance effort. Consequently, our estimates of minimum sufficient quarantine durations for variants of concern should be understood as applying only to those already identified. Analysis on quarantines aimed to prevent the invasion of an emerging variant would be useful, but there is no methodology evident to us for doing so. Rapid and accurate identification of variant properties is crucial for policy decisions, because attempting to forestall even a low-frequency variant would often entail near-complete border closure, which may be unnecessary if the public health threat of the variant is minor compared to other circulating variants. National efforts to address disease spread would be tremendously aided by genomic approaches to rapidly and accurately assess variant properties. Building international capacity for such surveillance should be a priority for all nations, so that decisions can be informed and judicious.

Our study focused on specifying quarantine durations for travel between only 26 European countries. However, some guidance for other countries may be obtained by finding the European country pairs most similar in circumstance to any origin-destination pair of interest. Furthermore, our modelling framework can directly inform travel quarantine duration between any two countries using country-specific input data. The applicability of our approach is not limited to travel between countries—where border controls are typically strongest and easiest to impose—but can be applied to any distinct populations for which disease prevalence, travel flow, and other input data can be quantified. Ultimately, international quarantine and testing should not be considered a substitute for national policy preventing the spread of disease at a more localised scale within the country itself. Even if a 0-day quarantine is sufficient between countries, widespread local travel may lead to infections where there were previously none. ^64,65^

The expected success of quarantine and testing can be undermined by non-compliance. We found that the level of adherence to self-isolation upon symptom onset had less impact on the estimated minimum sufficient quarantine duration than decreasing adherence to the quarantine policy. As adherence to quarantine decreases, the duration of sufficient quarantines increases. It is worth noting that non-compliance is increased by the stringency of the requirement, so that it is possible to achieve less by requiring more. For example, some entrants to Canada opt not to comply with its 14-day quarantine requirement. ^66–68^ As the first days of quarantine are often the most important to the prevention of SARS-CoV-2 transmission, ^10^ it can be disadvantageous to public health goals to require a longer quarantine that elicits poor adherence instead of a shorter quarantine with improved compliance. Furthermore, shorter country-specific quarantion durations could free up resources for measures that improve compliance such as providing low-cost, safe accommodations and convenient, rapid, and accurate testing.

There is some controversy as to whether antigen testing can substitute for RT-PCR testing. ^69–71^ Rapid antigen tests have the advantage of faster turnaround time, but it comes at the cost of lower sensitivity and specificity than the RT-PCR tests. Inferior specificity of rapid antigen tests would lead to a higher false-positive rate, but this effect on imminent infections will be negligible if nonzero. Nevertheless, false positives may incur additional expenses for travellers such as validation tests, opportunity costs, and psychological damage.^72^ With regard to their lower sensitivity, our results indicate that rapid antigen tests can play a significant role in effective quarantine and testing. The faster turnaround for the processing of antigen tests permits its use a day later during quarantine than the RT-PCR test. This feature enables its timing just before travel departure, while RT-PCR would need to be scheduled days in advance and an infection in its early stages may not be detected.^10^ We also showed that multiple rapid antigen tests could potentially be equivalent to or better than RT-PCR tests for prevention of post-quarantine transmission. Furthermore, use of an antigen test does not preclude one from sampling variants of concern, as positives can be referred for RT-PCR validation and genetic sequencing. However, the high volumes of testing required for travel quarantine poses challenges for utility of both RT-PCR and rapid antigen tests. The processing time of numerous RT-PCR samples could be made more feasible using high-throughput assays, while additional training for travellers may be required to reduce the number of missed cases from rapid antigen tests.

Our analytical approach was designed to determine sufficient quarantines during the global COVID-19 pandemic. However, the model can be reparameterized with the properties of other emerging diseases that may carry a global risk, to obtain policies of quarantine and testing that are sufficient to prevent increased within-country transmission. Border closure would be recommended during very early stages of pandemic spread to limit the global dissemination of an emerging disease from the epicentre, ^73^ but such a measure provides limited benefits once community transmission is established in the country. ^5^ As the trajectory of a pandemic continues to unfold and country-specific prevalence, circulation of variants, and potentially vaccination coverage changes, travel quarantine strategies can be adjusted to enable effective and judicious responses to new epidemiological conditions. Use of our model can provide an evidence-base approach to ameliorate policy decisions that often otherwise polarised entirely permissive or overly restrictive states, facilitating equitable and safe international travel conditions between countries.

## Data Availability

All data is referenced in the manuscript.

## Author contributions

CRW and AP contributed equally to this work. JPT conceived and designed the study. JPT developed the theory with contributions from CRW and AP. CRW and WSC assembled relevant data. CRW wrote computational code, and executed analyses with contributions from AP. All authors contributed to interpretation of results. CRW, AP, and JPT drafted the manuscript. MCF, BHS, SMM and APG contributed to revision of the draft manuscript. All authors approved the final version of the manuscript.

## Data availability

All data used in the analysis is referenced within the Methods and Supplementary Tables.

## Code availability

The computational MATLAB code is available online. ^74^ A supplementary Excel file is provided to determine the minimum sufficient quarantines (**Supplementary File**).

## Acknowledgements

JPT and APG gratefully acknowledge research funding from EasyJet. JPT gratefully acknowledges funding from the Elihu endowment and the Notsew Orm Sands Foundation. MCF gratefully acknowledges funding from the National Institutes of Health (5 K01 AI141576). APG gratefully acknowledges funding from the Burnett and Stender families’ endowment and the Notsew Orm Sands Foundation. SMM gratefully acknowledges funding from the Canadian Institutes of Health Research, and Natural Sciences and Engineering Research Council of Canada EIDM (Grant: MfPH).

## Editor note

The Lancet Group takes a neutral position with respect to territorial claims in published maps and institutional affiliations.

---

**Table 1.**
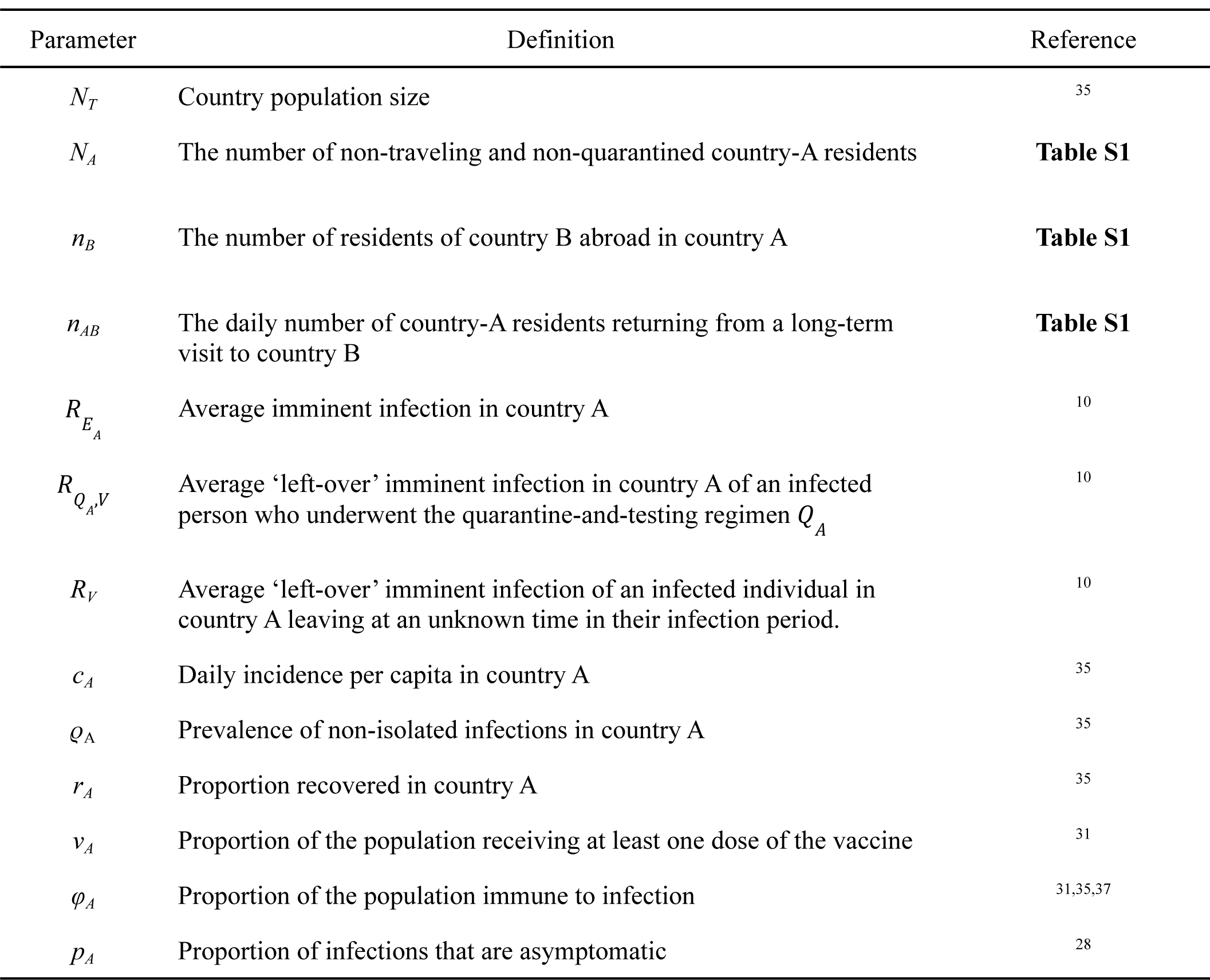
Parameter descriptions and references for determination of sufficient durations of travel quarantine

