## Supplementary Material for "Quarantine and testing strategies to ameliorate transmission due to travel during the COVID-19 pandemic: a modelling study"

### Table of Contents


**Supplementary methods** **3**
Sufficient travel quarantine 3
Age-dependent imminent infections 3
Variants of concern 4
Effective reproduction number 5
Non-traveller 6
Discounting infections for travellers leaving the destination country 6
Quarantine and testing for travellers 6
Zero day quarantine with RT-PCR test 7
Non-adherence to travel quarantine and testing 8
Infectivity profile 9
Temporal diagnostic sensitivity 9
RT-PCR test 9
Rapid antigen test 11
Parameterization 11
Travel 11
Vaccination and prevalence 13
Generalised approach using EU traffic-light system 14
Sensitivity analysis 15
Implementation 15

**Supplementary Figures** **16**

**Supplementary Tables** **50**

**References** **60**

#### Supplementary methods

##### ***Sufficient travel quarantine***

*Short-term stays*

For the calculation of the minimum sufficient travel quarantine as described in the main text, we calculated the effective reproduction number under the assumption that prevalence in travellers rises to the prevalence in the country visited by the time of return (i.e., long-term stay). Compared to the assumption of long-term stays, short-term stays would result in a lower number of secondary infections due to the infected individuals' departure from the country. Similarly, the risk of infection for uninfected travellers entering a high prevalence country for a short-duration would be lower on average. Thus, accounting for short-term stays would result in shorter quarantine durations than that estimated by our theory in the main text.

*Age-dependent imminent infections*

We extended the inequality for the daily new within-country imminent infections (Eq. 1 in main text) to account for age-dependent disease characteristics for $N$ age classes, as

$$\left(1 - \varphi_A\right)\sum_{j=1}^{N} \lambda_{0,j} \geq \left(1 - \varphi_A\right)\sum_{j=1}^{N}\left(\lambda_{1,j} + \lambda_{2,j} - \lambda_{3,j} + \lambda_{4,j} + \lambda_{5,j} - \lambda_{6,j}\right), \quad \text{(S1)}$$

where $\varphi_A = \sum_{j=1}^{N} w_{A,j}\varphi_{A,j}$, $w_{A,j}$ is the proportion of population A in age-class $j$ and $\varphi_{A,j}$ is the level of immunity in country A among those in age-class $j$. We summarise the terms of the inequality (Eq 1 and Eq S1) using indexed symbols (**Table S4**). Specifically, we simplify the different terms of the expressions to reflect age-dependent disease characteristics using $\lambda_{k,j}$ to denote the $k^{th}$ term for age class $j$ (**Table S4**).

For the effective reproduction number in the absence of self-isolation and vaccination in our theory, we conservatively used the basic reproduction number for Europe ($R = 3$). [1] As the infectivity curve over time is obtained by multiplying the basic reproduction number with relative infectivity over the disease progression (see **Supplementary**

**methods: Effective reproduction number**), $R$ from each of the effective reproduction numbers ($R_{E_A}$, $R_V$, and $R_{Q_A,V}$) in the inequality factors out. Thus, the minimum sufficient travel quarantine duration calculated is independent of the basic reproduction number.

*Variants of concern*

As an additional analysis, we integrate variants of concern into the calculation of the duration of the minimum sufficient quarantine. Specifying that $\theta_{k,A}$ proportion of infections are of variant k in country A, portion of the entire population immune to the infection $\varphi_{A,k} = \sum_{j=1}^{N} w_{A,j}\varphi_{A,k,j}$, increasing transmission by factor $\tau_k$, the inequalities determining the number of daily new within-country imminent infections in destination country *A* when interacting with origin country *B* is

$$(1 + \tau_k)(1 - \varphi_{k,A}) \sum_{j=1}^{N} \theta_{k,A} \cdot \lambda_{0,j} \geq (1 + \tau_k)(1 - \varphi_{k,A}) \sum_{j=1}^{N} \left( \theta_{k,A}\lambda_{1,j} + \theta_{k,A}\lambda_{2,j} - \theta_{k,A}\lambda_{3,j} + \theta_{k,B}\lambda_{4,j} + \theta_{k,B}\lambda_{5,j} - \theta_{k,A}\lambda_{6,j} \right).$$

Specifying $N_V$ variants of concern, the proportion of infections from general transmission is $\theta_{1,A} = 1 - \sum_{j=1}^{N_V} \theta_{(j+1),A}$.

When evaluating the minimum sufficient travel quarantine duration for multiple variants of concern simultaneously, we specify it to be the duration in which all inequalities for the variants of interest are satisfied (i.e. the maximum of all the minimum travel quarantine durations). Alternatively, one could evaluate sufficient quarantine durations by aggregating transmission from each VOC. However, examining imminent infections of each VOC independently provides a more conservative outcome and allows countries to focus on individual VOCs that may be of certain interest. For example, one VOC may have the ability to evade immunity and be highly transmissible but cause less severe disease, while another VOC causes more severe disease but less easily transmissible.

***Effective reproduction number***

Given the relative infectivity over the course of the disease $f(t)$ (**Supplementary methods: Infectivity profile**) , and the duration of the disease, $t_E$ , we denote the infectivity over time as

$$r(t) = \begin{cases} Rf(t) & t \leq t_E, \\ 0 & t > t_E \end{cases} .$$

The reproduction number is dependent on the integral of the infectivity profile and is not parameterized with contact patterns or age susceptibility of the disease. Thus, the average number of secondary infections in the absence of self-isolation or other interventions is then

$$R = \int_0^\infty r(t)\ \mathrm{d}t.$$

A proportion of infections $p_A$ are asymptomatic, with the remaining $1 - p_A$ developing symptoms $t_S$ days after infection (i.e. the incubation period). For an asymptomatic individual, the transmission over time is $r_A(t) = r(t)$. Once an individual exhibits symptoms, they may enter isolation. To model the isolation of infected individuals for whom symptoms manifest, we denote their transmission over time as $r_S(t) = r(t)\cdot(1\text{-}H(t))$, where the appearance of symptom post infection is indicated using a binary step function,

$$H(t) = \begin{cases} 0, & t < t_s, \\ 1, & t \geq t_s \end{cases} .$$

In our base case analysis, we assume that all symptomatic individuals isolate upon symptom onset. In a scenario analysis, a proportion $\alpha_I$ of symptomatic individuals are assumed to adhere to self-isolation, with non-adhering individuals still able to transmit the disease after symptom onset. Scenarios of 75%, 50%, and 25% adherence to self-isolation upon symptom onset were examined.

*Non-traveller*

Specifying that a proportion $\alpha_I$ self-isolate on symptom onset, we calculate the effective reproduction number for a soon-to-be symptomatic case as

$$R_S = \alpha_I \int_{t=0}^{\infty} r_S(t)\mathrm{d}t + (1 - \alpha_I) \int_{t=0}^{\infty} r(t)\mathrm{d}t.$$

For asymptomatics we calculate the effective reproduction number as

$$R_A = \int_{t=0}^{\infty} r(t)\mathrm{d}t.$$

Thus, the effective reproduction number including non-immunological public health measures in country A is

$$R_{E_A} = (1 - p_A)R_S + p_A R_A. \tag{S2}$$

*Discounting infections for travellers leaving the destination country*

Some individuals travelling abroad will be infectious and will no longer contribute to transmission within the country. The remaining amount of transmission for these individuals is analogous to a zero-day quarantine with no test. The expected remaining transmission for a soon-to-be symptomatic case is

$$R_{V,S} = \alpha_I\left(\frac{1}{t_S}\int_{u=0}^{t_S}\int_{t=0}^{\infty} r_S(t + u)\mathrm{d}t\,\mathrm{d}u\right) + (1 - \alpha_I)\left(\frac{1}{t_S}\int_{u=0}^{t_S}\int_{t=0}^{\infty} r(t + u)\mathrm{d}t\,\mathrm{d}u\right).$$

For asymptomatic carriers,

$$R_{V,A} = \frac{1}{t_E}\int_{u=0}^{t_E}\int_{t=0}^{\infty} r(t + u)\mathrm{d}t\,\mathrm{d}u.$$

Therefore, the expected remaining transmission for infected travellers is

$$R_V = (1 - p_A)R_{V,S} + p_A R_{V,A}. \tag{S3}$$

*Quarantine and testing for travellers*

We model travellers entering quarantine randomly over the period in which they do not exhibit symptoms and assume that resident and traveller interactions are the same. The diagnostic sensitivity of a test at time *t* is denoted *s*(*t*) (**Supplementary methods: Temporal diagnostic sensitivity**). Individuals who are showing symptoms are not eligible for quarantine and instead are placed in isolation. For a specified duration of quarantine ($q$), delay in obtaining test result ($d_t$) testing occurring over the course of quarantine ($t_i$), and $\alpha_I$ isolating upon symptom onset (assuming isolation is enforced if symptoms present during quarantine), the expected post-quarantine transmission for a soon-to-be symptomatic case who is tested for disease at any time $0 \leq t_n \leq q - d_t$ during the quarantine is

$$R_{Q,V,S}(q) = \alpha_I \left( \frac{1}{t_S} \int_{u=0}^{t_S} \int_{t=q}^{\infty} r_S(t+u) \cdot \prod_{n=1}^{N} \left(1 - s\left(t_n + u\right)\right) \mathrm{d}t\, \mathrm{d}u \right)$$

$$+ (1-\alpha_I) \left( \frac{1}{t_S} \int_{u=0}^{\max(t_S - q, 0)} \int_{t=q}^{\infty} r(t+u) \cdot \prod_{n=1}^{N} \left(1 - s\left(t_n + u\right)\right) \mathrm{d}t\, \mathrm{d}u \right).$$

For asymptomatic carriers,

$$R_{Q,V,A}(q) = \frac{1}{t_E} \int_{u=0}^{t_E} \int_{t=q}^{\infty} r(t+u) \cdot \prod_{n=1}^{N} \left(1 - s\left(t_n + u\right)\right) \mathrm{d}t\, \mathrm{d}u.$$

Therefore, the expected post-quarantine transmission is

$$R_{Q_A V}(q) = (1 - p_A) R_{Q,V,S}(q) + p_A R_{Q,V,A}(q). \quad \text{(S4)}$$

*Zero day quarantine with RT-PCR test*

For the strategy of testing with RT-PCR but no quarantine, equation (S4) cannot be used to estimate the expected post-quarantine infection ($R_{Q_A V}$) because delays are associated with obtaining results from RT-PCR tests. Thus, we account for the potential that infection can occur between the time of test sample and result. If testing occurs at time $\delta \leq v \leq t_S$ prior to travel, individuals who are infected subsequent to the test and prior to travel—and who will exhibit symptoms at time $t_S$—will contribute

$$R_{S,w^{\mapsto}}(\delta) = \alpha_I \left( \frac{1}{\delta} \int_{u=0}^{\delta} \int_{t=\delta-u}^{\infty} r_S(t) \mathrm{d}u\, \mathrm{d}t \right) + (1-\alpha_I) \left( \frac{1}{\delta} \int_{u=0}^{\delta} \int_{t=\delta-u}^{\infty} r(t) \mathrm{d}u\, \mathrm{d}t \right)$$

In contrast, individuals who are infected prior to the test (and prior to travel)—and who will exhibit symptoms at time $t_S$—will contribute

$$R_{S,x^{\mapsto}}(\delta) = \alpha_I\left(\frac{1}{t_S-\delta}\int_{u=0}^{t_S-\delta}\int_{t=u+\delta}^{\infty}(1-s(u))\cdot r_S(t)\mathrm{d}u\,\mathrm{d}t\right) + (1-\alpha_I)\left(\frac{1}{t_S-\delta}\int_{u=0}^{t_S-\delta}\int_{t=u+\delta}^{\infty}(1-s(u))\cdot r(t)\mathrm{d}u\,\mathrm{d}t\right)$$

Because these events are exhaustive and exclusive, the total post-travel transmission from an infected traveller who will manifest symptoms, based on when the pre-travel test is administered, is

$$R_{T,V,S}(\delta) = \frac{\delta}{t_S}R_{S,w^{\mapsto}}(\delta) + \frac{t_S-\delta}{t_S}R_{S,x^{\mapsto}}(\delta).$$

Similarly for individuals who will not exhibit symptoms or self-isolate without a positive test result, if testing occurs $\delta \leq v \leq t_S \leq t_E$ days prior to travel, they will contribute

$$R_{A,w^{\mapsto}}(\delta) = \frac{1}{\delta}\int_{u=0}^{\delta}\int_{t=\delta-u}^{t_E} r(t)\mathrm{d}u\,\mathrm{d}t$$

In contrast, individuals who are infected prior to the test (and prior to travel) and not exhibit symptoms will contribute

$$R_{A,x^{\mapsto}}(w) = \frac{1}{t_E-\delta}\int_{u=0}^{t_E-\delta}\int_{t=u+\delta}^{t_E}(1-s(u))\cdot r(t)\mathrm{d}u\,\mathrm{d}t.$$

Therefore, the extent of transmission after the pre-travel test for asymptomatics is

$$R_{T,V,A}(\delta) = \frac{\delta}{t_E}R_{A,w^{\mapsto}}(\delta) + \frac{t_E-\delta}{t_E}R_{A,x^{\mapsto}}(\delta).$$

Incorporating both symptomatic and asymptomatic infections, the total post-travel transmission from infected travellers for the strategy of no quarantine and testing with RT-PCR is

$$R_{Q_A V}(0) = (1-p_A)R_{T,V,S}(\delta) + p_A R_{T,V,A}(\delta).$$

*Non-adherence to travel quarantine and testing*

In our base case scenario, adherence to quarantine was assumed to be 100%. However, we also evaluated quarantine durations when there is imperfect adherence to quarantine. For individuals who do not adhere to quarantine, we assume that the extent of transmission upon arrival to the destination country is equivalent to the scenario of no test and no quarantine for those not adhering to the quarantine policy. Specifying $\alpha_Q$ adherence to quarantine, the total post-travel transmission from infected travellers is

$$\overline{R_{Q_A V}}(q) = \alpha_Q R_{Q_A V}(q) + (1 - \alpha_Q) R_V.$$

***Infectivity profile***

We informed the relative infectivity profile of an infected individual on average *f*($t$) based on 77 transmission pairs of COVID-19 cases from the computational code provided by He et al [2,3], specifying a latent period of 2.9 days and an incubation period of 5.72 days [4–6]. The duration of the disease time course infectivity is 20 days after symptom onset [7–9], after which we specify the infectivity to be zero (i.e. $r(t)$=0). Over the duration of the infectivity profile, 53.5% of the infections happen before symptom onset and by 10 days after symptom onset 98.2% of infections have occurred (**Fig. S22**). Similarly, we constructed the relative infectivity profile for an incubation period of 8.29 days [10] and 11.66 day to perform a scenario analysis for assessing the impact of length of incubation period (**Fig. S22**).

We inferred the longer incubation period of 11.66 days from a meta-analysis study. [11] For our distribution of a longer incubation period, we specified a log-normal distribution with a median of 11.60 days, based on the reported median of the 95th percentile in the study.[11] We estimated the standard deviation of this log-Normal distribution by fitting its 2.5th and 97.5th percentile to the minimum and maximum values reported for the 95th percentile of the incubation period (9.49–14.20 days). From this estimated distribution, we obtained an average incubation period of 11.66 days.

***Temporal diagnostic sensitivity***

For quarantine and testing strategies, we considered both RT-PCR test and rapid antigen test. We calculated their temporal diagnostic sensitivity as follows.

*RT-PCR test*

To determine the diagnostic sensitivity of the RT-PCR assay, we use serial testing data conducted within the healthcare setting [12]. We use a similar methodology as Hellewell et al [12] to infer the diagnostic sensitivity, except we specified the diagnostic sensitivity function to be represented by the log-Normal probability density function that peaks at the time in which infectivity peaks. Specifically, the probability the RT-PCR test is positive (i.e. diagnostic sensitivity) at time $t$ is expressed as

$$s(t) = \frac{C}{tz\sqrt{2\pi}} \exp\left\{\frac{-(\log(t) - K)^2}{2z^2}\right\}$$

where $s(0) = 0$, $C$ and $z$ are estimated in the model fitting, and $K$ is calculated such that the diagnostic sensitivity peaks at the same time as the infectivity curve.

Data from Hellewell et al [12] specifies whether or not the individual exhibited symptoms on the testing date, the outcome of the self-administered nasal RT-PCR test, as well as the Ct value. Hellewell et al [12] used the serology testing to obtain a subset of 27 healthcare workers who seroconverted during the study period. To determine the shape of the diagnostic sensitivity curve of the RT-PCR assay (**Fig. S21**), we maximised the log-likelihood

$$L = L_T + L_P$$

where $L_T$ is the log-likelihood for the time of infection and $L_P$ is the log-likelihood of the result of the RT-PCR test.

The methodology specified by Hellewell et al [12], accounts for the censoring of symptoms between the testing time to determine the time of infection. Given the date at which they were last asymptomatic ($t_i^L$) and the time of in which they are first symptomatic ($t_i^F$) for the $N$ individuals, the log-likelihood for the time of infection is

$$L_T = \sum_{i=1}^{N} \log\left(F(t_i^F - T_i) - F(t_i^L - T_i)\right),$$

where $F$ is the cumulative distribution of the incubation period specified by Ashcroftet al [13]. We specify the upper bound of the time of infection to be the minimum time of the first positive test, the first day of symptoms, and the first day of a non-zero cycle threshold. The lower bound is assumed to be a month prior to the data-driven upper bound.

For the specified diagnostic sensitivity of the test for all individuals and $M_i$ tests for individual $i$, the likelihood for the test result is expressed as

$$L_P = \sum_{i=1}^{N} \sum_{j=1}^{M_i} \log\left(s(x_{i,j})^{T_{i,j}}(1 - s(x_{i,j}))^{1-T_{i,j}}\right),$$

where $T_{i,j}$ indicates the result of test $j$ for individual $i$.

To infer the diagnostic sensitivity curve for the 8.29 day incubation period, we used distribution specified by Qin et. al. [10]. For inferring the diagnostic sensitivity curve under the 11.66 day incubation period, our estimated log-Normal distribution was used (**Supplementary methods: Infectivity profile**).

*Rapid antigen test*

The diagnostic sensitivity of the rapid antigen test is expressed as the product of the diagnostic sensitivity of the RT-PCR and the percent positive agreement of the rapid antigen test with an RT-PCR test. For the rapid antigen test, we utilised percent positive agreement data for BD Veritor, which was provided with relative to the time of symptom onset [14]. We model the percent positive agreement at time $t$ post infection for the rapid antigen test using a linear logistic model

$$\ln\left(\frac{p(t)}{1-p(t)}\right) = \beta_0 + \beta_1(t - t_S).$$

We fit the model to the data by maximising the log-likelihood

$$L = \sum_{i=1}^{N} \log\left(p(t_i)^{S_i}(1 - p(t_i))^{F_i}\right),$$

where $S_i$ is the number of successes at time point $t_i$ and $F_i$ is the number of failures. We specify that $\beta_1 < 0$such that the percent positive agreement declines with respect to time during the optimization. However, there is no data for the percent positive agreement during the incubation period. To infer the percent positive agreement during the incubation period (i.e. $t < t_S$), we extrapolated the logistic regression back to the time of peak infectivity and then constructed a mapping between the relative infectivity, $f(t)$, and the percent positive agreement post-symptom onset.

***Parameterization***

*Travel*

We quantified the extent of travel flow between two countries ($n_{A,B}$ and $n_{B,A}$) using the annual number of arrivals at paid accommodations for 2019. We divided this annual number of 365 to obtain the daily average number of travellers arriving into the country. This level of arrival data was used to maintain a consistent scale of travel flow between the countries, as data pertaining to border arrival was not available for all countries. Thus, the absolute number of daily travellers is underestimated based on 2019 travel but the ratio of travellers between countries should remain relatively constant.

Within the calculation of daily new within-country imminent infections for a specified travel quarantine, we require the number of travellers who are abroad, which we estimate as

$$n_A = m_{AB} \cdot d_{AB} \cdot N_{T,A}$$

where $m_{AB}$ is the fraction of the total population travelling per day and $N_{T,A}$ is the population size of country A such that $n_{A,B} = m_{AB} \cdot N_{T,A}$. The average number of days a traveller from country A spends in country B, $d_{AB}$ , is informed by country specific travel data (**Table S1**). This duration of stay is strictly used to quantify the accumulated number of travellers abroad under the assumption of long-term stay and does not dictate the extent of transmission. Without the duration of stay, one would be left assuming an indefinitely long travel duration, a condition for which approximating the number of travellers visiting countries becomes infeasible. Since travel is assumed to be constant and in steady state, the daily number of country-A residents returning for a long-term visit to country B is equal to the daily number of country-A residents leaving for a long-term visit to country B.

The number of non-traveling and non-quarantined country-A residents for a specified quarantine $q_A$ is

$$N_A = N_{T,A}(1 - m_{AB} \cdot (d_{AB} + \alpha_Q q_A)).$$

We consider only the quarantine duration of the destination country ($q_A$), as the choice of quarantine is independent of the paired countries strategy. Thus, specifying no quarantine for the origin country provides a conservative estimate of the duration of quarantine specified by the destination country.

The data was obtained from 21 different sources, which primarily consists of data from TourMIS and publicly available governmental statistical databases (**Table S1**). For Cyprus, Estonia and France, we had to utilise data from 2018 as data for 2019 was not available. Country specific duration of stay was not available for Denmark, Norway, and Spain. The duration of stay for these countries was the global average duration of stay (i.e. the average for any given traveller to the country). For the duration of stay in Norway, the duration of stay for January 2021 was used as 2018 to 2020 data was not available for this measure. There was no annual number of arrivals at paid accommodations available for Spain, and Denmark. To approximate the number of arrivals at paid accommodations for Spain, we scaled the number of arrivals at the border by the percentage of travellers staying in hotels compared to all venue types. For Denmark, the number of arrivals was determined by dividing the number of bed nights by the duration of stay.

As an alternative data source to inform travel flow between countries, we used annual air passenger transport between countries.[15] This data accounts for travellers whose journey ends at an airport within the reporting country (i.e. destination country), but does not provide indication as to which country the individual resides (i.e. whether from origin country or from destination country and returning to home). Consequently, the asymmetry of travel may not be comprehensively captured with this dataset.

*Vaccination and prevalence*

We utilise age-specific vaccination coverage for each country [16–19] and the age demographics within Europe. [20,21] Those recovered or actively infected were assumed to vaccinate at equal frequency to those never infected. To account for the time to attain vaccine-elicited immunity, the level of vaccine acquired immunity is based on the vaccination coverage in the age class reported two-weeks prior. For example, the level of vaccine acquired immunity for November 21 is dependent on the number of people partially and fully vaccinated as of November 7. Thus, the country specific vaccination coverage is based on the coverage reported for the week containing November 7, 2021, [22] where we determine those that have received only one dose and those who have received two doses.

We utilise age specific vaccine effectiveness in the reduction of documented infection ($\varepsilon$), stratified by dose. [23] To infer the level of natural immunity in the population, we assume that natural immunity is the cumulative number of infections in the country and discount the active infections capable of transmitting and not isolated. With the incidence data being at the daily time scale, we round the incubation period and duration of disease to the closest integer, denoted$\lfloor t \rceil$. Thus, our approximation of the level of natural immunity in the population is

$$r = \left(I_C - \sum_{a=1}^{N} w_a \left[\alpha_I \cdot (1 - p_{A,a}) \cdot \overline{I_T}(\lfloor t_S \rceil) + (1 - \alpha_I) \cdot (1 - p_{A,a}) \cdot \overline{I_T}(\lfloor t_E \rceil) + p_{A,a} \cdot \overline{I_T}(\lfloor t_E \rceil)\right]\right)/N_T,$$

where $I_C$ is the cumulative infections to date, $w_a$ is the proportion of the population in age class $a$, and that $\overline{I_T}(t)$ is the cumulative number of infections over the past $t$ days in the whole population (e.g. $\overline{I_T}(\lfloor 5.72 \rceil)$ is the cumulative infections over the past six days; i.e. infections from November 16 upto and including November 21). The term $\left[\alpha_I \cdot (1 - p_{A,a}) \cdot \overline{I_T}(\lfloor t_S \rceil) + (1 - \alpha_I) \cdot (1 - p_{A,a}) \cdot \overline{I_T}(\lfloor t_E \rceil) + p_{A,a} \cdot \overline{I_T}(\lfloor t_E \rceil)\right]$ is the number of active infections capable of transmitting and not isolated in age class $a$. Specifying uniform natural immunity across age groups, the cumulative number of infections over the past $t$ days within age class $a$ is

$$\overline{I_a}(t) = \overline{I_T}(t) \cdot w_a \cdot \left(1 - (1 - r) \cdot \left(\varepsilon_{a,1}\nu_{a,1} + \varepsilon_{a,2}\nu_{a,2}\right) - r\right) / \sum_{j=1}^{N} w_j \cdot \left(1 - (1 - r) \cdot \left(\varepsilon_{j,1}\nu_{j,1} + \varepsilon_{j,2}\nu_{j,2}\right) - r\right).$$

Specifying $p_{A,a}$ proportion of infections are asymptomatic in age class $a$, the prevalence of non-isolated active infections capable of transmission in age class $a$ is

$$\rho_a = \left(\alpha_I \cdot (1 - p_{A,a}) \cdot \overline{I_a}(\lfloor t_S \rfloor) + (1 - \alpha_I) \cdot (1 - p_{A,a}) \cdot \overline{I_a}(\lfloor t_E \rfloor) + p_{A,a} \cdot \overline{I_a}(\lfloor t_E \rfloor)\right)/(w_a \cdot N_T).$$

Therefore, the proportion of age class $a$ who are immune to infection is specified as

$$\varphi_a = \left(1 - r - \rho_a\right) \cdot \left(\varepsilon_{a,1}\nu_{a,1} + \varepsilon_{a,2}\nu_{a,2}\right) + r + \rho_a,$$

where the level of vaccine acquired immunity in age class $a$ being $\left(1 - r - \rho_a\right) \cdot \left(\varepsilon_{a,1}\nu_{a,1} + \varepsilon_{a,2}\nu_{a,2}\right)$.

***Generalised approach using EU traffic-light system***

For this generalised analysis, we considered four categories of countries based on the European Union traffic-light system that classifies travel-risk using the number of new cases in the past 14 days. We specifically examine the scenarios of 25 (Green status), 150 (Amber status), 500 (Red status) and 1,000 (Dark Red) bi-weekly case-loads per 100,1000 residents. Population size and level of naturally acquired immunity under each category was considered the same and was informed by averaging over all 26 European countries (**Table S6**). The age and dose specific vaccination coverage is informed by that observed within Europe.[24] Each category was assumed to have the same age demographics as Europe[25]. The extent of travel was assumed to be symmetric, with an equal duration of stay that was determined from the average of all origin-destination country pairs (**Table S6**). The number of travellers each day was determined by calculating the product of the average proportion of country travelling and average population size (**Table S6)**.

Daily incidence per capita and prevalence differed for each category. Daily incidence per capita was calculated by averaging over the two weeks of daily cases from the EU traffic-light stratification (e.g., $c$ =(25 cases / 100,000 people) / 14 days for a Green status country). Considering duration of infection was specified to be $t_E$ days and assuming a constant daily incidence, the estimated prevalence of non-isolated infections was calculated as

$$c(t_S(1 - p_A)\alpha_L + t_E(1 - p_A)(1 - \alpha_L) + t_E p_A).$$

***Sensitivity analysis***

To identify the parameters that have the largest one-way influence on the estimated sufficient quarantine durations of travel quarantine, we iteratively altered the value of each parameter by one standard deviation towards the median of the 26 countries while retaining all other parameters fixed. For the age-based parameters—prevalence, vaccine-acquired immunity, natural immunity, and age-demographics—the age-specific values were perturbed by one standard deviation towards the median calculated for the age class. To eliminate the division by zero, the level of immunity was set to 99.9% when the immunity in the age class was 100% or larger after the perturbation. to prevent the population size from adopting negative values due to the perturbation, a $\log_{10}$ transformation of scale was first conducted on the population size parameter.

***Implementation***

We used MATLAB R2019b for the computation and analysis of sufficient travel quarantine durations. When evaluating the inequality for imminent infections, we specified an upper bound error tolerance of $5 \times 10^{-19}$ to avoid the effects of floating-point precision error in the calculation.

### Supplementary Figures

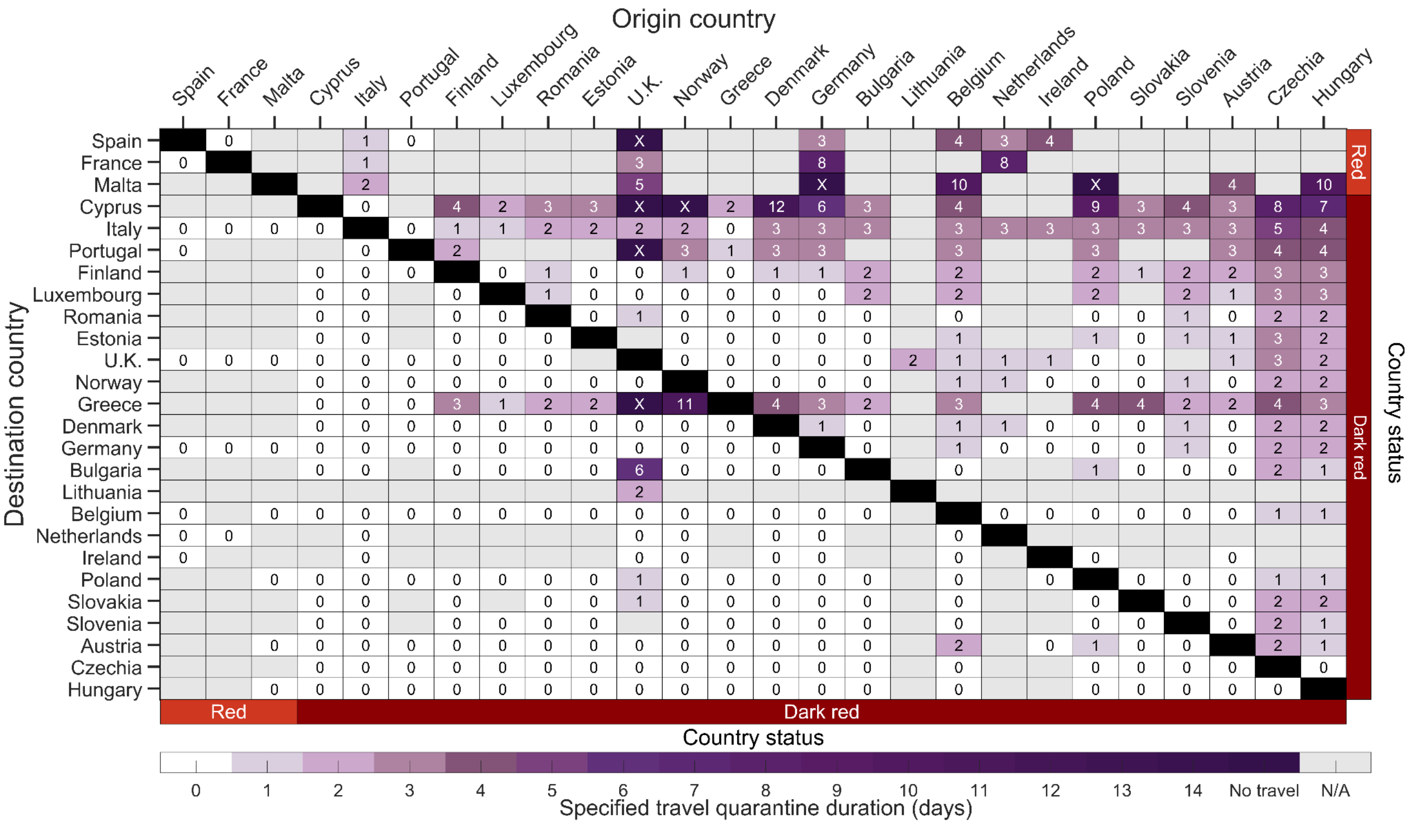


**Figure S1.** The estimated minimum duration of travel quarantine with an RT-PCR test on exit from quarantine for specified origin-destination country pairs that reduces within-country imminent infections to be equivalent to border closure. Specifying an incubation period of 5.72 days, age-dependent vaccine effectiveness and proportion of asymptomatic infections, as well as country-specific demographics, incidence, prevalence of non-isolated infections, vaccine coverage, seroprevalence and travel flow, we determine the minimum duration of travel quarantine with an RT-PCR test on exit from quarantine (colour gradient) that should be stated by the destination country for individuals arriving from the origin country. We consider travel quarantine durations of zero-days (white) to no travel (dark purple, i.e., specified travel quarantine can exceed 14 days). Within-country travel quarantine is not evaluated in the analysis (black). Travel flow data was not available for all country pairs (grey). The countries are ranked based on their estimated incidence per 100,000 over the last two weeks (November 8 to November 21), and stratified based on the European Union country classification system: Green, < 25 cases per 100,000; Amber, 25 to 150 cases per 100,000; Red, 150 to 500 cases per 100,000; and Dark Red, > 500 cases per 100,000. For travel quarantine durations of one day or longer, the RT-PCR test was conducted 24-h before exit from quarantine. For a zero-day travel quarantine, the RT-PCR test was conducted 24-h before travel.

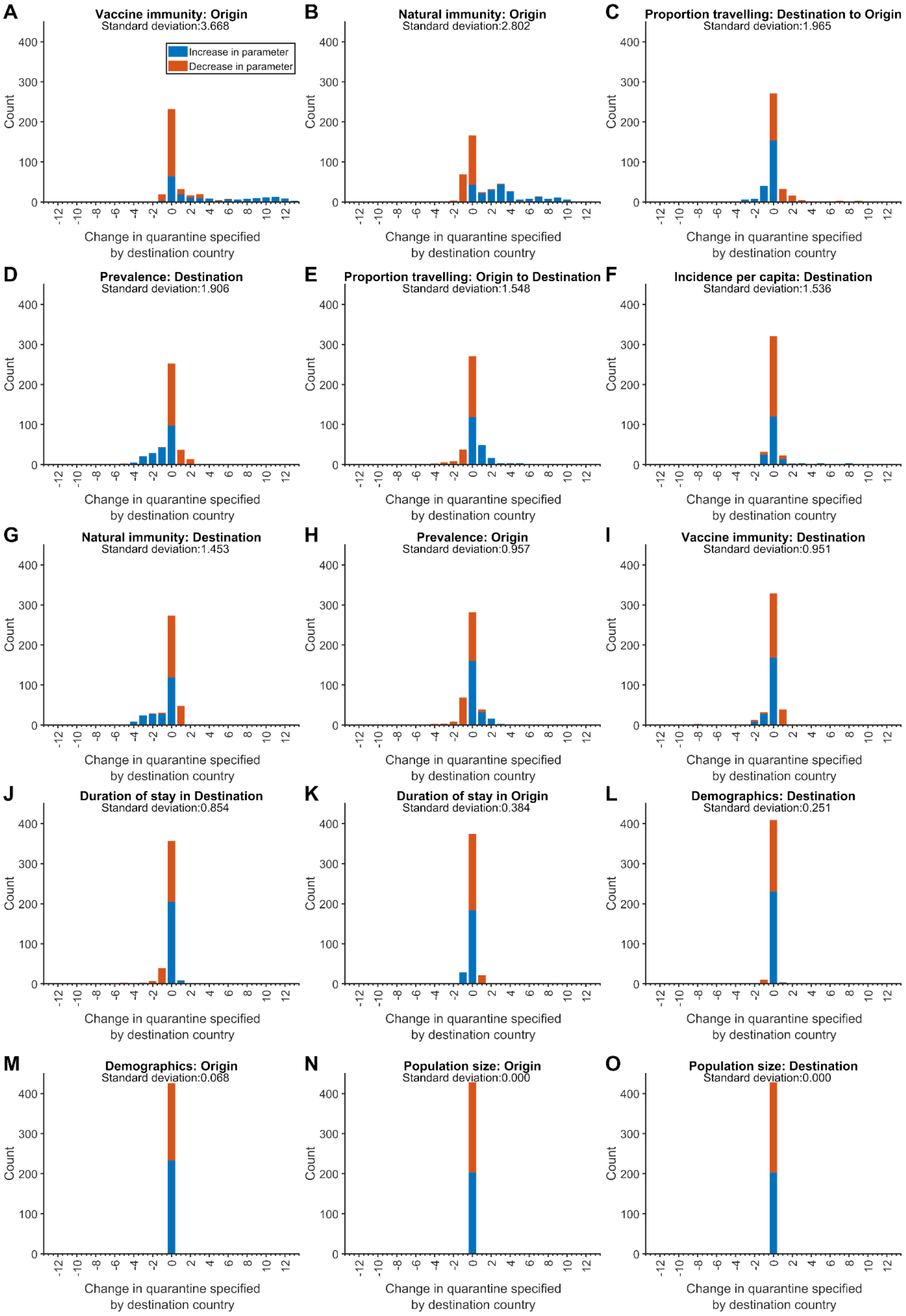
A
Vaccine immunity: Origin
Standard deviation:3.668
Increase in parameter
Decrease in parameter
B
Natural immunity: Origin
Standard deviation:2.802
C
Proportion travelling: Destination to Origin
Standard deviation:1.965
D
Prevalence: Destination
Standard deviation:1.906
E
Proportion travelling: Origin to Destination
Standard deviation:1.548
F
Incidence per capita: Destination
Standard deviation:1.536
G
Natural immunity: Destination
Standard deviation:1.453
H
Prevalence: Origin
Standard deviation:0.957
I
Vaccine immunity: Destination
Standard deviation:0.951
J
Duration of stay in Destination
Standard deviation:0.854
K
Duration of stay in Origin
Standard deviation:0.384
L
Demographics: Destination
Standard deviation:0.251
M
Demographics: Origin
Standard deviation:0.068
N
Population size: Origin
Standard deviation:0.000
O
Population size: Destination
Standard deviation:0.000
Count
Change in quarantine specified by destination country

**Figure S2.** Sensitivity of the minimum sufficient travel quarantine duration to changes in parameters for an RT-PCR test on exit from quarantine. Specifying an incubation period of 5.72 days, age-dependent vaccine effectiveness and proportion of asymptomatic infections, as well as country-specific demographics, incidence, prevalence of non-isolated infections, vaccine coverage, seroprevalence, and travel flow, we determine the change in the minimum sufficient duration of travel quarantine with an RT-PCR test on exit from quarantine when **A**) prevalence in the destination country, **B**) proportion of the population travelling from the destination country to the origin, **C**) proportion of the population travelling from the origin country to the destination, **D**) prevalence in the origin country, **E**) daily incidence per capita in the destination country, **F**) duration of stay in destination (used in determining population abroad), **G**) natural immunity in origin, **H**) duration of stay in origin (used in determining population abroad), **I**) natural immunity in the destination country, **J**) vaccine immunity in the destination country, **K**) vaccine immunity in the origin country, **L**) age demographics in the destination country, **M**) age demographics in the origin country, **N**) the $\log_{10}$ populations size of the origin country, and **O**) the $\log_{10}$ population size in the destination country is moved one standard deviation towards the median of the 26 countries while other parameters remained fixed. We included retention of travel ban in our counts of a zero change in travel quarantine. We excluded from our counts the cases where either a travel ban is lifted or added as a result of the perturbation. Ban of travel was lifted in less than 1.5% of the scenarios for prevalence in the destination, proportion of the population travelling from the origin country to the destination, proportion of the population travelling from the origin country to the destination, daily incidence per capita in the destination. A switch from implementing quarantine to mandating ban of travel occurred in less than 7% of the scenarios for proportion of the population travelling from the origin country to the destination, proportion of the population travelling from the origin country to the destination, daily incidence per capita in the destination, duration of stay in destination, natural immunity in origin, and vaccine immunity in destination. Direction of changes in duration of quarantine are indicated by colour (increased, blue; decreased, orange). Panels are ordered by decreasing standard deviation in the change of quarantine duration (top left). For travel quarantine durations of one day or longer, the RT-PCR test was conducted 24 h before exit from quarantine. For a zero-day travel quarantine, the RT-PCR test was conducted 24-h before travel.

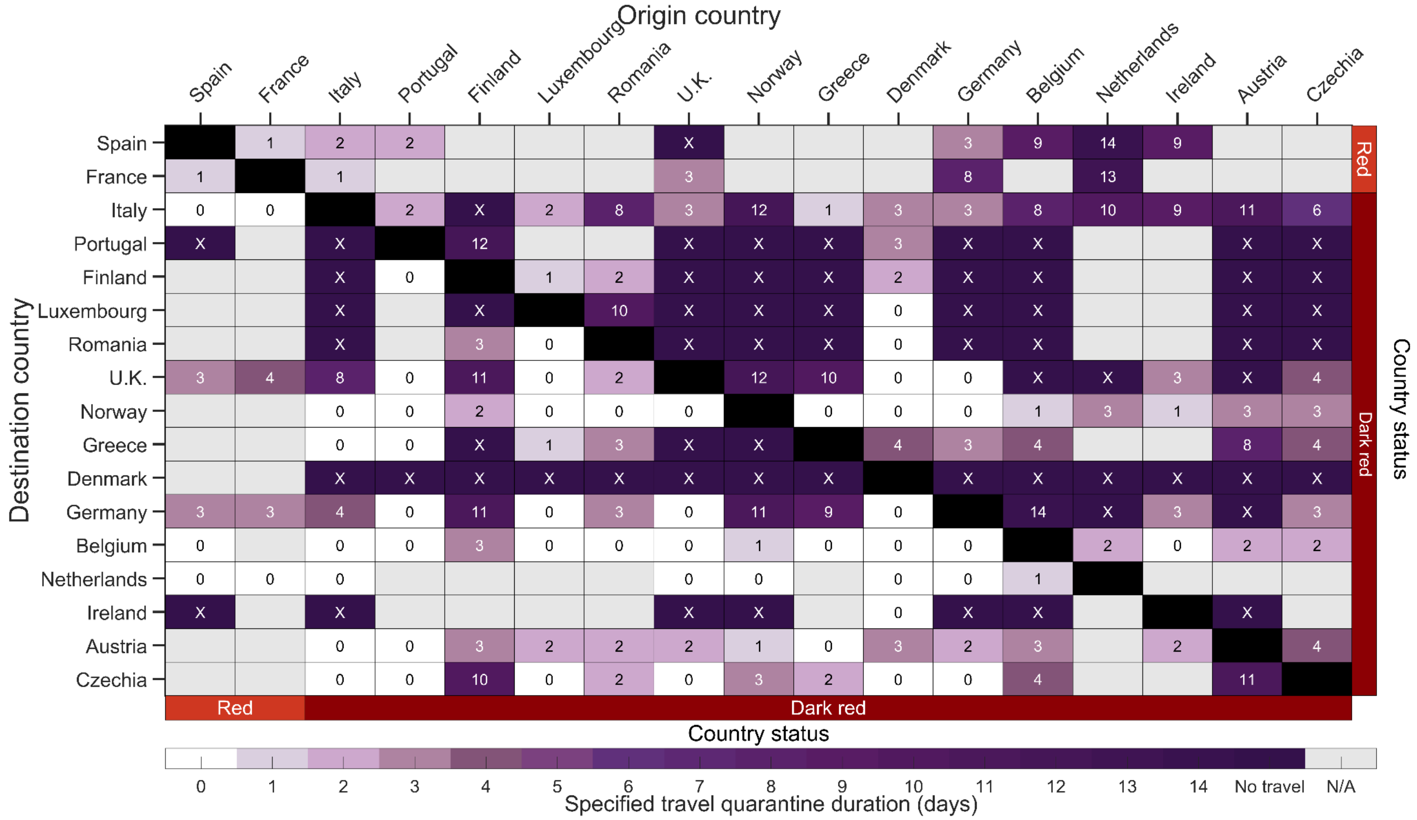


**Figure S3.** The estimated minimum duration of travel quarantine, with an RT-PCR test on exit from quarantine, for specified origin-destination country pairs that reduces within-country imminent infections to be equivalent to border closure when considering variants of concern. Specifying an incubation period of 5.72 days, age-dependent vaccine effectiveness and proportion of asymptomatic infections, as well as country-specific demographics, incidence, prevalence of non-isolated infections, percentage of variants of concern, vaccine coverage, seroprevalence and travel flow, we determine the minimum duration of travel quarantine (colour gradient) that should be stated by the destination country for individuals arriving from the origin country when considering the transmission of the variants of concern Delta G/478K.V1 and Omicron B.1.1.529+BAand general transmission (i.e. variants except Delta G/478K.V1 and Omicron B.1.1.529+BA). We consider travel quarantine durations of zero-days (white) to no travel (dark purple, i.e., specified quarantine can exceed 14 days). Within- country travel quarantine is not evaluated in the analysis (black). Travel flow data was not available for all country pairs (grey). The countries are ranked based on their estimated incidence per 100,000 over the last two weeks (November 8 to November 21), and stratified based on the European Union country classification system: Green, < 25 cases per 100,000; Amber, 25 to 150 cases per 100,000; Red, 150 to 500 cases per 100,000; and Dark Red, > 500 cases per 100,000. For travel quarantine durations of one day or longer, the RT-PCR test was conducted 24 h before exit from quarantine. For a zero-day travel quarantine, the RT-PCR test was conducted 24 h before travel. Please note that the frequency of sampling in some of the countries presented in this Supplementary Figure may not be sufficient enough to adequately assess the level of circulation of the VOCs in the country.

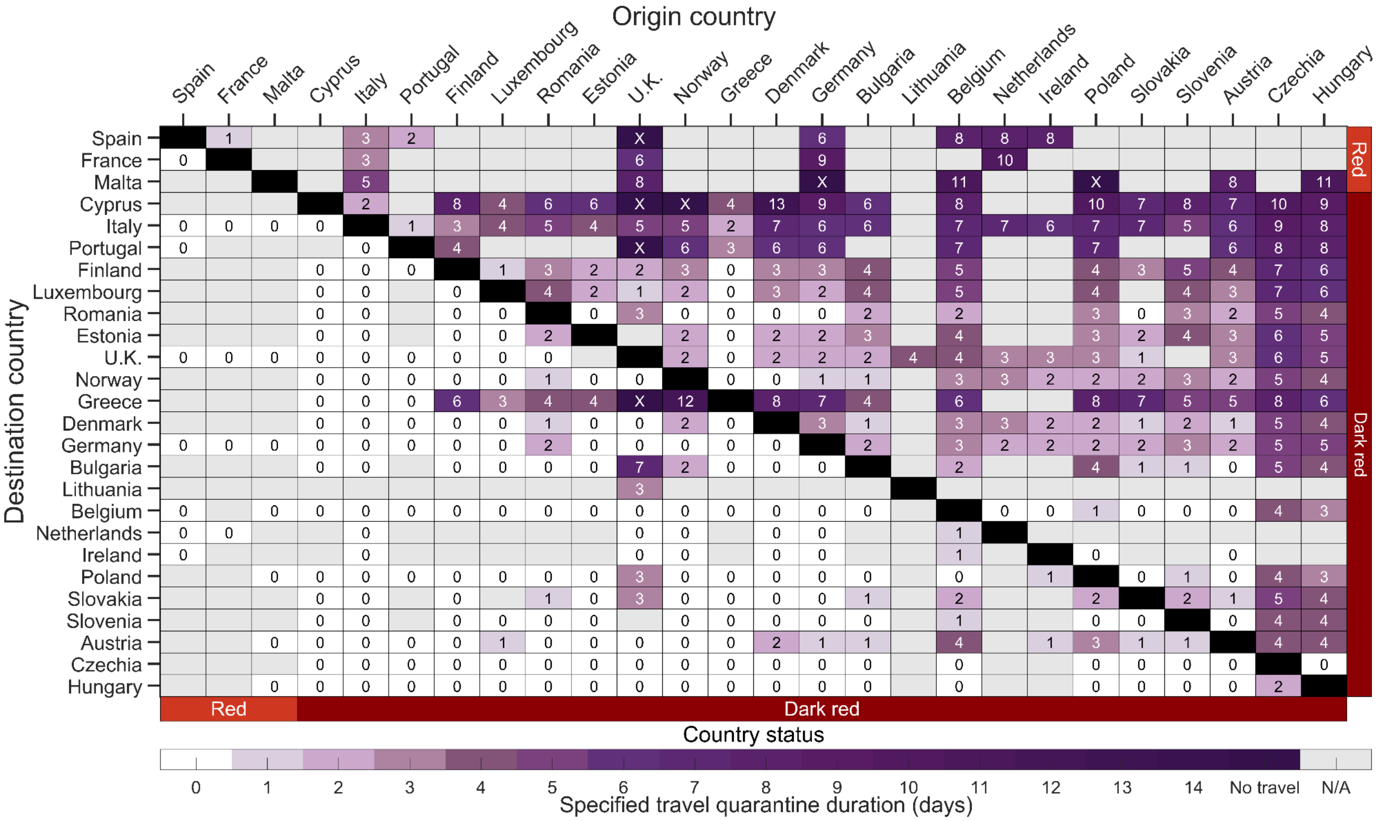


**Figure S4.** The estimated minimum duration of travel quarantine with no testing for specified origin-destination country pairs that reduces within-country imminent infections to be equivalent to border closure for the country-paired analysis. Specifying an incubation period of 5.72 days, age-dependent vaccine effectiveness and proportion of asymptomatic infections, as well as country-specific demographics, incidence, prevalence of non-isolated infections, vaccine coverage, seroprevalence and travel flow, we determine the minimum duration of travel quarantine with no test conducted (colour gradient) that should be stated by the destination country for individuals arriving from the origin country. We consider travel quarantine durations of zero-days (white) to no travel (dark purple, i.e., specified travel quarantine can exceed 14 days). Within-country travel quarantine is not evaluated in the analysis (black). Travel flow data was not available for all country pairs (grey). The countries are ranked based on their estimated incidence per 100,000 over the last two weeks (November 8 to November 21), and stratified based on the European Union country classification system: Green, < 25 cases per 100,000; Amber, 25 to 150 cases per 100,000; Red, 150 to 500 cases per 100,000; and Dark Red, > 500 cases per 100,000.

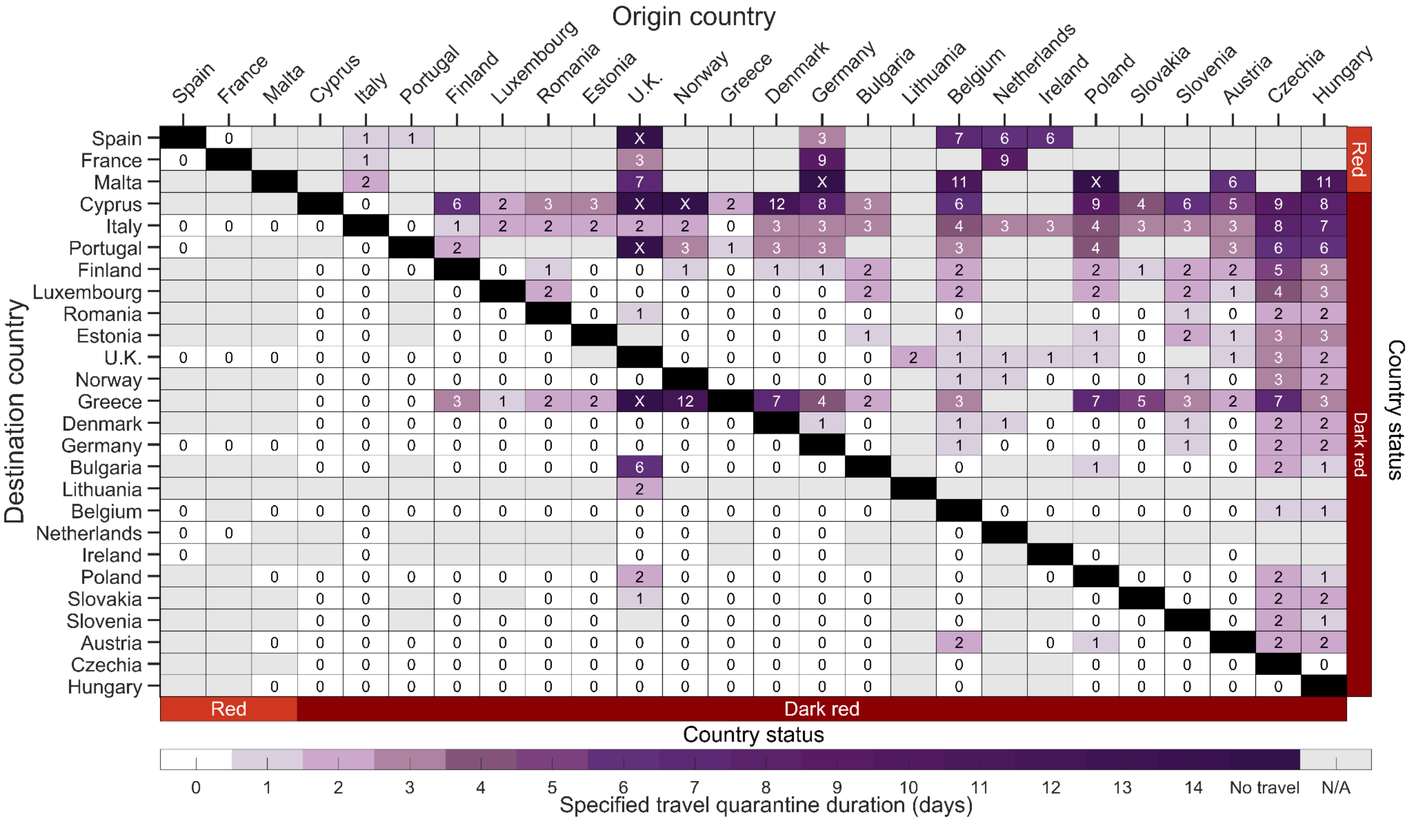


**Figure S5.** The estimated minimum duration of travel quarantine with a rapid antigen test on exit from quarantine for specified origin-destination country pairs that reduces within-country imminent infections to be equivalent to border closure for the country-paired analysis. Specifying an incubation period of 5.72 days, age-dependent vaccine effectiveness and proportion of asymptomatic infections, as well as country-specific demographics, incidence, prevalence of non-isolated infections, vaccine coverage, seroprevalence and travel flow, we determine the minimum duration of travel quarantine with a rapid antigen test on exit (colour gradient) that should be stated by the destination country for individuals arriving from the origin country. We consider travel quarantine durations of zero-days (white) to no travel (dark purple, i.e., specified travel quarantine can exceed 14 days). Within-country travel quarantine is not evaluated in the analysis (black). Travel flow data was not available for all country pairs (grey). The countries are ranked based on their estimated incidence per 100,000 over the last two weeks (November 8 to November 21), and stratified based on the European Union country classification system: Green, < 25 cases per 100,000; Amber, 25 to 150 cases per 100,000; Red, 150 to 500 cases per 100,000; and Dark Red, > 500 cases per 100,000. We assumed that there was no delay in obtaining the test result from a rapid antigen test.

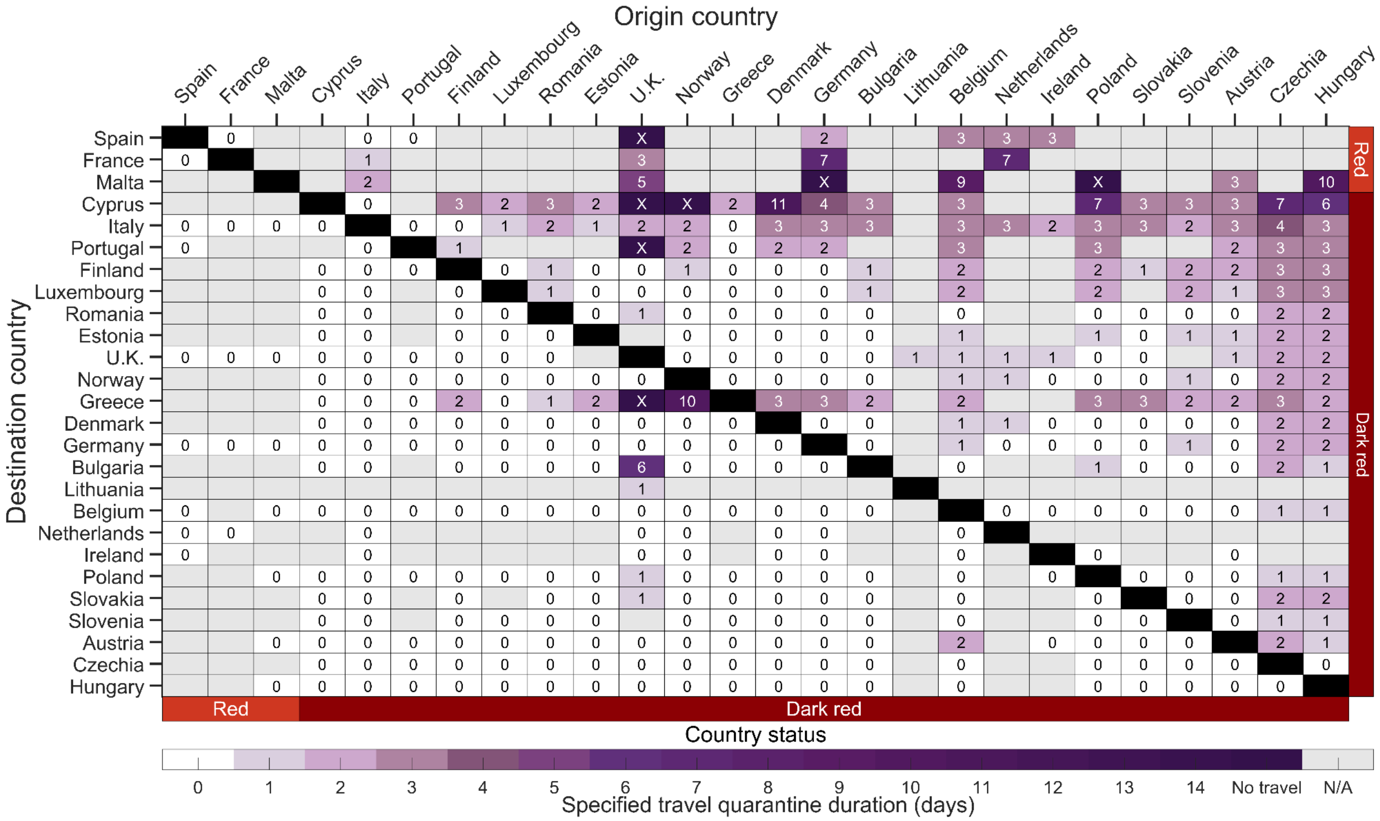


**Figure S6.** The estimated minimum duration of travel quarantine with a rapid antigen test on entry to and exit from quarantine for specified origin-destination country pairs that reduces within-country imminent infections to be equivalent to border closure for the country-paired analysis. Specifying an incubation period of 5.72 days, age-dependent vaccine effectiveness and proportion of asymptomatic infections, as well as country-specific demographics, incidence, prevalence of non-isolated infections, vaccine coverage, seroprevalence and travel flow, we determine the minimum duration of travel quarantine with a rapid antigen test on entry and exit (colour gradient) that should be stated by the destination country for individuals arriving from the origin country. We consider travel quarantine durations of zero-days (white) to no travel (dark purple, i.e., specified travel quarantine can exceed 14 days). Within–country travel quarantine is not evaluated in the analysis (black). Travel flow data was not available for all country pairs (grey). The countries are ranked based on their estimated incidence per 100,000 over the last two weeks (November 8 to November 21), and stratified based on the European Union country classification system: Green, < 25 cases per 100,000; Amber, 25 to 150 cases per 100,000; Red, 150 to 500 cases per 100,000; and Dark Red, > 500 cases per 100,000. There was assumed to be no delay in obtaining the test result from a rapid antigen test.

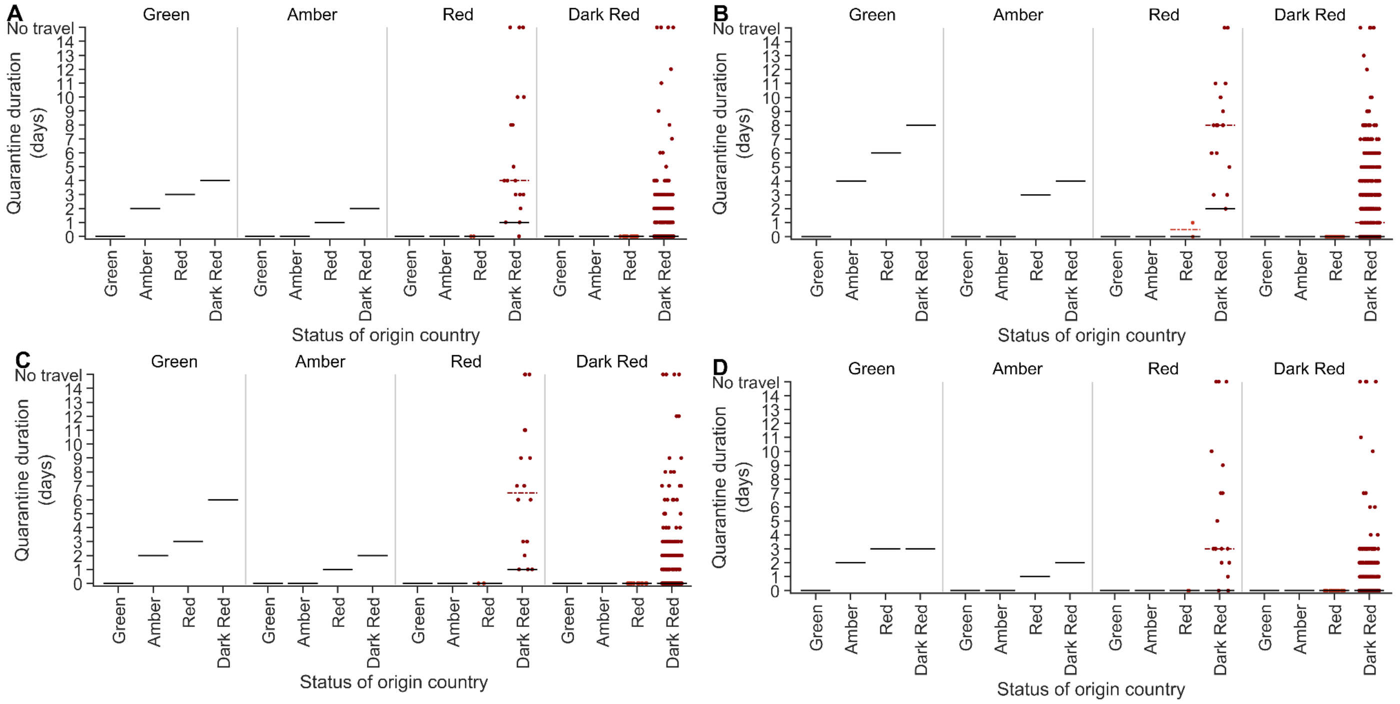


**Figure S7.** Comparison between the country-specific quarantine durations and the durations determined from the EU COVID classification system. For the epidemic situation as of November 21, the estimates for the minimum quarantine duration are stratified by destination country and origin country (*x* axis) for the country-pair analysis (dots; median dashed line) and our tier-based analysis (solid line) for a quarantine with **A**) a RT-PCR test on exit from quarantine, **B**) no testing conducted, **C**) rapid antigen test on exit from quarantine, and **D**) a rapid antigen test on both entry to and exit from quarantine. For travel quarantine durations of one day or longer, the RT-PCR test was conducted 24-h before exit from quarantine and with no delay in obtaining the rapid antigen test. For a zero-day travel quarantine,  the RT-PCR test was conducted 24 h before travel.

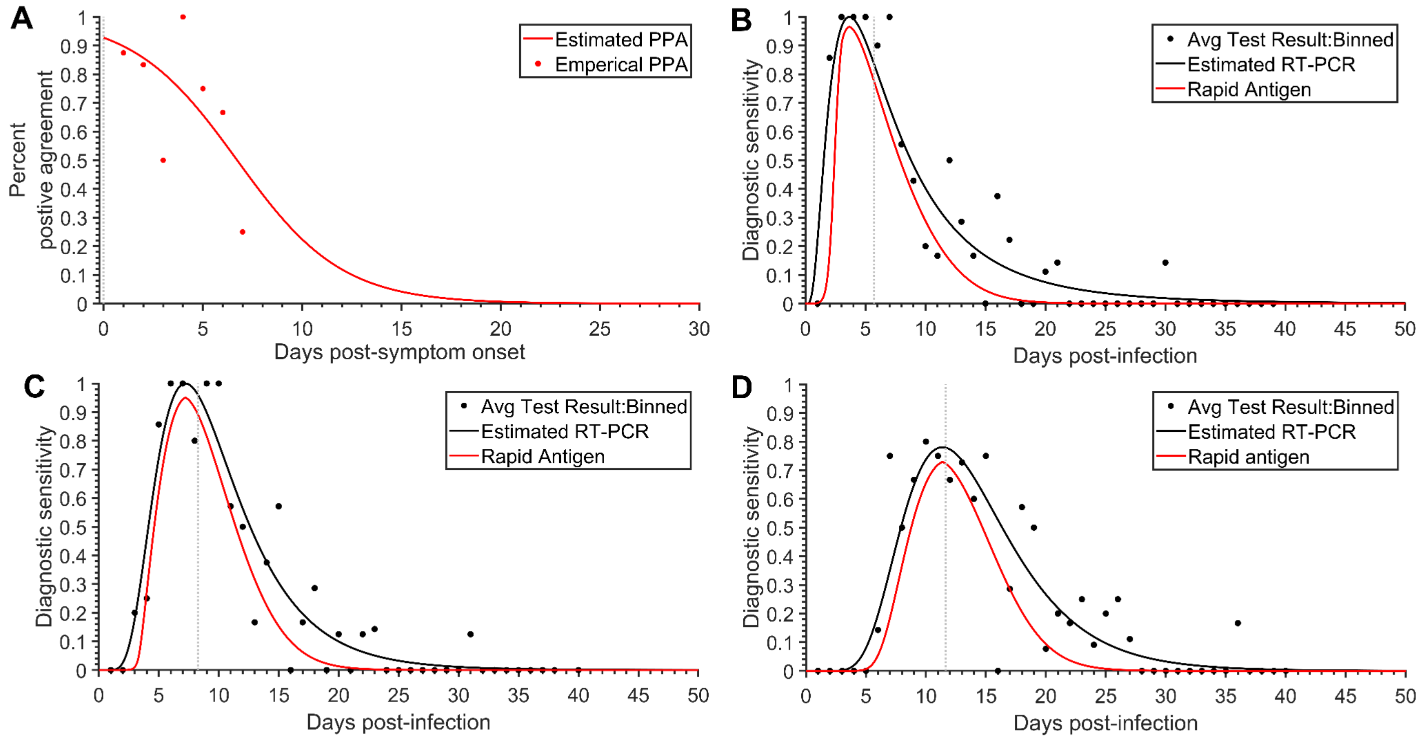


**Figure S8.** The temporal diagnostic sensitivity of a RT-PCR test and rapid antigen test**. A**) the estimated percent positive agreement for a rapid antigen test (red line) that was fit to the percent positive agreement data for the BD Veritor rapid antigen test [14] (red dots). Specifying an incubation period of 5.72 days (vertical grey dashed line) and a latent period of 2.9 days, the **B**) diagnostic sensitivity of an RT-PCR test (represented by a log-Normal probability density function; black line) that was fit to the empirical data from Hellewell et al [26] and corresponding rapid antigen test (red line). [4] Specifying an 8.29 day incubation period and a latent period of 2.9 days, the **C**) diagnostic sensitivity of an RT-PCR test (represented by a log-Normal probability density function; black line) that was fit to the empirical data from Hellewell et al [26] and corresponding rapid antigen test (red line) using the data from Hellewell et al [26]. Specifying an 11.66 day incubation period and a latent period of 2.9 days, the **D**) diagnostic sensitivity of an RT-PCR test (represented by a log-Normal probability density function; black line) that was fit to the empirical data from Hellewell et al [26] and corresponding rapid antigen test (red line) using the data from Hellewell et al [26].

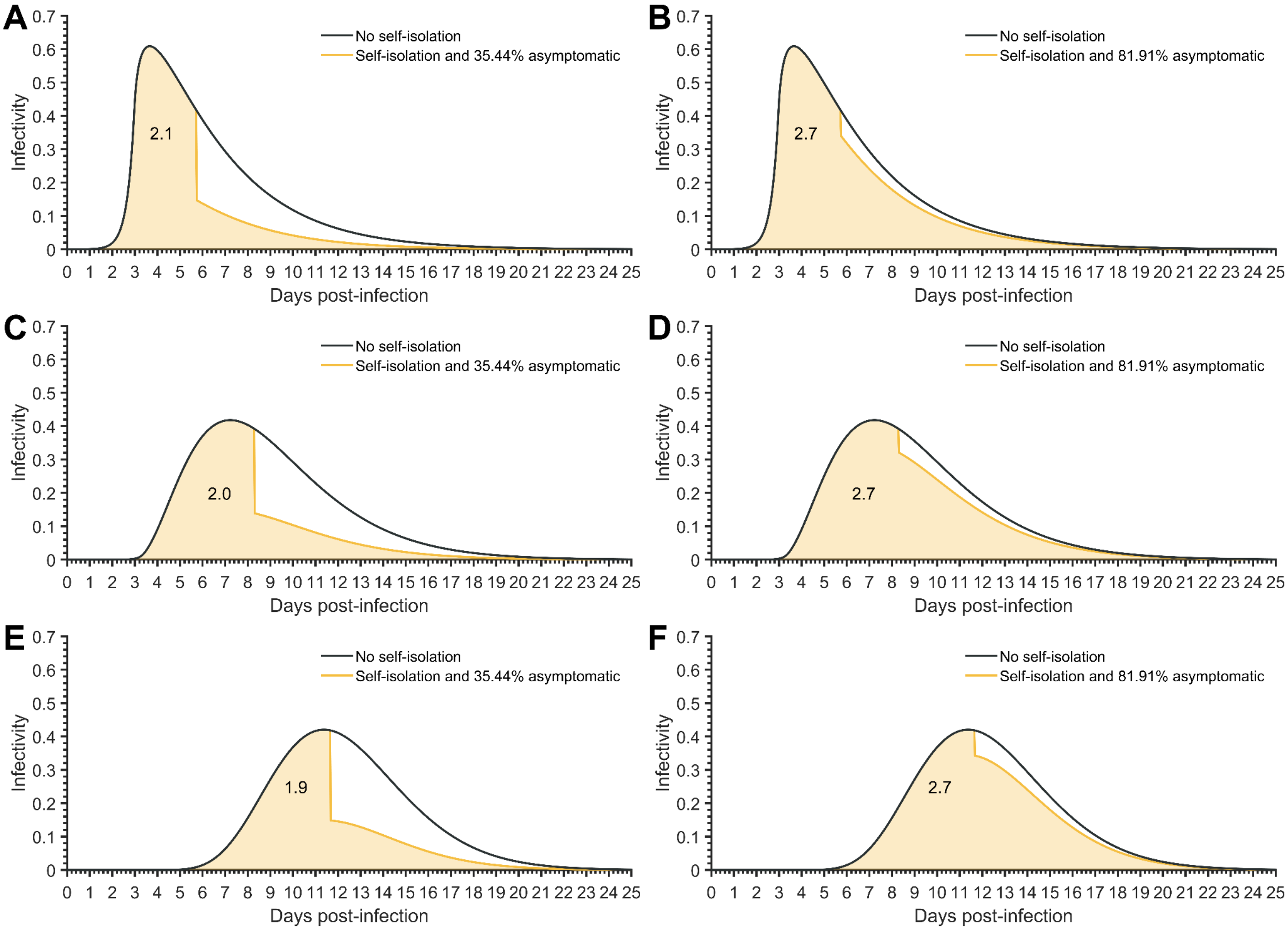


**Figure S9.** The average temporal infectivity curve**.** Specifying a latent period of 2.9 days and a basic reproduction number of three, the average infectivity curve for a known time of infection under no self-isolation upon symptom onset (black) and perfect isolation upon symptom onset (yellow line) for **A**) an incubation period of 5.72 days and 35.44% of infections being asymptomatic (resulting in 2 secondary infections, yellow fill), **B**) an incubation period of 5.72 days and 81.91% of infections being asymptomatic (resulting in 2.7 secondary infections, yellow fill), **C**) an incubation period of 8.29 days and 35.44% of infections being asymptomatic (resulting in 2.1 secondary infections, yellow fill), **D**) an incubation period of 8.29 days and 81.91% of infections being asymptomatic (resulting in 2.7 secondary infections, yellow fill), **E**) an incubation period of 11.66 days and 35.44% of infections being asymptomatic (resulting in 1.9 secondary infections, yellow fill), and **F**) an incubation period of 11.66 days and 81.91% of infections being asymptomatic (resulting in 2.7 secondary infections, yellow fill).

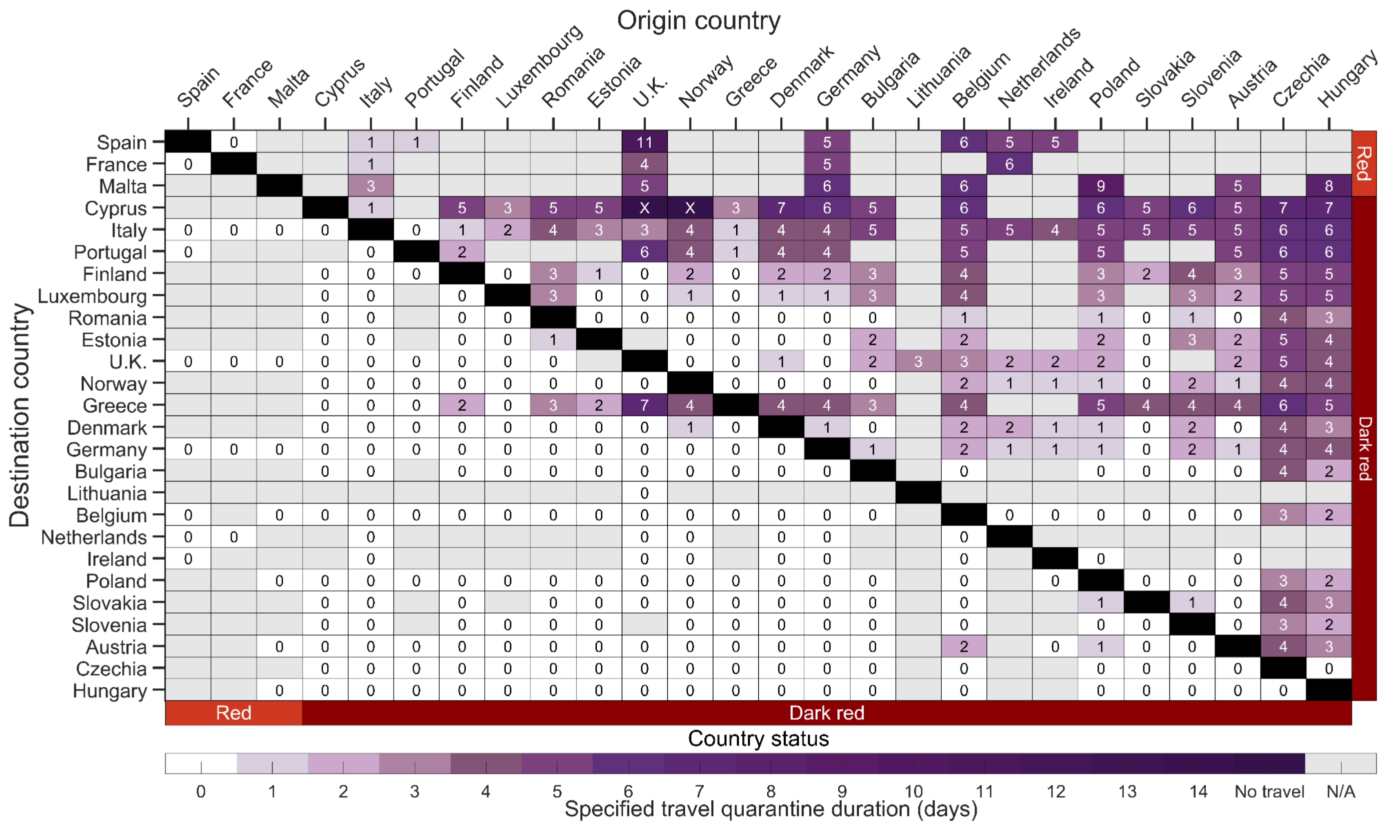


**Figure S10.** The estimated minimum duration of travel quarantine with an RT-PCR test on exit from quarantine for specified origin-destination country pairs that reduces within-country imminent infections to be equivalent to border closure for an incubation period of 8.29 days. Specifying age-dependent vaccine effectiveness and proportion of asymptomatic infections, as well as country-specific demographics, incidence, prevalence of non-isolated infections, vaccine coverage, seroprevalence and travel flow, we determine the minimum duration of travel quarantine with an RT-PCR test on exit from quarantine (colour gradient) that should be stated by the destination country for individuals arriving from the origin country. We consider travel quarantine durations of zero-days (white) to no travel (dark purple, i.e., specified travel quarantine can exceed 14 days). Within-country travel quarantine is not evaluated in the analysis (black). Travel flow data was not available for all country pairs (grey). The countries are ranked based on their estimated incidence per 100,000 over the last two weeks (November 8 to November 21), and stratified based on the European Union country classification system: Green, < 25 cases per 100,000; Amber, 25 to 150 cases per 100,000; Red, 150 to 500 cases per 100,000; and Dark Red, > 500 cases per 100,000. For travel quarantine durations of one day or longer, the RT-PCR test was conducted 24-h before exit from quarantine. For a zero-day travel quarantine, the RT-PCR test was conducted 24-h before travel.

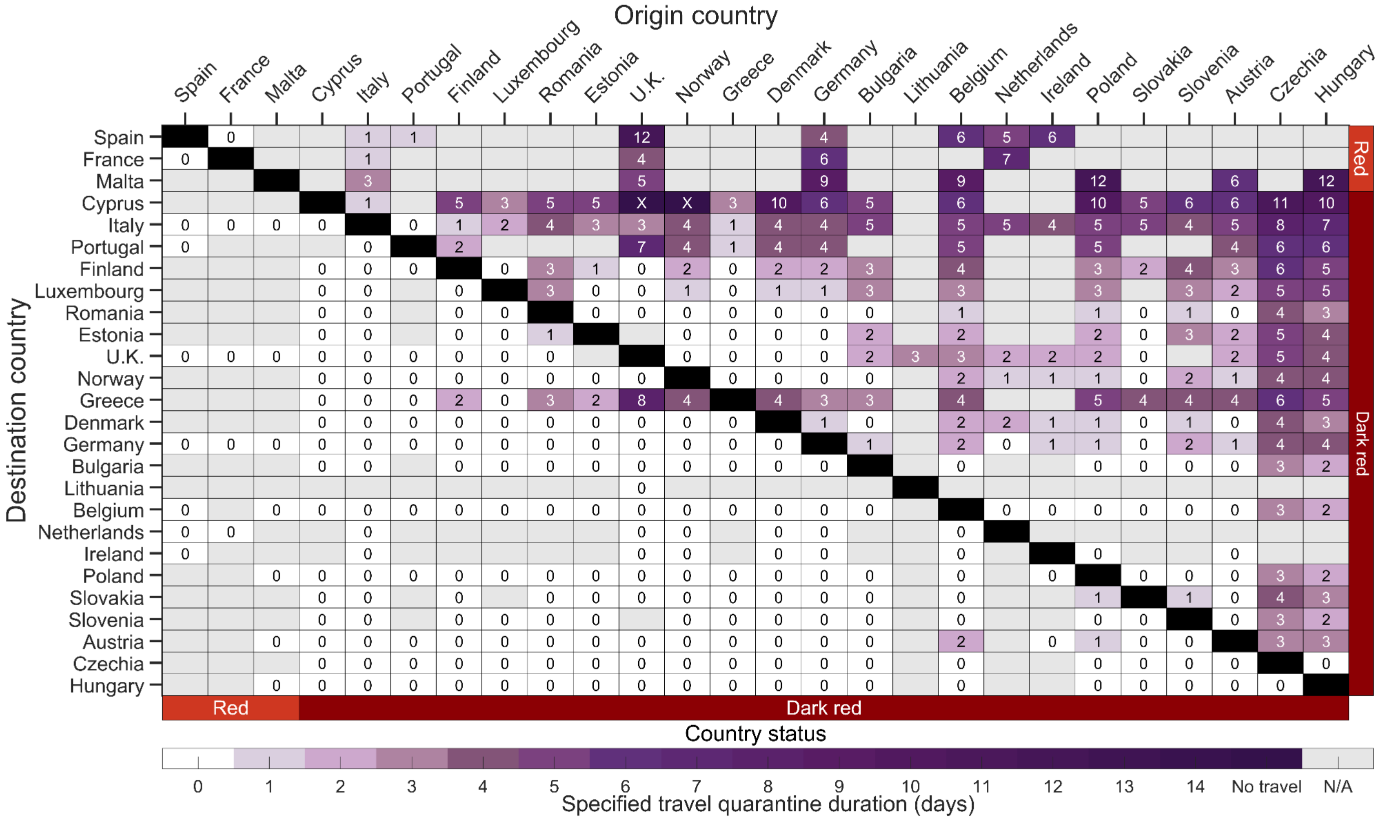


**Figure S11.** The estimated minimum duration of travel quarantine with a rapid antigen test on exit from quarantine for specified origin-destination country pairs that reduces within-country imminent infections to be equivalent to border closure for the country-paired analysis for an incubation period of 8.29 days. Specifying age-dependent vaccine effectiveness and proportion of asymptomatic infections, as well as country-specific demographics, incidence, prevalence of non-isolated infections, vaccine coverage, seroprevalence and travel flow, we determine the minimum duration of travel quarantine with a rapid antigen test on exit (colour gradient) that should be stated by the destination country for individuals arriving from the origin country. We consider travel quarantine durations of zero-days (white) to no travel (dark purple, i.e., specified travel quarantine can exceed 14 days). Within-country travel quarantine is not evaluated in the analysis (black). Travel flow data was not available for all country pairs (grey). The countries are ranked based on their estimated incidence per 100,000 over the last two weeks (November 8 to November 21), and stratified based on the European Union country classification system: Green, < 25 cases per 100,000; Amber, 25 to 150 cases per 100,000; Red, 150 to 500 cases per 100,000; and Dark Red, > 500 cases per 100,000. We assumed that there was no delay in obtaining the test result from a rapid antigen test.

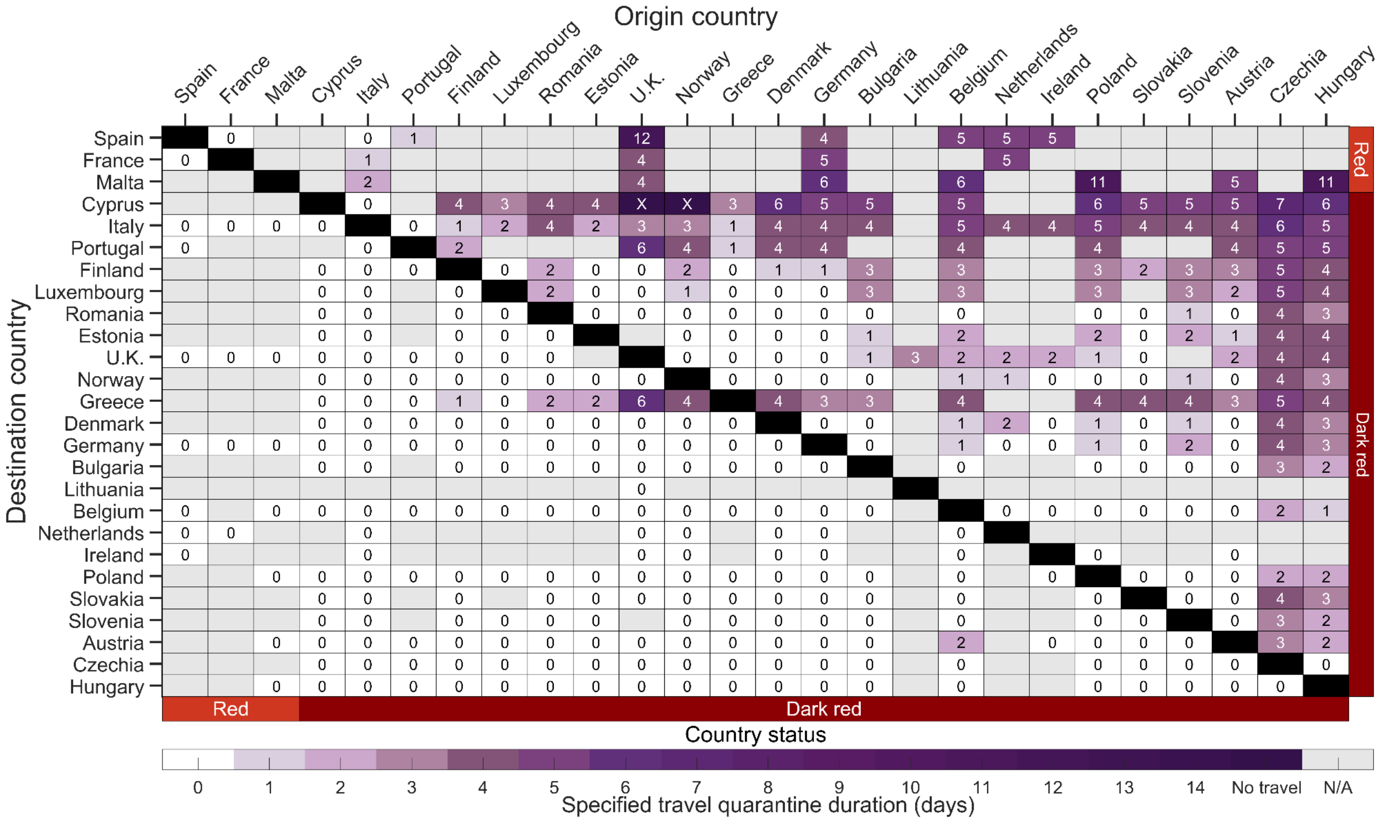


**Figure S12.** The estimated minimum duration of travel quarantine with a rapid antigen test on entry to and exit from quarantine for specified origin-destination country pairs that reduces within-country imminent infections to be equivalent to border closure for the country-paired analysis for an incubation period of 8.29 days. Specifying age-dependent vaccine effectiveness and proportion of asymptomatic infections, as well as country-specific demographics, incidence, prevalence of non-isolated infections, vaccine coverage, seroprevalence and travel flow, we determine the minimum duration of travel quarantine with a rapid antigen test on entry and exit (colour gradient) that should be stated by the destination country for individuals arriving from the origin country. We consider travel quarantine durations of zero-days (white) to no travel (dark purple, i.e., specified travel quarantine can exceed 14 days). Within–country travel quarantine is not evaluated in the analysis (black). Travel flow data was not available for all country pairs (grey). The countries are ranked based on their estimated incidence per 100,000 over the last two weeks (November 8 to November 21), and stratified based on the European Union country classification system: Green, < 25 cases per 100,000; Amber, 25 to 150 cases per 100,000; Red, 150 to 500 cases per 100,000; and Dark Red, > 500 cases per 100,000. There was assumed to be no delay in obtaining the test result from a rapid antigen test.

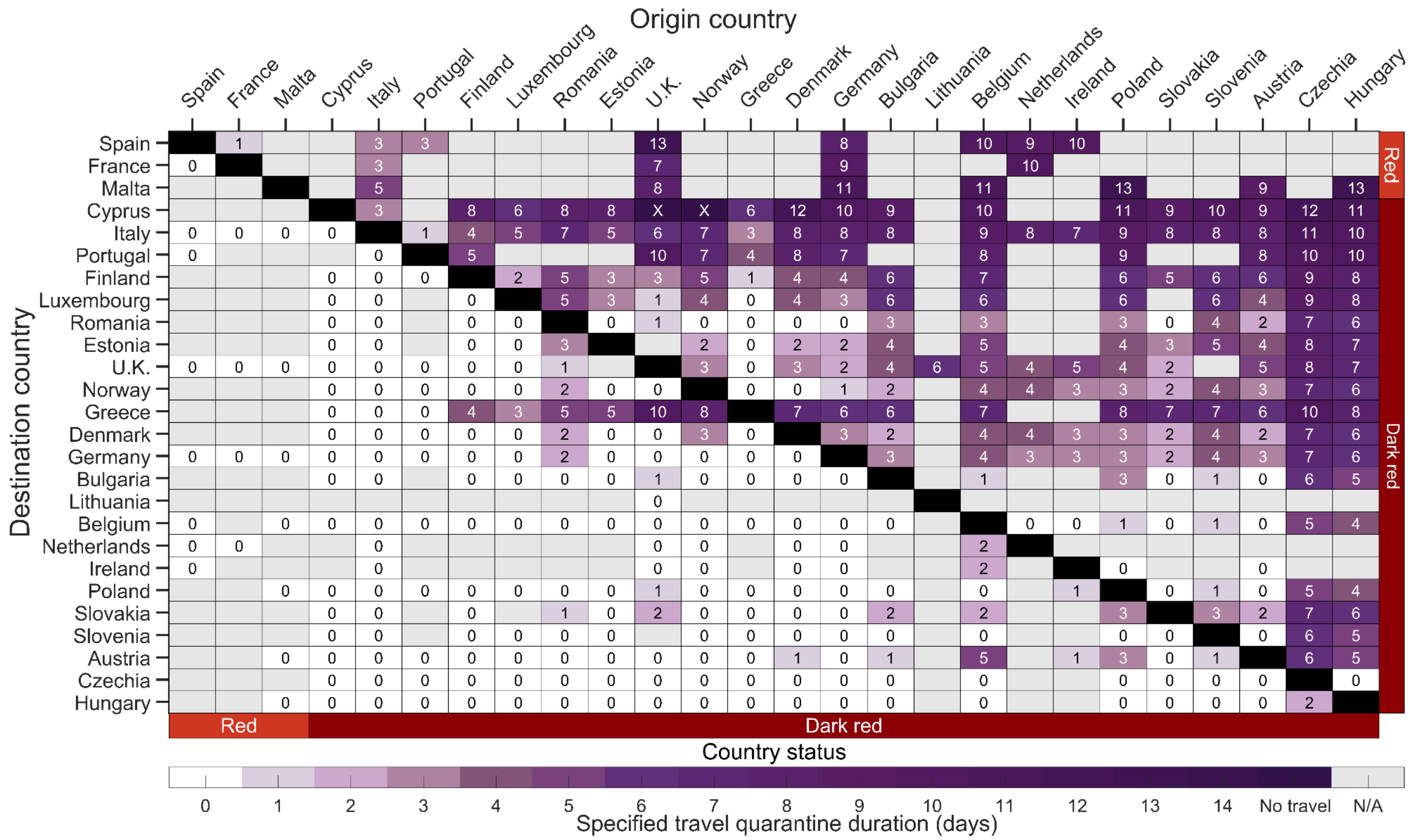


**Figure S13.** The estimated minimum duration of travel quarantine with no testing for specified origin-destination country pairs that reduces within-country imminent infections to be equivalent to border closure for the country-paired analysis for an incubation period of 8.29 days. Specifying age-dependent vaccine effectiveness and proportion of asymptomatic infections, as well as country-specific demographics, incidence, prevalence of non-isolated infections, vaccine coverage, seroprevalence and travel flow, we determine the minimum duration of travel quarantine with no test conducted (colour gradient) that should be stated by the destination country for individuals arriving from the origin country. We consider travel quarantine durations of zero-days (white) to no travel (dark purple, i.e., specified travel quarantine can exceed 14 days). Within-country travel quarantine is not evaluated in the analysis (black). Travel flow data was not available for all country pairs (grey). The countries are ranked based on their estimated incidence per 100,000 over the last two weeks (November 8 to November 21), and stratified based on the European Union country classification system: Green, < 25 cases per 100,000; Amber, 25 to 150 cases per 100,000; Red, 150 to 500 cases per 100,000; and Dark Red, > 500 cases per 100,000.

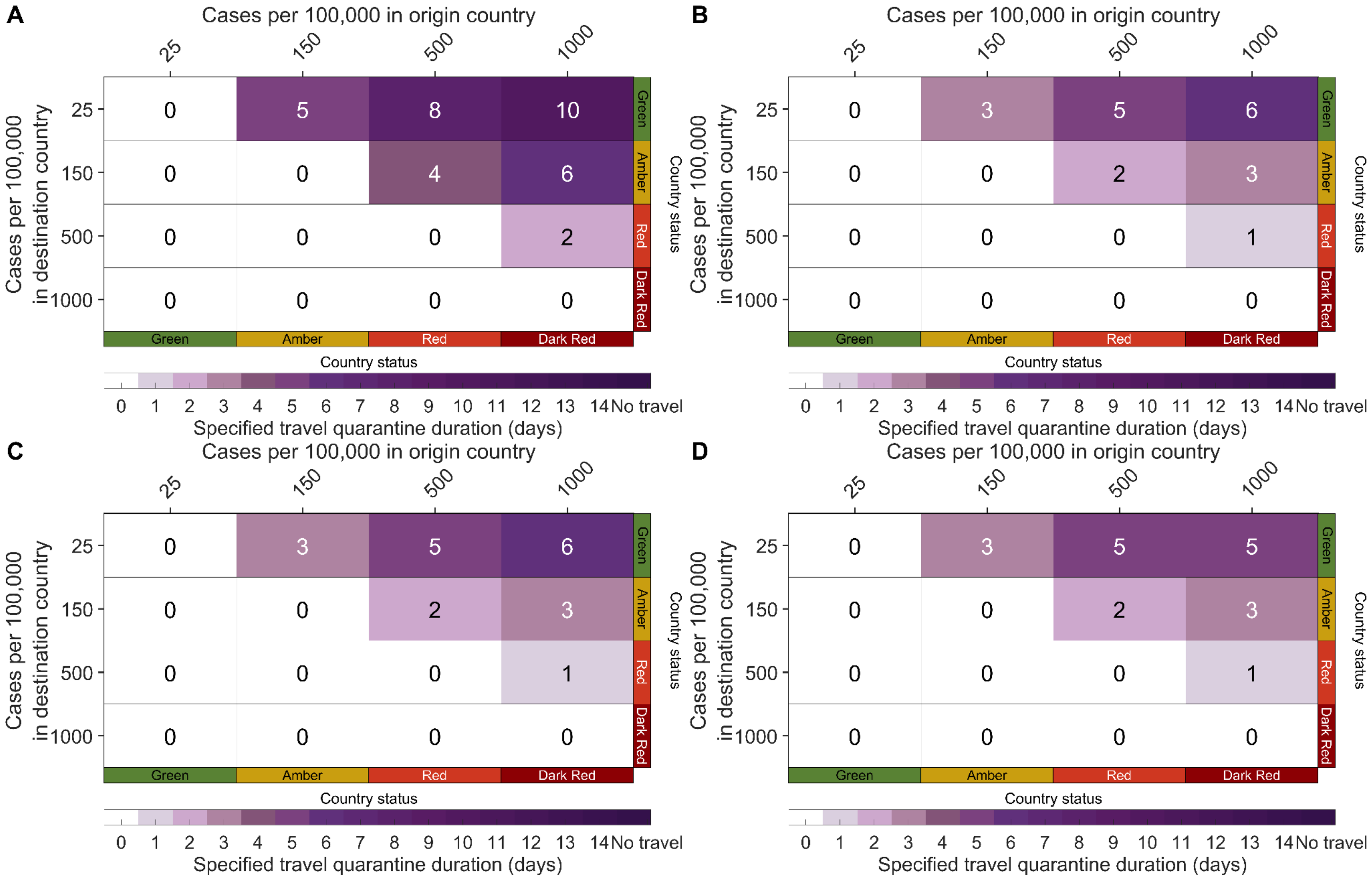


**Figure S14.** The estimated minimum duration of travel quarantine for specified origin-destination country pairs based on the European Union risk states that reduces within-country imminent infections to be equivalent to border closure for an incubation period of 8.29 days**.** Specifying age-dependent vaccine effectiveness and proportion of asymptomatic infections, as well as European demographics, 42% vaccine elicited immunity, 32% natural immunity, we determine the minimum travel duration of quarantine (colour gradient) that should be stated by the destination country for individuals arriving from the origin country. We consider travel quarantine durations of zero days (white) to a travel ban (dark purple, i.e., specified quarantine exceeding 14 days) with **A**) no test, **B**) an RT-PCR test on exit from quarantine, **C**) a rapid antigen test on exit from quarantine, and **D**) a rapid antigen test on both entry to and exit from quarantine. For travel quarantine durations of one day or longer, the RT-PCR test was conducted 24 h before exit from quarantine. and no delay in obtaining the rapid antigen test. For a zero-day travel quarantine, the RT-PCR test was conducted 24 h before travel.

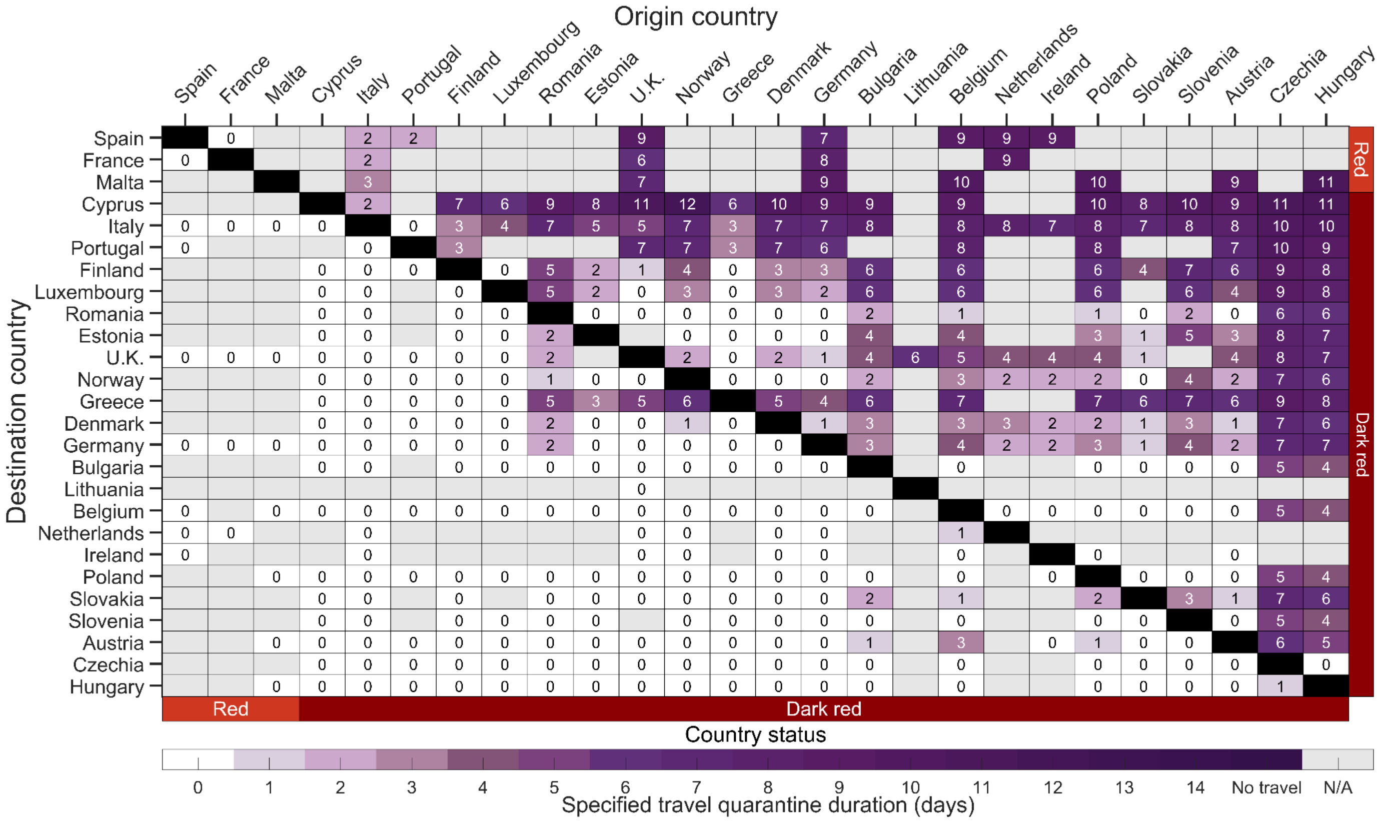


**Figure S15.** The estimated minimum duration of travel quarantine with an RT-PCR test on exit from quarantine for specified origin-destination country pairs that reduces within-country imminent infections to be equivalent to border closure for an incubation period of 11.66 days. Specifying age-dependent vaccine effectiveness and proportion of asymptomatic infections, as well as country-specific demographics, incidence, prevalence of non-isolated infections, vaccine coverage, seroprevalence and travel flow, we determine the minimum duration of travel quarantine with an RT-PCR test on exit from quarantine (colour gradient) that should be stated by the destination country for individuals arriving from the origin country. We consider travel quarantine durations of zero-days (white) to no travel (dark purple, i.e., specified travel quarantine can exceed 14 days). Within-country travel quarantine is not evaluated in the analysis (black). Travel flow data was not available for all country pairs (grey). The countries are ranked based on their estimated incidence per 100,000 over the last two weeks (November 8 to November 21),  and stratified based on the European Union country classification system: Green, < 25 cases per 100,000; Amber, 25 to 150 cases per 100,000; Red, 150 to 500 cases per 100,000; and Dark Red, > 500 cases per 100,000. For travel quarantine durations of one day or longer, the RT-PCR test was conducted 24-h before exit from quarantine. For a zero-day travel quarantine, the RT-PCR test was conducted 24-h before travel.

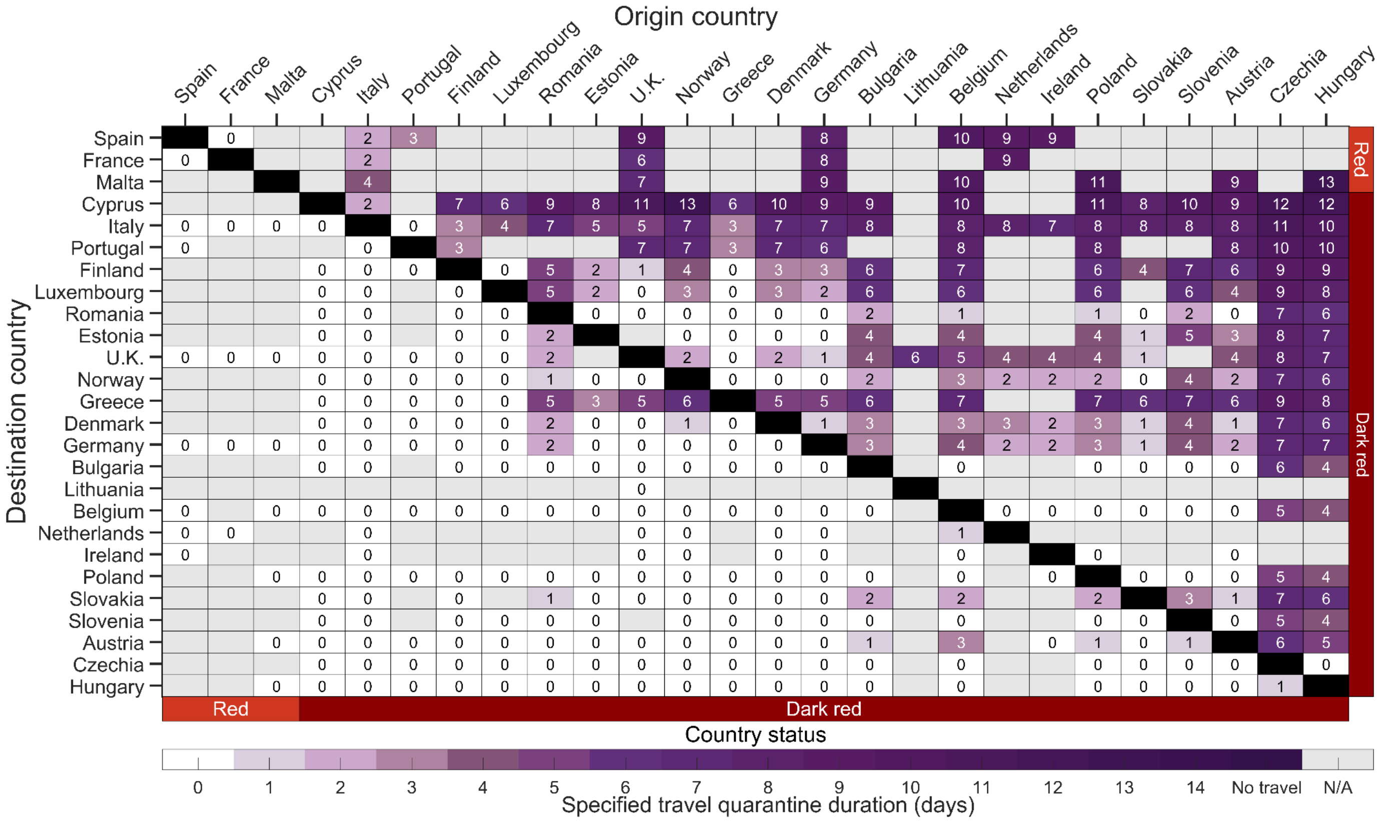

**Figure S16.** The estimated minimum duration of travel quarantine with a rapid antigen test on exit from quarantine for specified origin-destination country pairs that reduces within-country imminent infections to be equivalent to border closure for the country-paired analysis for an incubation period of 11.66 days. Specifying age-dependent vaccine effectiveness and proportion of asymptomatic infections, as well as country-specific demographics, incidence, prevalence of non-isolated infections, vaccine coverage, seroprevalence and travel flow, we determine the minimum duration of travel quarantine with a rapid antigen test on exit (colour gradient) that should be stated by the destination country for individuals arriving from the origin country. We consider travel quarantine durations of zero-days (white) to no travel (dark purple, i.e., specified travel quarantine can exceed 14 days). Within-country travel quarantine is not evaluated in the analysis (black). Travel flow data was not available for all country pairs (grey). The countries are ranked based on their estimated incidence per 100,000 over the last two weeks (November 8 to November 21), and stratified based on the European Union country classification system: Green, < 25 cases per 100,000; Amber, 25 to 150 cases per 100,000; Red, 150 to 500 cases per 100,000; and Dark Red, > 500 cases per 100,000. We assumed that there was no delay in obtaining the test result from a rapid antigen test.

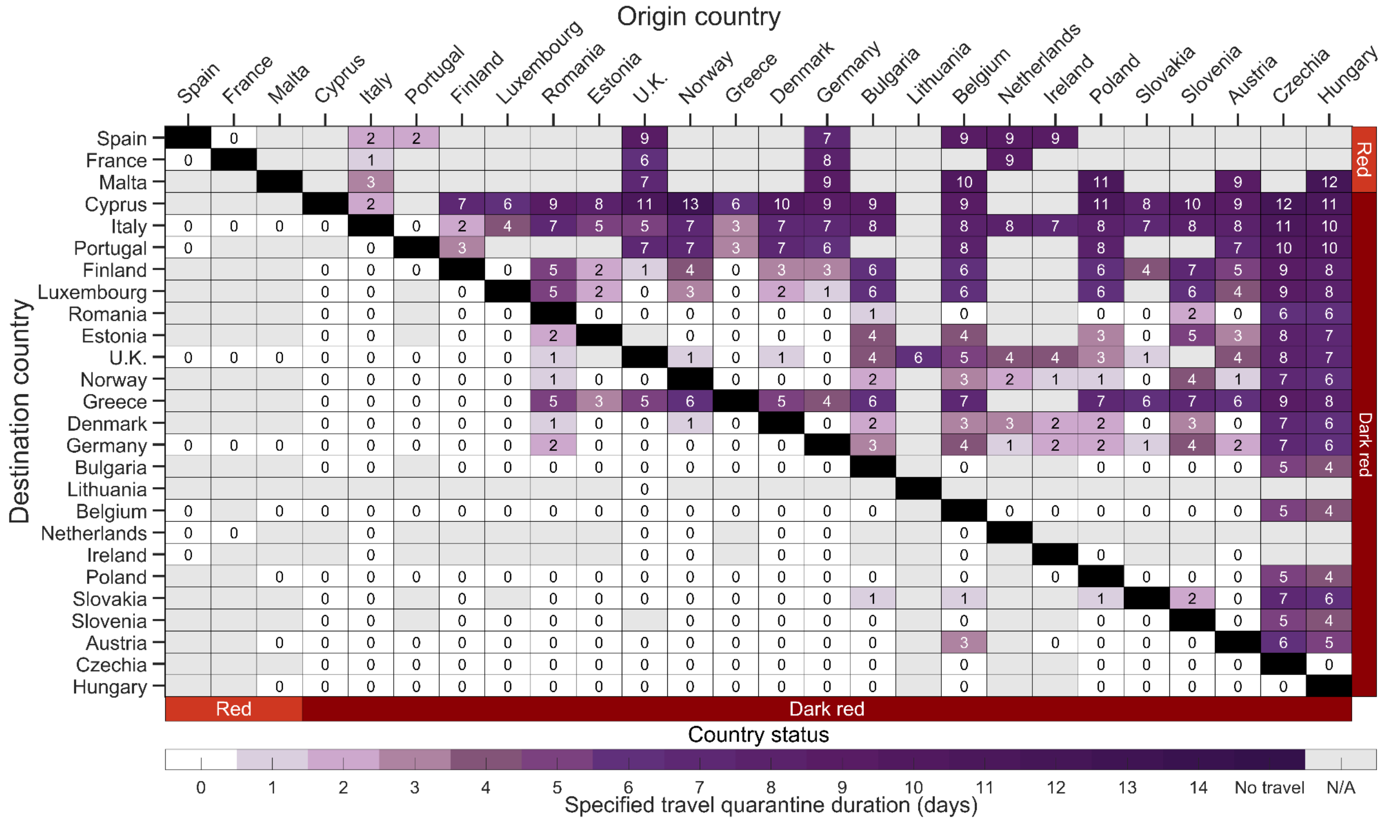

**Figure S17.** The estimated minimum duration of travel quarantine with a rapid antigen test on entry to and exit from quarantine for specified origin-destination country pairs that reduces within-country imminent infections to be equivalent to border closure for the country-paired analysis for an incubation period of 11.66 days. Specifying age-dependent vaccine effectiveness and proportion of asymptomatic infections, as well as country-specific demographics, incidence, prevalence of non-isolated infections, vaccine coverage, seroprevalence and travel flow, we determine the minimum duration of travel quarantine with a rapid antigen test on entry and exit (colour gradient) that should be stated by the destination country for individuals arriving from the origin country. We consider travel quarantine durations of zero-days (white) to no travel (dark purple, i.e., specified travel quarantine can exceed 14 days). Within–country travel quarantine is not evaluated in the analysis (black). Travel flow data was not available for all country pairs (grey). The countries are ranked based on their estimated incidence per 100,000 over the last two weeks (November 8 to November 21), and stratified based on the European Union country classification system: Green, < 25 cases per 100,000; Amber, 25 to 150 cases per 100,000; Red, 150 to 500 cases per 100,000; and Dark Red, > 500 cases per 100,000. There was assumed to be no delay in obtaining the test result from a rapid antigen test.

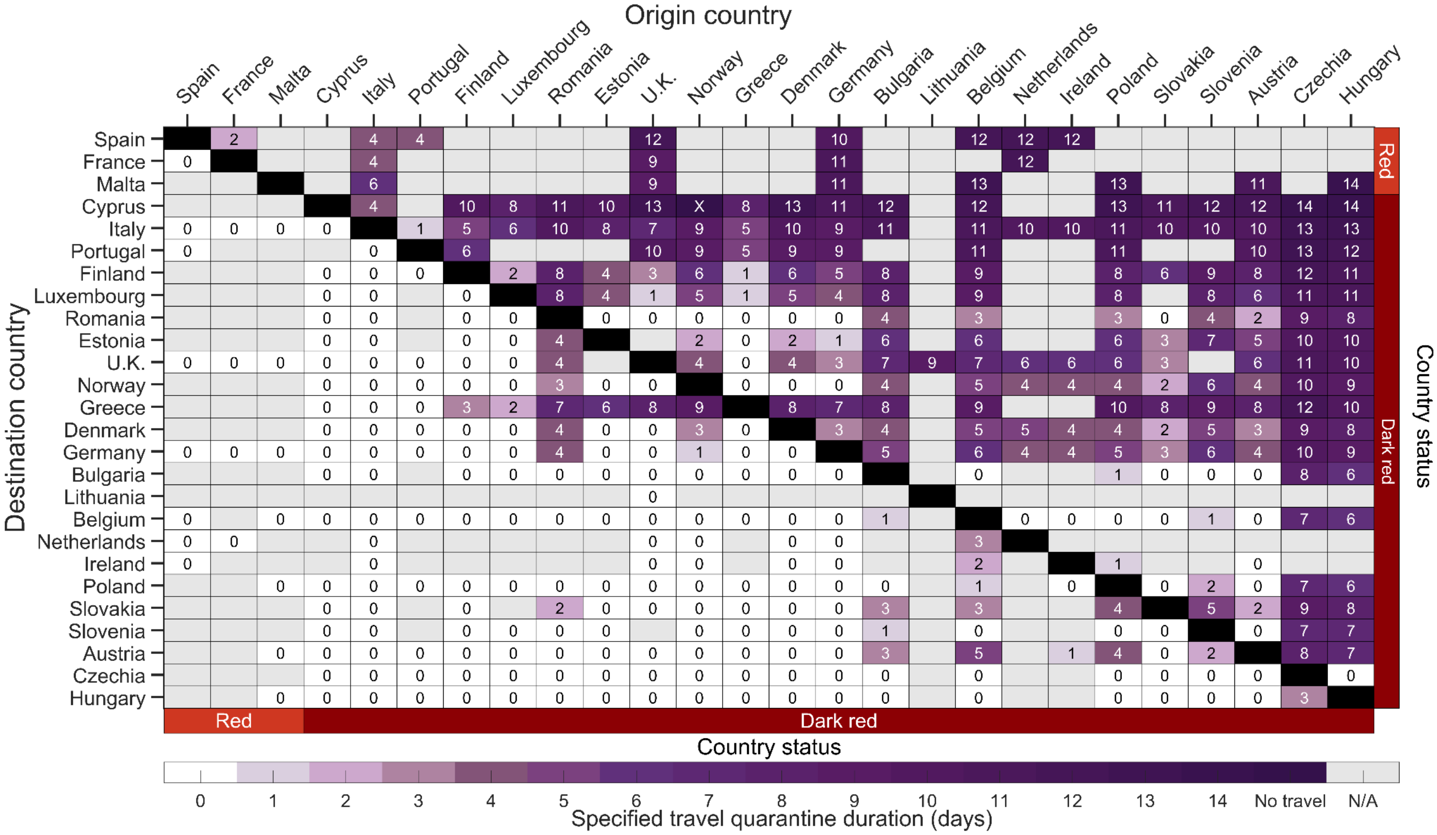


**Figure S18.** The estimated minimum duration of travel quarantine with no testing for specified origin-destination country pairs that reduces within-country imminent infections to be equivalent to border closure for the country-paired analysis for an incubation period of 11.66 days. Specifying age-dependent vaccine effectiveness and proportion of asymptomatic infections, as well as country-specific demographics, incidence, prevalence of non-isolated infections, vaccine coverage, seroprevalence and travel flow, we determine the minimum duration of travel quarantine with no test conducted (colour gradient) that should be stated by the destination country for individuals arriving from the origin country. We consider travel quarantine durations of zero-days (white) to no travel (dark purple, i.e., specified travel quarantine can exceed 14 days). Within-country travel quarantine is not evaluated in the analysis (black). Travel flow data was not available for all country pairs (grey). The countries are ranked based on their estimated incidence per 100,000 over the last two weeks (November 8 to November 21), and stratified based on the European Union country classification system: Green, < 25 cases per 100,000; Amber, 25 to 150 cases per 100,000; Red, 150 to 500 cases per 100,000; and Dark Red, > 500 cases per 100,000.

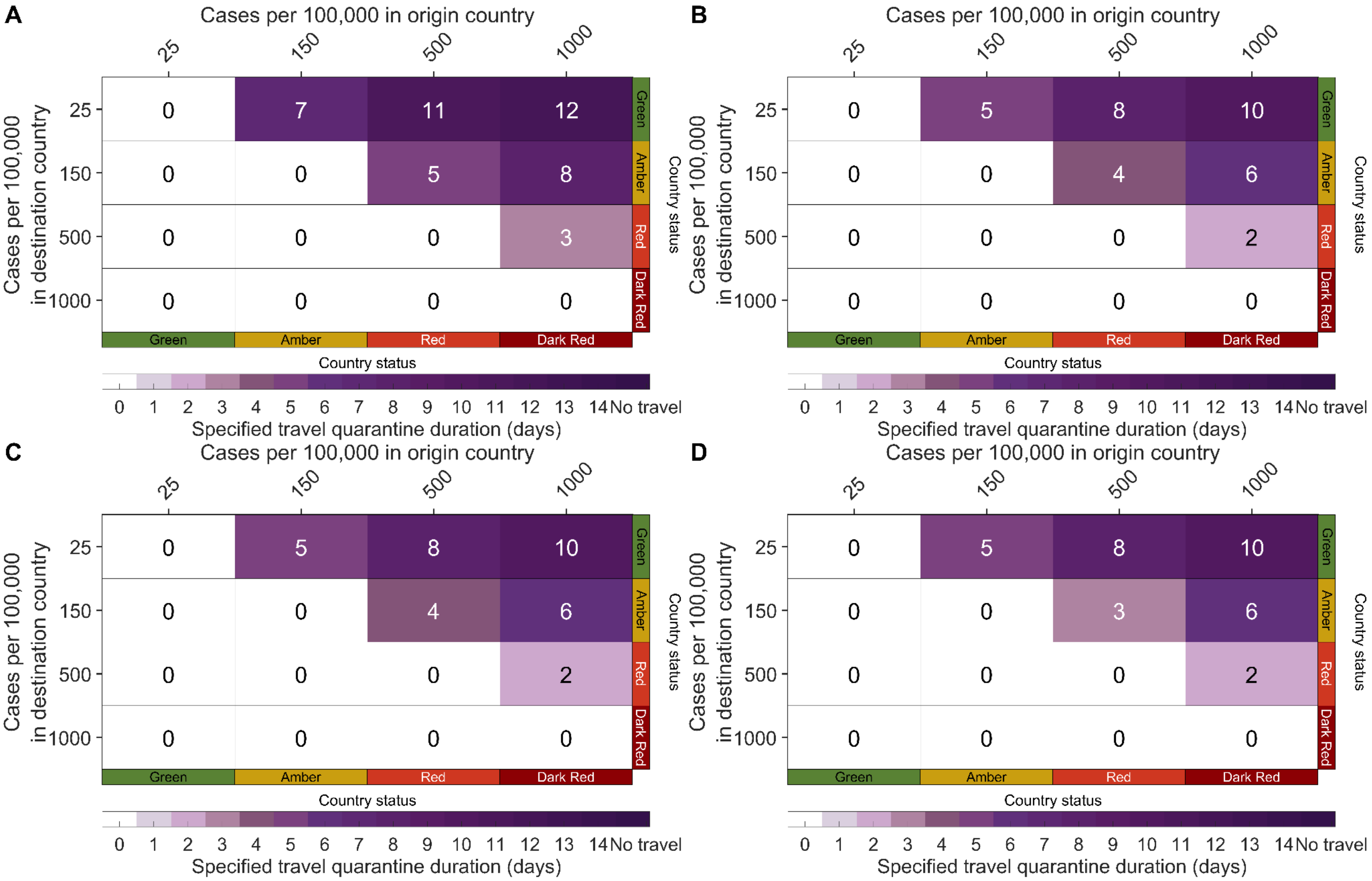


**Figure S19.** The estimated minimum duration of travel quarantine for specified origin-destination country pairs based on the European Union risk states that reduces within-country imminent infections to be equivalent to border closure for an incubation period of 11.66 days**.** Specifying age-dependent vaccine effectiveness and proportion of asymptomatic infections, as well as European demographics, 42% vaccine elicited immunity, 32% natural immunity, we determine the minimum travel duration of quarantine (colour gradient) that should be stated by the destination country for individuals arriving from the origin country. We consider travel quarantine durations of zero days (white) to a travel ban (dark purple, i.e., specified quarantine exceeding 14 days) with **A**) no test, **B**) an RT-PCR test on exit from quarantine, **C**) a rapid antigen test on exit from quarantine, and **D**) a rapid antigen test on both entry to and exit from quarantine. For travel quarantine durations of one day or longer, the RT-PCR test was conducted 24 h before exit from quarantine. and no delay in obtaining the rapid antigen test. For a zero-day travel quarantine, the RT-PCR test was conducted 24 h before travel.

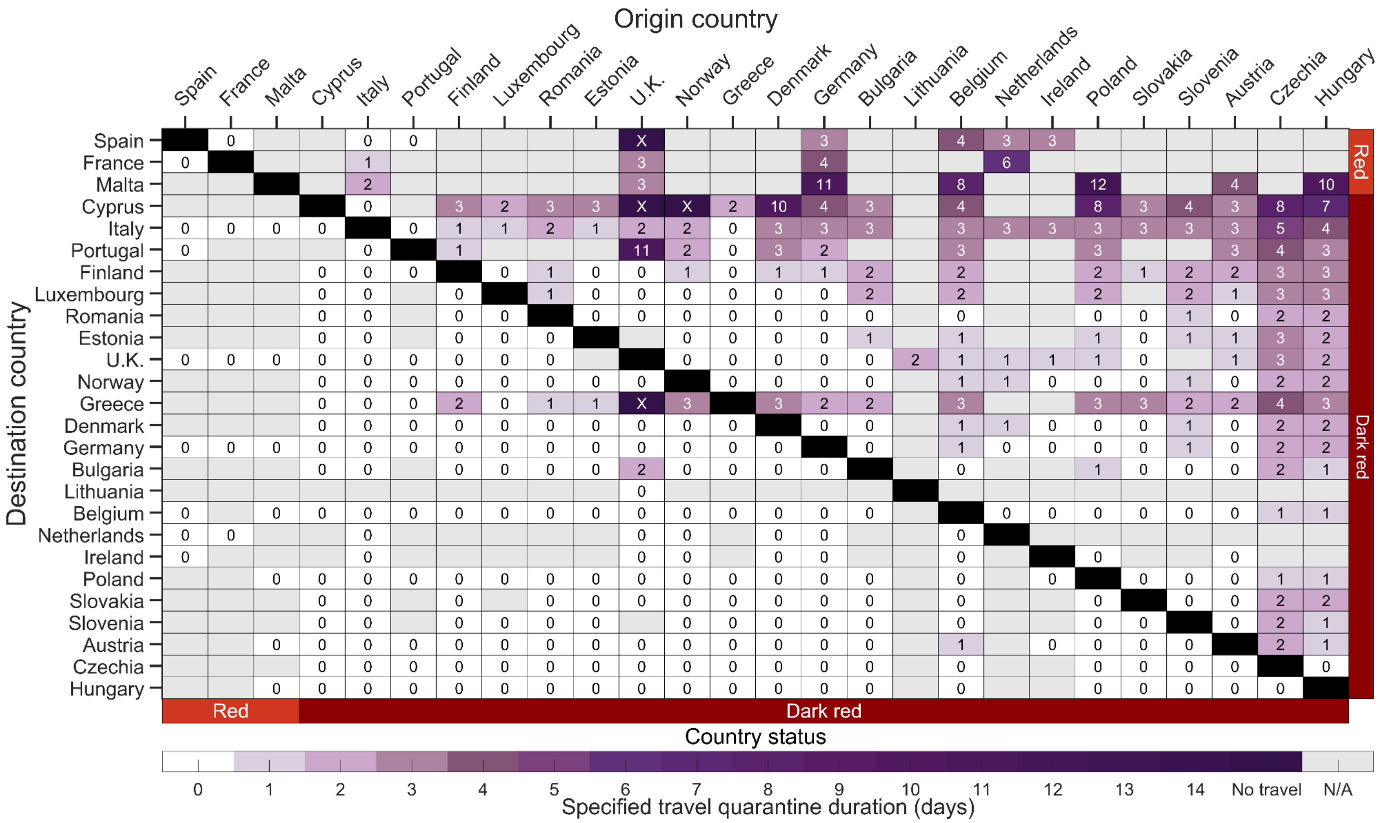


**Figure S20.** The estimated minimum duration of travel quarantine with an RT-PCR test on exit from quarantine for specified origin-destination country pairs that reduces within-country imminent infections to be equivalent to border closure with 75% adherence to self-isolation upon symptom onset. Specifying an incubation period of 5.72 days, age-dependent vaccine effectiveness and proportion of asymptomatic infections, as well as country-specific demographics, incidence, prevalence of non-isolated infections, vaccine coverage, seroprevalence and travel flow, we determine the minimum duration of travel quarantine with an RT-PCR test on exit from quarantine (colour gradient) that should be stated by the destination country for individuals arriving from the origin country. We consider travel quarantine durations of zero-days (white) to no travel (dark purple, i.e., specified travel quarantine can exceed 14 days). Within-country travel quarantine is not evaluated in the analysis (black). Travel flow data was not available for all country pairs (grey). The countries are ranked based on their estimated incidence per 100,000 over the last two weeks (November 8 to November 21), and stratified based on the European Union country classification system: Green, < 25 cases per 100,000; Amber, 25 to 150 cases per 100,000; Red, 150 to 500 cases per 100,000; and Dark Red, > 500 cases per 100,000. For travel quarantine durations of one day or longer, the RT-PCR test was conducted 24-h before exit from quarantine. For a zero-day travel quarantine, the RT-PCR test was conducted 24-h before travel.

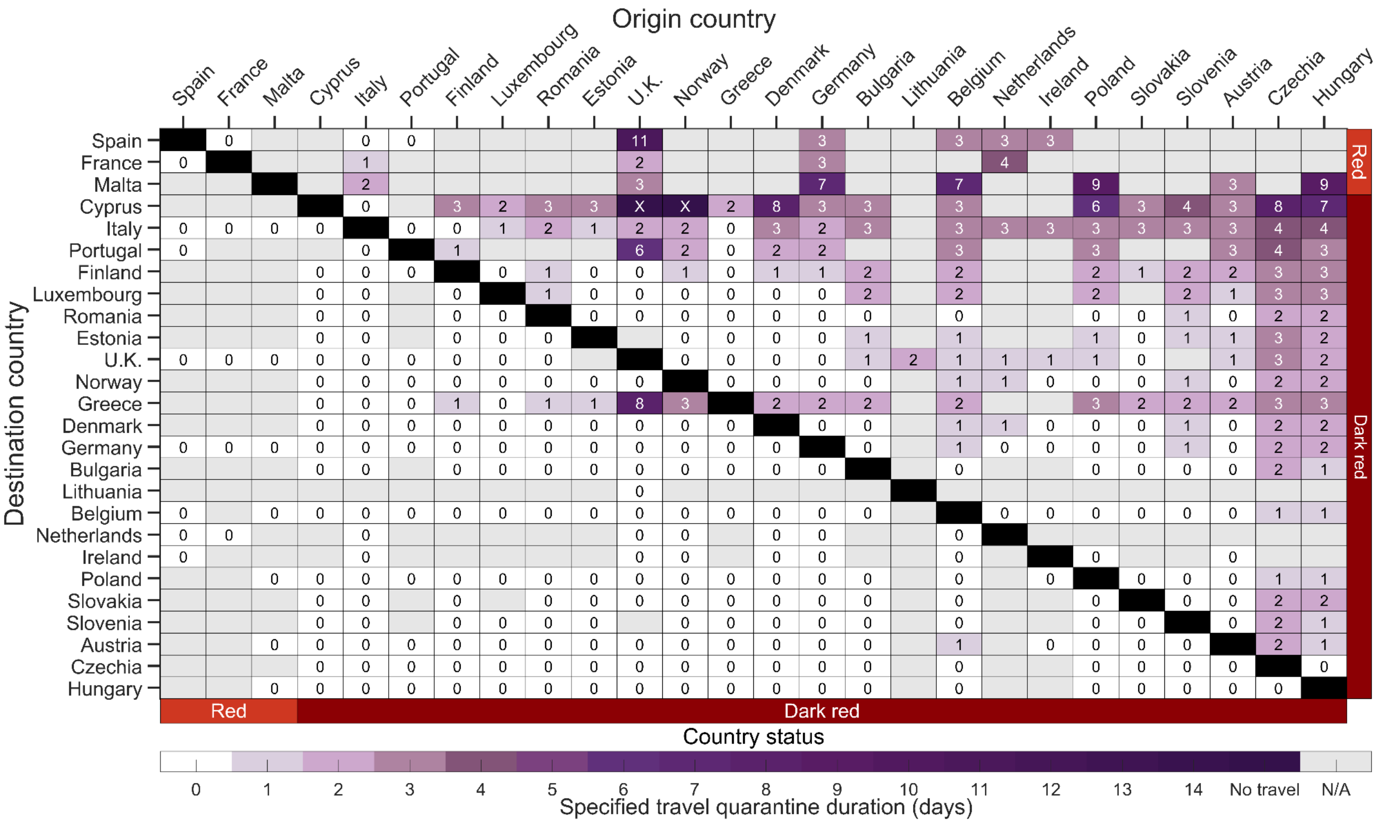


**Figure S21.** The estimated minimum duration of travel quarantine with an RT-PCR test on exit from quarantine for specified origin-destination country pairs that reduces within-country imminent infections to be equivalent to border closure with 50% adherence to self-isolation upon symptom onset. Specifying an incubation period of 5.72 days, age-dependent vaccine effectiveness and proportion of asymptomatic infections, as well as country-specific demographics, incidence, prevalence of non-isolated infections, vaccine coverage, seroprevalence and travel flow, we determine the minimum duration of travel quarantine with an RT-PCR test on exit from quarantine (colour gradient) that should be stated by the destination country for individuals arriving from the origin country. We consider travel quarantine durations of zero-days (white) to no travel (dark purple, i.e., specified travel quarantine can exceed 14 days). Within-country travel quarantine is not evaluated in the analysis (black). Travel flow data was not available for all country pairs (grey). The countries are ranked based on their estimated incidence per 100,000 over the last two weeks (November 8 to November 21), and stratified based on the European Union country classification system: Green, < 25 cases per 100,000; Amber, 25 to 150 cases per 100,000; Red, 150 to 500 cases per 100,000; and Dark Red, > 500 cases per 100,000. For travel quarantine durations of one day or longer, the RT-PCR test was conducted 24-h before exit from quarantine. For a zero-day travel quarantine, the RT-PCR test was conducted 24-h before travel.

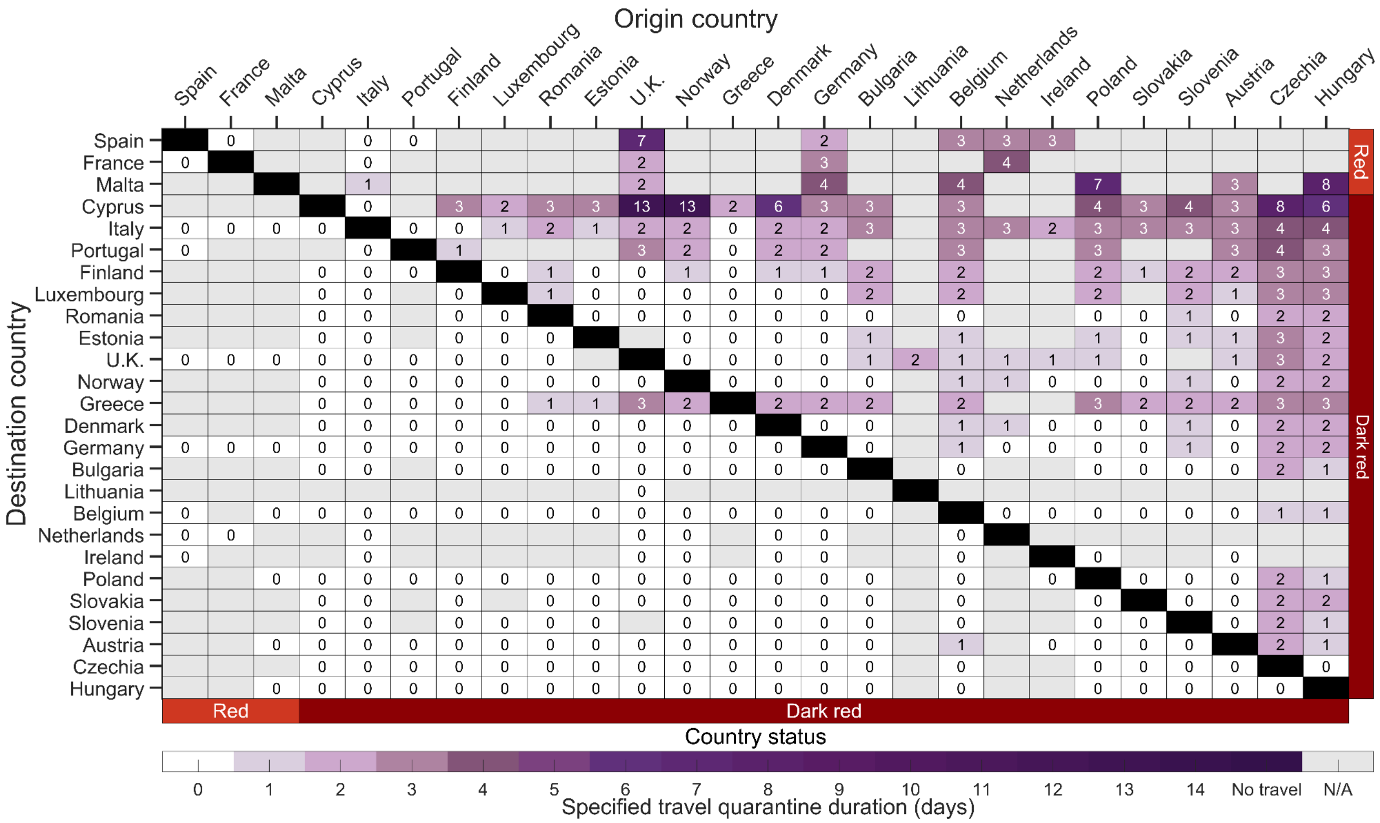


**Figure S22.** The estimated minimum duration of travel quarantine with an RT-PCR test on exit from quarantine for specified origin-destination country pairs that reduces within-country imminent infections to be equivalent to border closure with 25% adherence to self-isolation upon symptom onset. Specifying an incubation period of 5.72 days, age-dependent vaccine effectiveness and proportion of asymptomatic infections, as well as country-specific demographics, incidence, prevalence of non-isolated infections, vaccine coverage, seroprevalence and travel flow, we determine the minimum duration of travel quarantine with an RT-PCR test on exit from quarantine (colour gradient) that should be stated by the destination country for individuals arriving from the origin country. We consider travel quarantine durations of zero-days (white) to no travel (dark purple, i.e., specified travel quarantine can exceed 14 days). Within-country travel quarantine is not evaluated in the analysis (black). Travel flow data was not available for all country pairs (grey). The countries are ranked based on their estimated incidence per 100,000 over the last two weeks (November 8 to November 21), and stratified based on the European Union country classification system: Green, < 25 cases per 100,000; Amber, 25 to 150 cases per 100,000; Red, 150 to 500 cases per 100,000; and Dark Red, > 500 cases per 100,000. For travel quarantine durations of one day or longer, the RT-PCR test was conducted 24-h before exit from quarantine. For a zero-day travel quarantine, the RT-PCR test was conducted 24-h before travel.

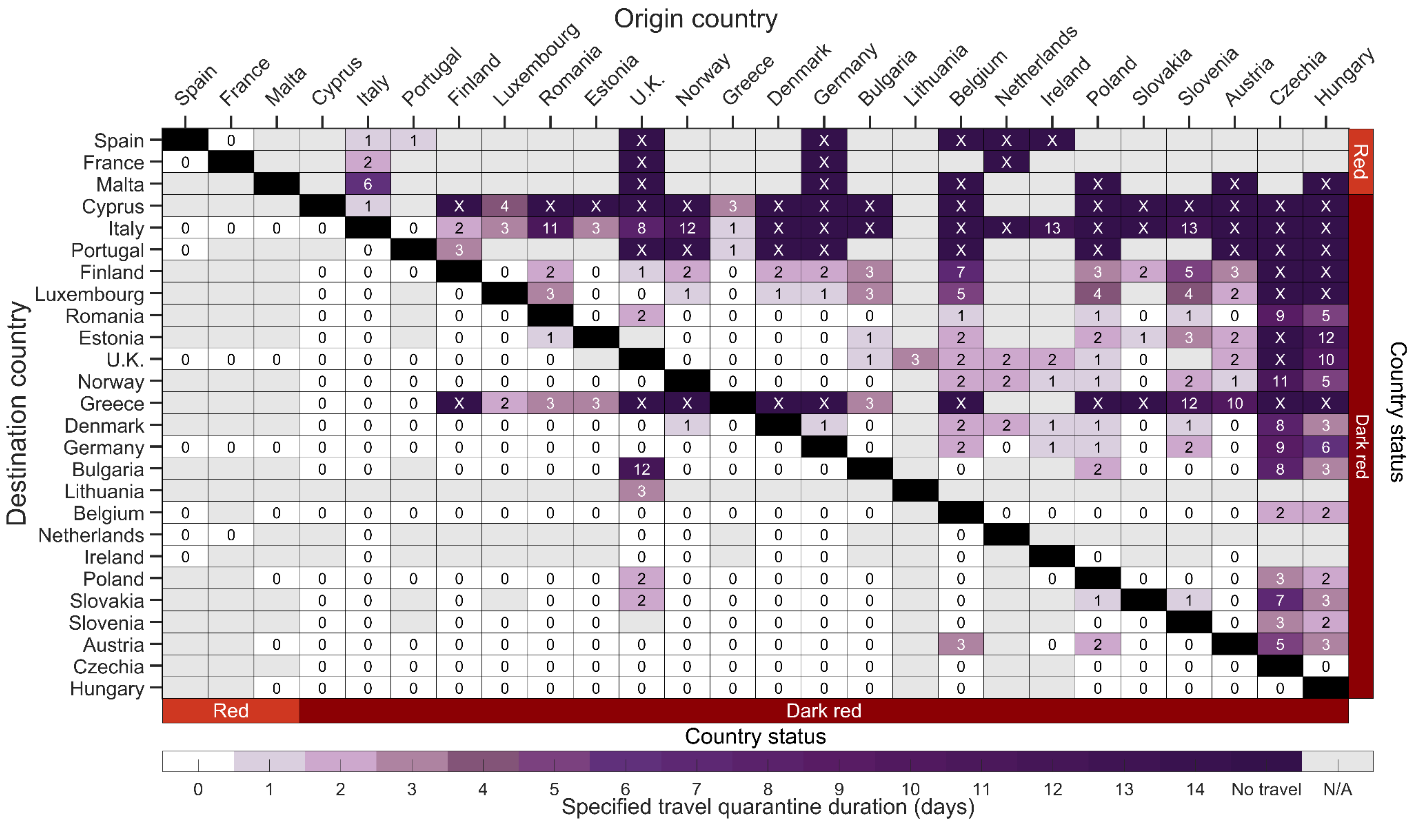


**Figure S23.** The estimated minimum duration of travel quarantine with an RT-PCR test on exit from quarantine for specified origin-destination country pairs that reduces within-country imminent infections to be equivalent to border closure with 75% adherence to the quarantine policy. Specifying an incubation period of 5.72 days, age-dependent vaccine effectiveness and proportion of asymptomatic infections, as well as country-specific demographics, incidence, prevalence of non-isolated infections, vaccine coverage, seroprevalence and travel flow, we determine the minimum duration of travel quarantine with an RT-PCR test on exit from quarantine (colour gradient) that should be stated by the destination country for individuals arriving from the origin country. We consider travel quarantine durations of zero-days (white) to no travel (dark purple, i.e., specified travel quarantine can exceed 14 days). Within-country travel quarantine is not evaluated in the analysis (black). Travel flow data was not available for all country pairs (grey). The countries are ranked based on their estimated incidence per 100,000 over the last two weeks (November 8 to November 21), and stratified based on the European Union country classification system: Green, < 25 cases per 100,000; Amber, 25 to 150 cases per 100,000; Red, 150 to 500 cases per 100,000; and Dark Red, > 500 cases per 100,000. For travel quarantine durations of one day or longer, the RT-PCR test was conducted 24-h before exit from quarantine. For a zero-day travel quarantine, the RT-PCR test was conducted 24-h before travel.

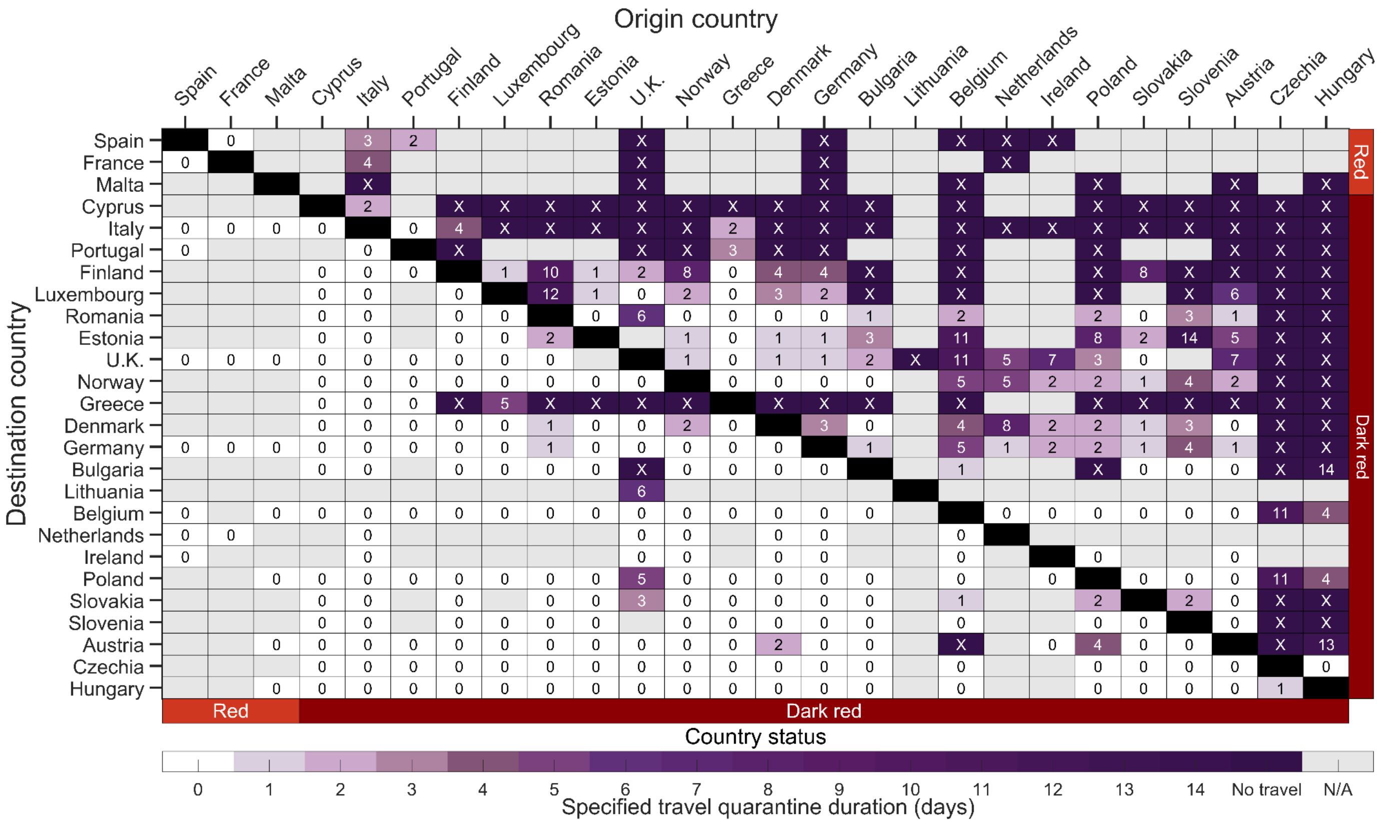


**Figure S24.** The estimated minimum duration of travel quarantine with an RT-PCR test on exit from quarantine for specified origin-destination country pairs that reduces within-country imminent infections to be equivalent to border closure with 50% adherence to the quarantine policy. Specifying an incubation period of 5.72 days, age-dependent vaccine effectiveness and proportion of asymptomatic infections, as well as country-specific demographics, incidence, prevalence of non-isolated infections, vaccine coverage, seroprevalence and travel flow, we determine the minimum duration of travel quarantine with an RT-PCR test on exit from quarantine (colour gradient) that should be stated by the destination country for individuals arriving from the origin country. We consider travel quarantine durations of zero-days (white) to no travel (dark purple, i.e., specified travel quarantine can exceed 14 days). Within-country travel quarantine is not evaluated in the analysis (black). Travel flow data was not available for all country pairs (grey). The countries are ranked based on their estimated incidence per 100,000 over the last two weeks (November 8 to November 21), and stratified based on the European Union country classification system: Green, < 25 cases per 100,000; Amber, 25 to 150 cases per 100,000; Red, 150 to 500 cases per 100,000; and Dark Red, > 500 cases per 100,000. For travel quarantine durations of one day or longer, the RT-PCR test was conducted 24-h before exit from quarantine. For a zero-day travel quarantine, the RT-PCR test was conducted 24-h before travel.

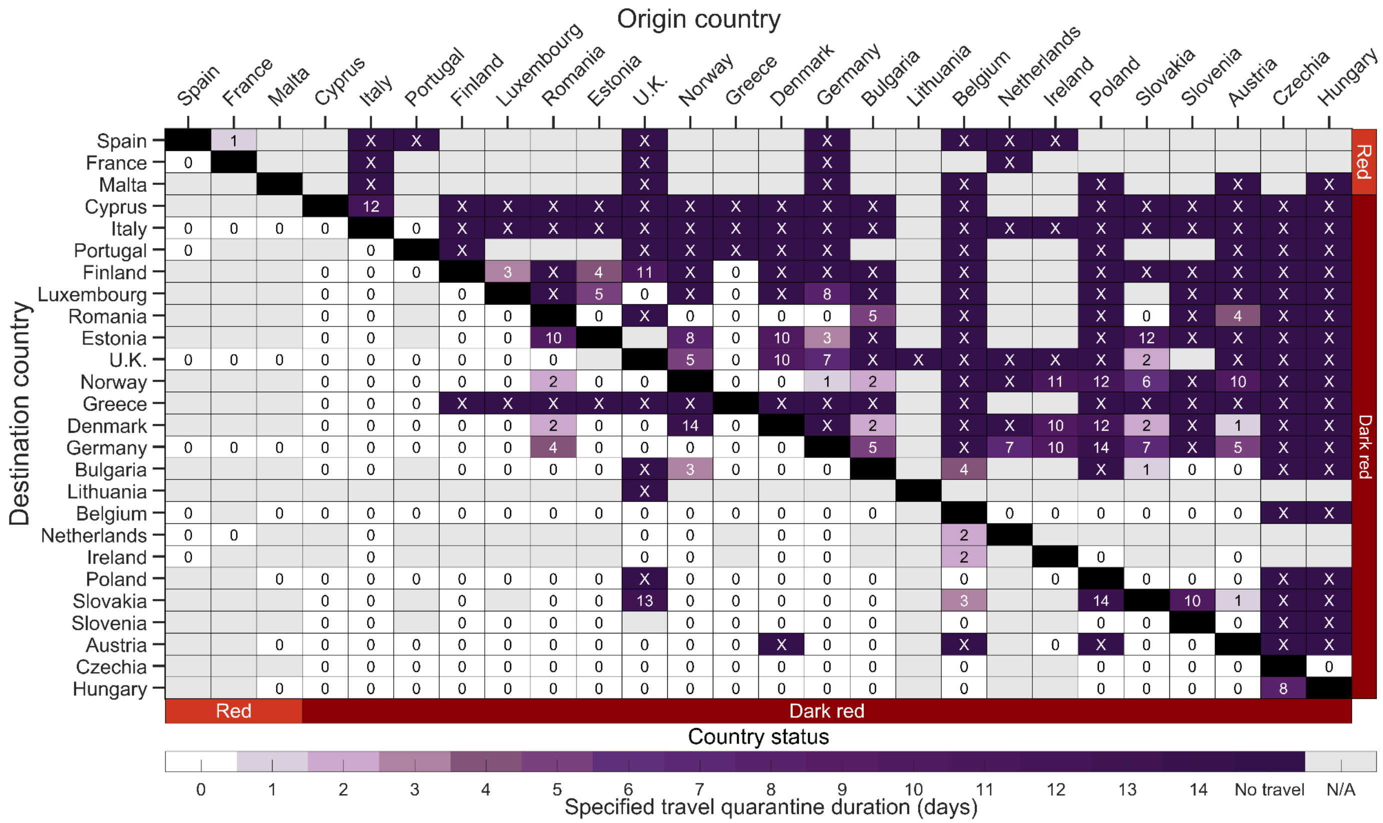


**Figure S25.** The estimated minimum duration of travel quarantine with an RT-PCR test on exit from quarantine for specified origin-destination country pairs that reduces within-country imminent infections to be equivalent to border closure with 25% adherence to the quarantine policy. Specifying an incubation period of 5.72 days, age-dependent vaccine effectiveness and proportion of asymptomatic infections, as well as country-specific demographics, incidence, prevalence of non-isolated infections, vaccine coverage, seroprevalence and travel flow, we determine the minimum duration of travel quarantine with an RT-PCR test on exit from quarantine (colour gradient) that should be stated by the destination country for individuals arriving from the origin country. We consider travel quarantine durations of zero-days (white) to no travel (dark purple, i.e., specified travel quarantine can exceed 14 days). Within-country travel quarantine is not evaluated in the analysis (black). Travel flow data was not available for all country pairs (grey). The countries are ranked based on their estimated incidence per 100,000 over the last two weeks (November 8 to November 21), and stratified based on the European Union country classification system: Green, < 25 cases per 100,000; Amber, 25 to 150 cases per 100,000; Red, 150 to 500 cases per 100,000; and Dark Red, > 500 cases per 100,000. For travel quarantine durations of one day or longer, the RT-PCR test was conducted 24-h before exit from quarantine. For a zero-day travel quarantine, the RT-PCR test was conducted 24-h before travel.

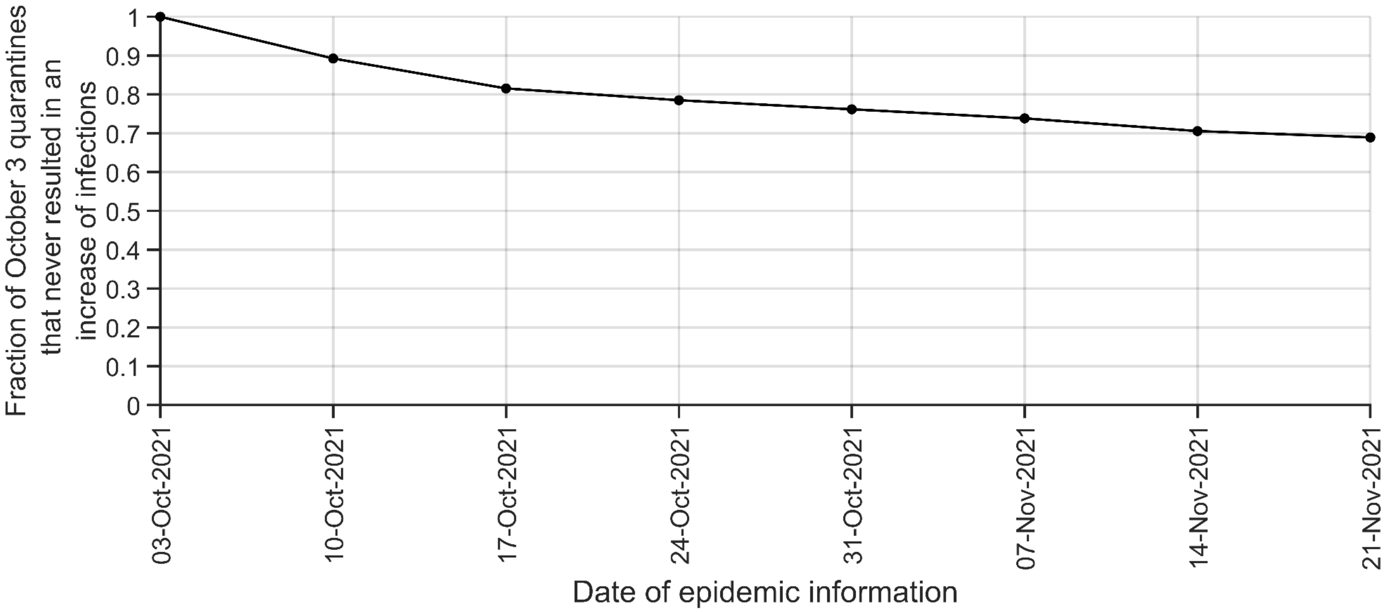


**Figure S26.** The durability of a calculation of the minimum sufficient quarantine duration. The fraction of country pairings in which the quarantine specified on June 20, 2021 did not result in imminent infections greater than border closure (i.e. minimum sufficient or greater than the minimum) at weekly intervals upto November 21, 2021.

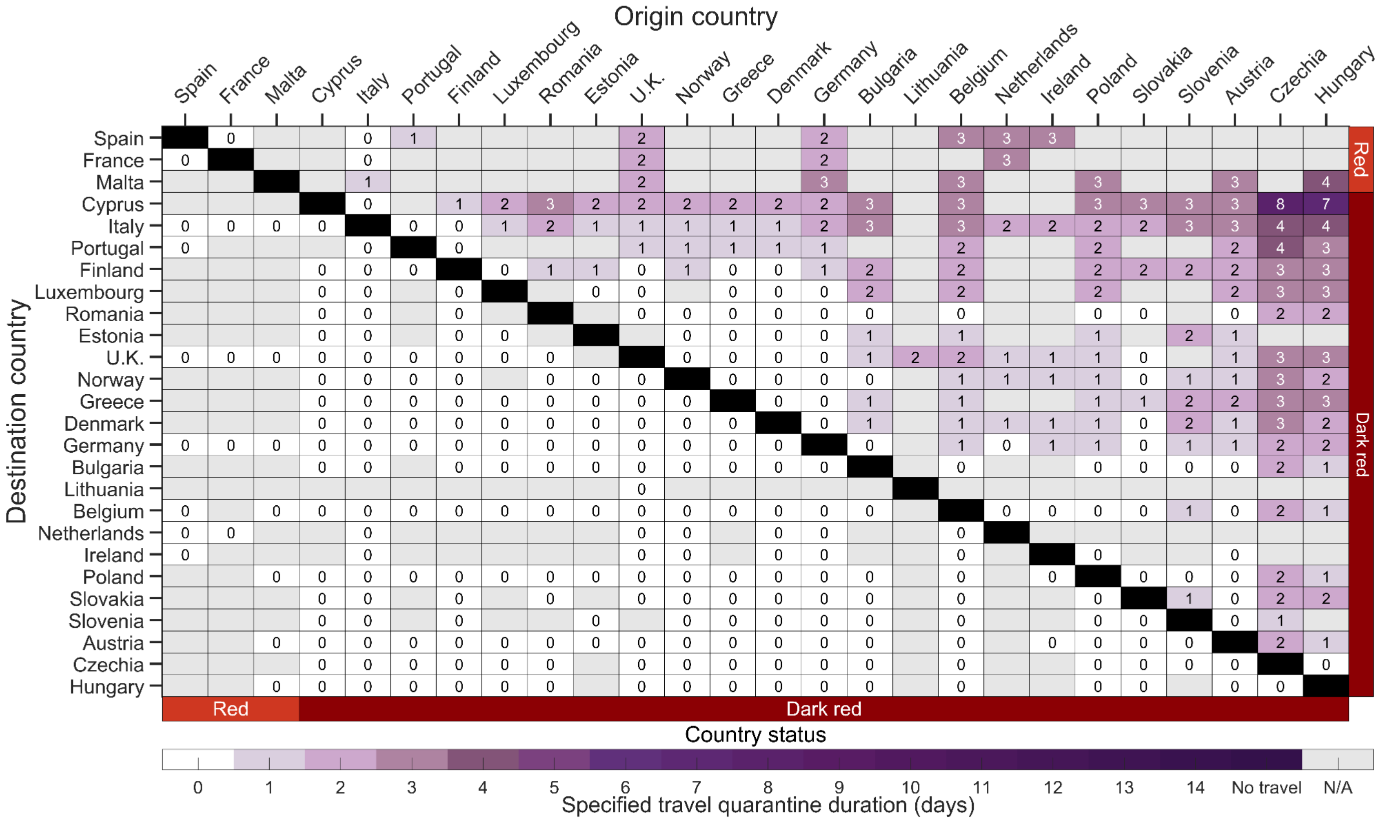


**Figure S27.** The estimated minimum duration of travel quarantine with an RT-PCR test on exit from quarantine for specified origin-destination country pairs that reduces within-country imminent infections to be equivalent to border closure for travel flow informed by air passenger transport between countries. Specifying an incubation period of 5.72 days, age-dependent vaccine effectiveness and proportion of asymptomatic infections, as well as country-specific demographics, incidence, prevalence of non-isolated infections, vaccine coverage, seroprevalence and travel flow, we determine the minimum duration of travel quarantine with an RT-PCR test on exit from quarantine (colour gradient) that should be stated by the destination country for individuals arriving from the origin country. We consider travel quarantine durations of zero-days (white) to no travel (dark purple, i.e., specified travel quarantine can exceed 14 days). Within-country travel quarantine is not evaluated in the analysis (black). Travel flow data was not available for all country pairs (grey). The countries are ranked based on their estimated incidence per 100,000 over the last two weeks (November 8 to November 21), and stratified based on the European Union country classification system: Green, < 25 cases per 100,000; Amber, 25 to 150 cases per 100,000; Red, 150 to 500 cases per 100,000; and Dark Red, > 500 cases per 100,000. For travel quarantine durations of one day or longer, the RT-PCR test was conducted 24-h before exit from quarantine. For a zero-day travel quarantine, the RT-PCR test was conducted 24-h before travel.

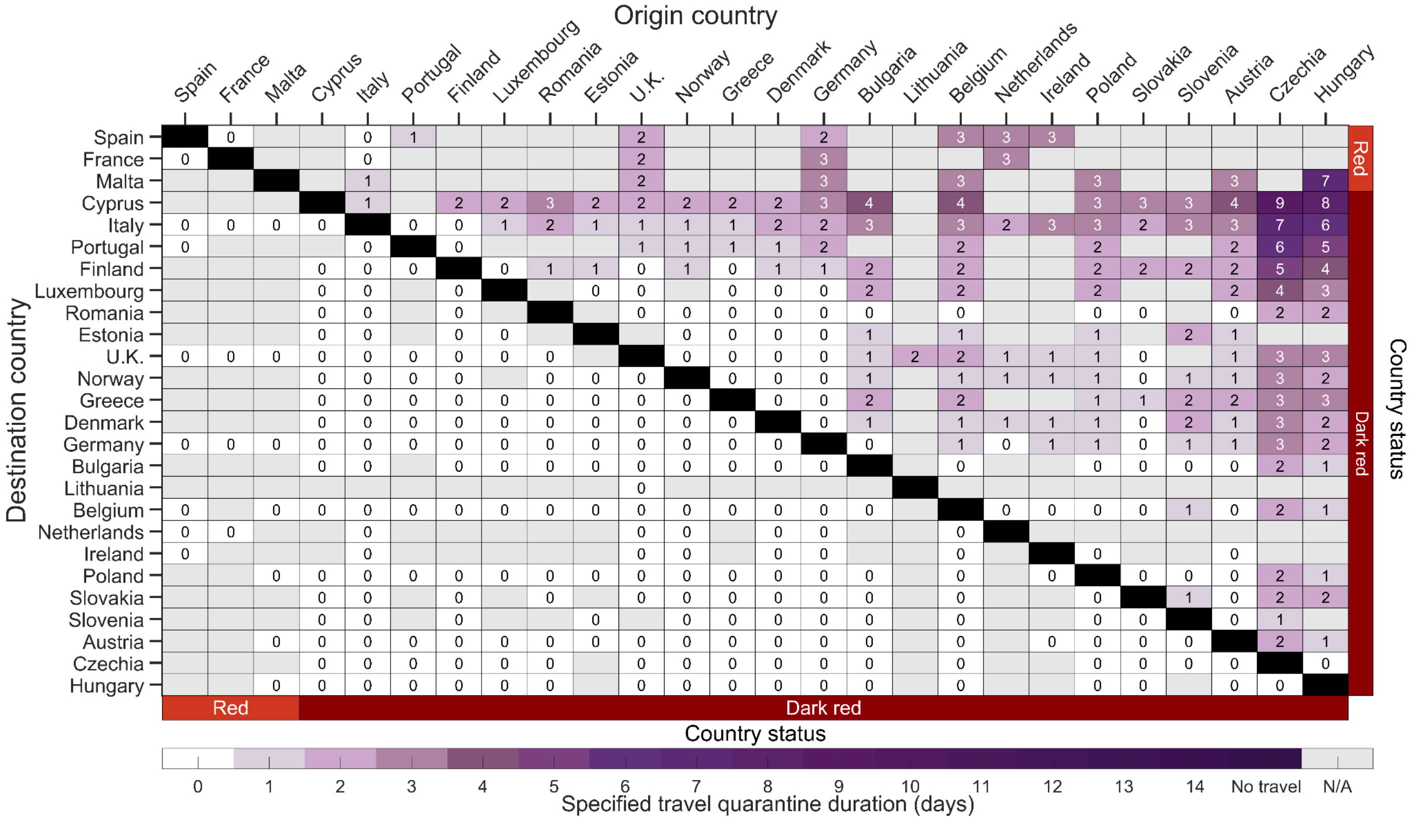


**Figure S28.** The estimated minimum duration of travel quarantine with a rapid antigen test on exit from quarantine for specified origin-destination country pairs that reduces within-country imminent infections to be equivalent to border closure for the country-paired analysis for travel flow informed by air passenger transport between countries. Specifying an incubation period of 5.72 days, age-dependent vaccine effectiveness and proportion of asymptomatic infections, as well as country-specific demographics, incidence, prevalence of non-isolated infections, vaccine coverage, seroprevalence and travel flow, we determine the minimum duration of travel quarantine with a rapid antigen test on exit (colour gradient) that should be stated by the destination country for individuals arriving from the origin country. We consider travel quarantine durations of zero-days (white) to no travel (dark purple, i.e., specified travel quarantine can exceed 14 days). Within-country travel quarantine is not evaluated in the analysis (black). Travel flow data was not available for all country pairs (grey). The countries are ranked based on their estimated incidence per 100,000 over the last two weeks (November 8 to November 21), and stratified based on the European Union country classification system: Green, < 25 cases per 100,000; Amber, 25 to 150 cases per 100,000; Red, 150 to 500 cases per 100,000; and Dark Red, > 500 cases per 100,000. We assumed that there was no delay in obtaining the test result from a rapid antigen test.

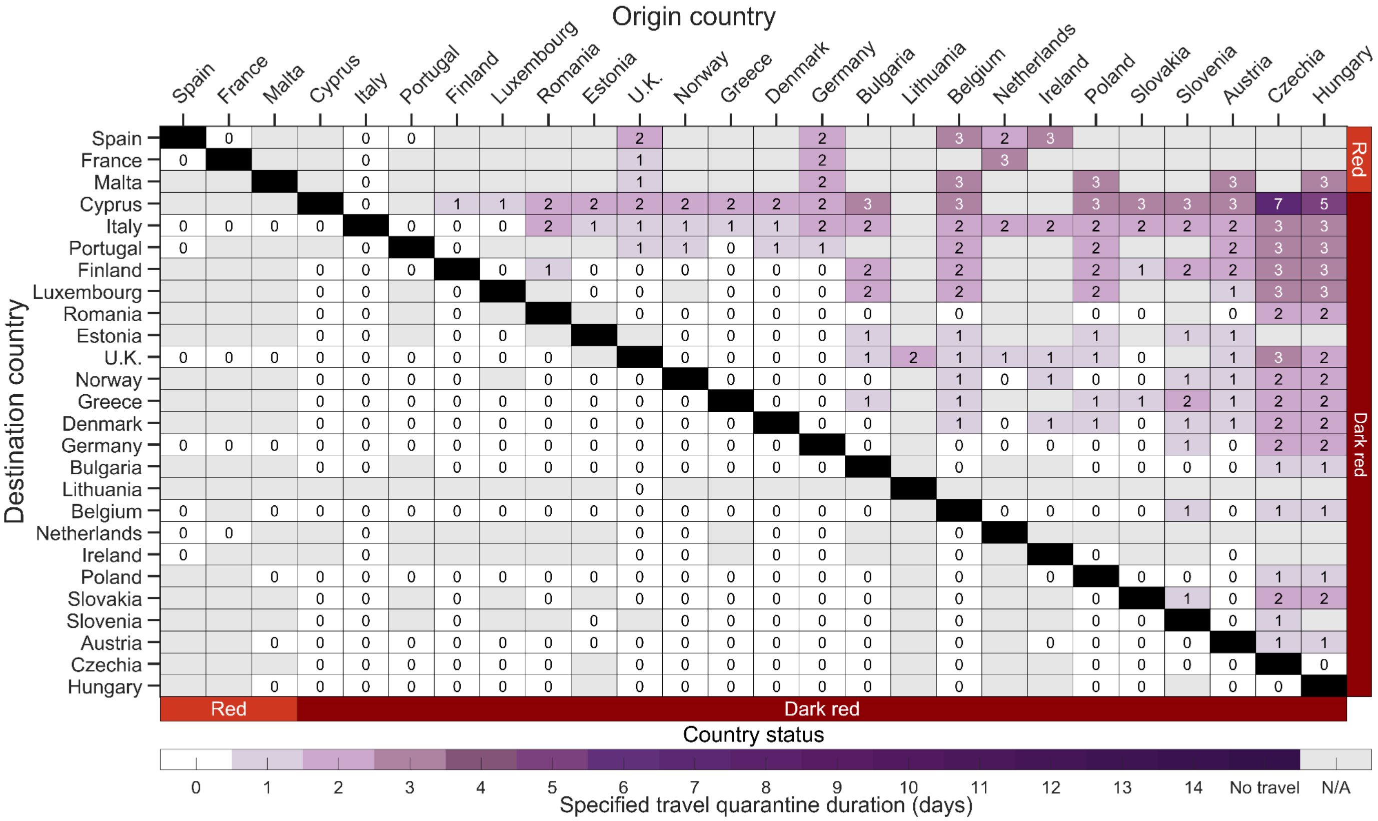

**Figure S29.** The estimated minimum duration of travel quarantine with a rapid antigen test on entry to and exit from quarantine for specified origin-destination country pairs that reduces within-country imminent infections to be equivalent to border closure for the country-paired analysis for travel flow informed by air passenger transport between countries. Specifying an incubation period of 5.72 days, age-dependent vaccine effectiveness and proportion of asymptomatic infections, as well as country-specific demographics, incidence, prevalence of non-isolated infections, vaccine coverage, seroprevalence and travel flow, we determine the minimum duration of travel quarantine with a rapid antigen test on entry and exit (colour gradient) that should be stated by the destination country for individuals arriving from the origin country. We consider travel quarantine durations of zero-days (white) to no travel (dark purple, i.e., specified travel quarantine can exceed 14 days). Within–country travel quarantine is not evaluated in the analysis (black). Travel flow data was not available for all country pairs (grey). The countries are ranked based on their estimated incidence per 100,000 over the last two weeks (November 8 to November 21), and stratified based on the European Union country classification system: Green, < 25 cases per 100,000; Amber, 25 to 150 cases per 100,000; Red, 150 to 500 cases per 100,000; and Dark Red, > 500 cases per 100,000. There was assumed to be no delay in obtaining the test result from a rapid antigen test.

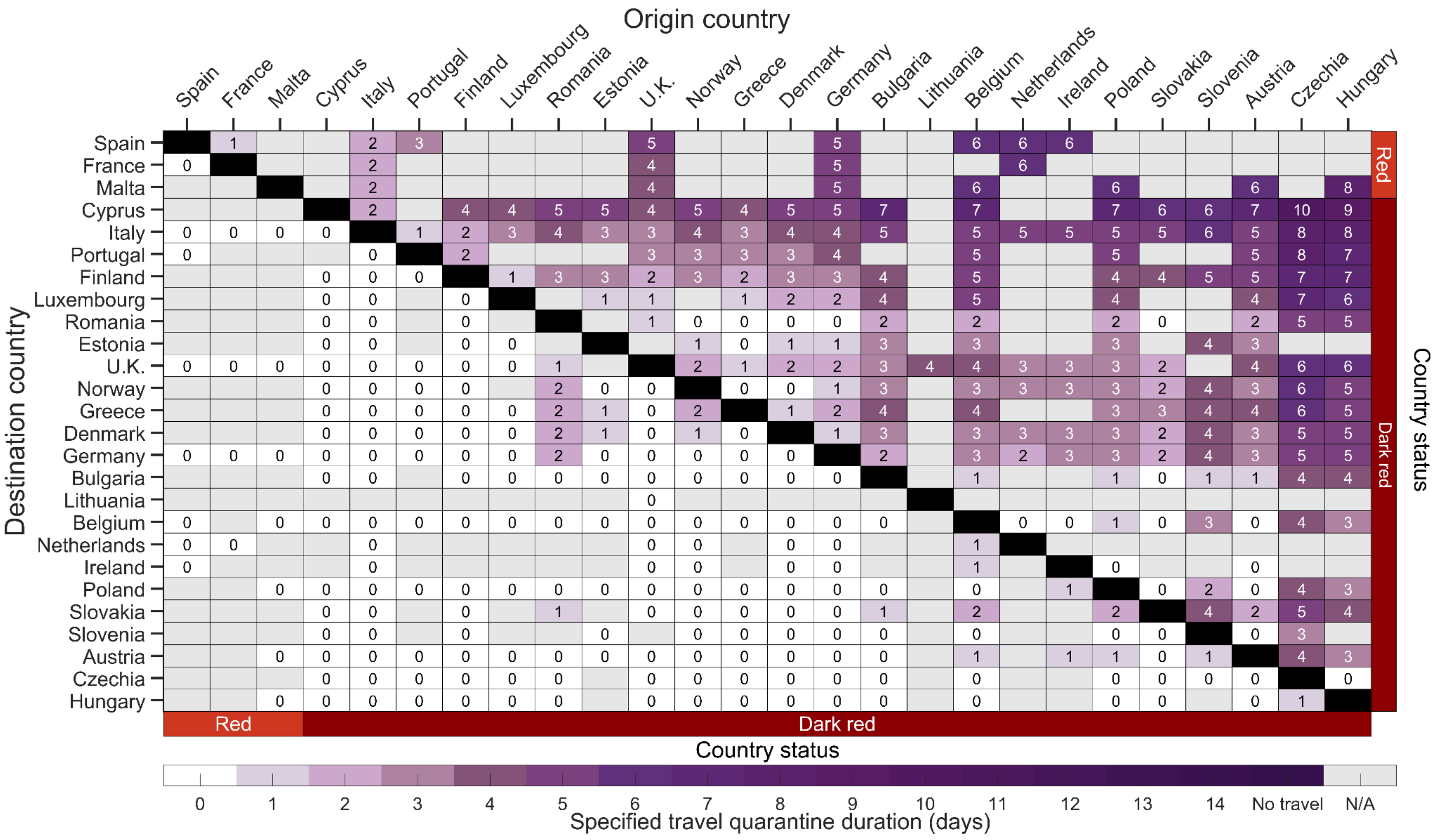

**Figure S30.** The estimated minimum duration of travel quarantine with no testing for specified origin-destination country pairs that reduces within-country imminent infections to be equivalent to border closure for the country-paired analysis for travel flow informed by air passenger transport between countries. Specifying an incubation period of 5.72 days, age-dependent vaccine effectiveness and proportion of asymptomatic infections, as well as country-specific demographics, incidence, prevalence of non-isolated infections, vaccine coverage, seroprevalence and travel flow, we determine the minimum duration of travel quarantine with no test conducted (colour gradient) that should be stated by the destination country for individuals arriving from the origin country. We consider travel quarantine durations of zero-days (white) to no travel (dark purple, i.e., specified travel quarantine can exceed 14 days). Within-country travel quarantine is not evaluated in the analysis (black). Travel flow data was not available for all country pairs (grey). The countries are ranked based on their estimated incidence per 100,000 over the last two weeks (November 8 to November 21), and stratified based on the European Union country classification system: Green, < 25 cases per 100,000; Amber, 25 to 150 cases per 100,000; Red, 150 to 500 cases per 100,000; and Dark Red, > 500 cases per 100,000.

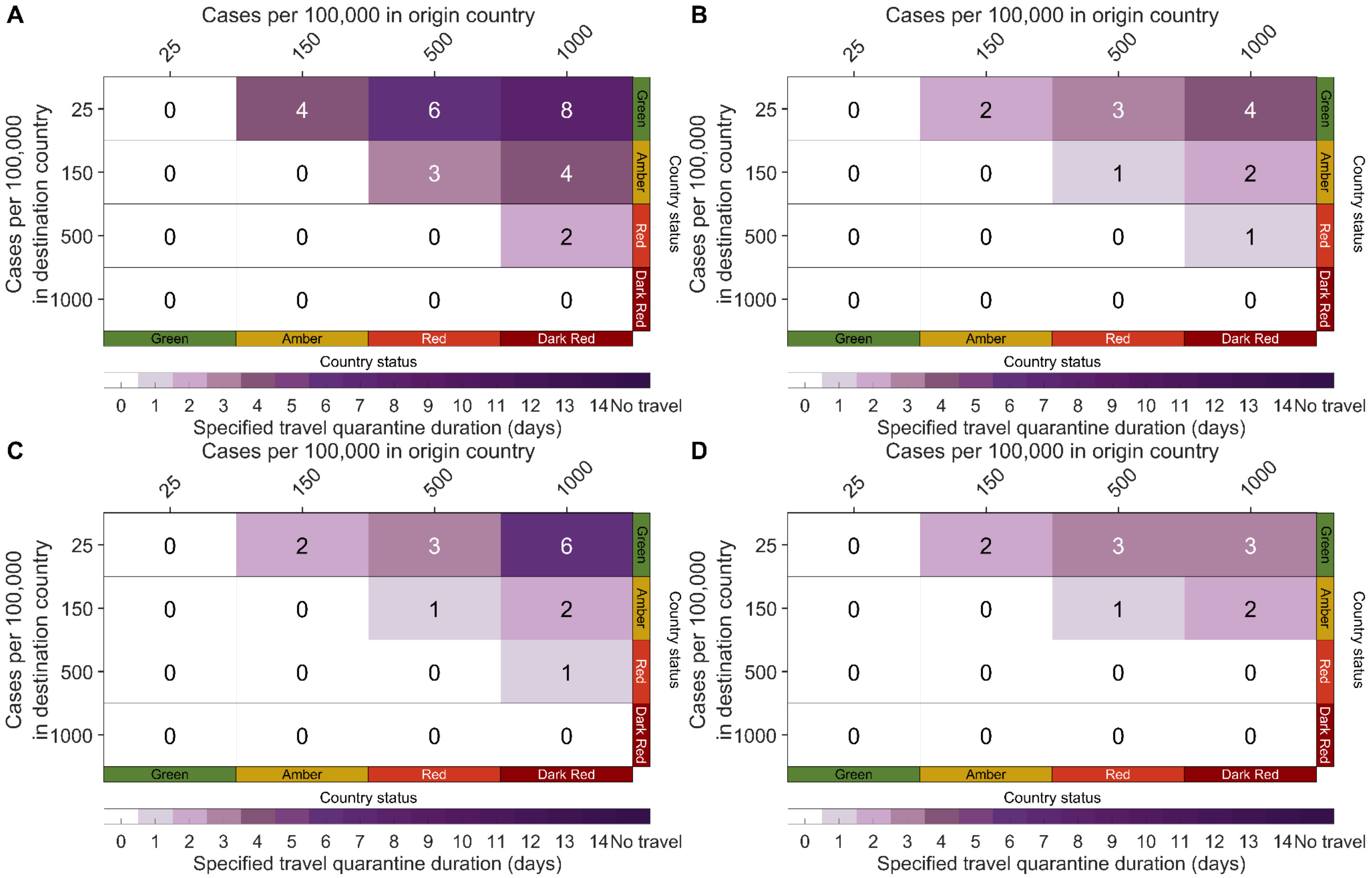


**Figure S31.** The estimated minimum duration of travel quarantine for specified origin-destination country pairs based on the European Union risk states that reduces within-country imminent infections to be equivalent to border closure for travel flow informed by air passenger transport between countries.. Specifying an incubation period of 5.72 days, age-dependent vaccine effectiveness and proportion of asymptomatic infections, as well as European demographics, 42% vaccine elicited immunity, 32% natural immunity, we determine the minimum travel duration of quarantine (colour gradient) that should be stated by the destination country for individuals arriving from the origin country. We consider travel quarantine durations of zero days (white) to a travel ban (dark purple, i.e., specified quarantine exceeding 14 days) with **A**) no test, **B**) an RT-PCR test on exit from quarantine, **C**) a rapid antigen test on exit from quarantine, and **D**) a rapid antigen test on both entry to and exit from quarantine. For travel quarantine durations of one day or longer, the RT-PCR test was conducted 24 h before exit from quarantine. and no delay in obtaining the rapid antigen test. For a zero-day travel quarantine, the RT-PCR test was conducted 24 h before travel.

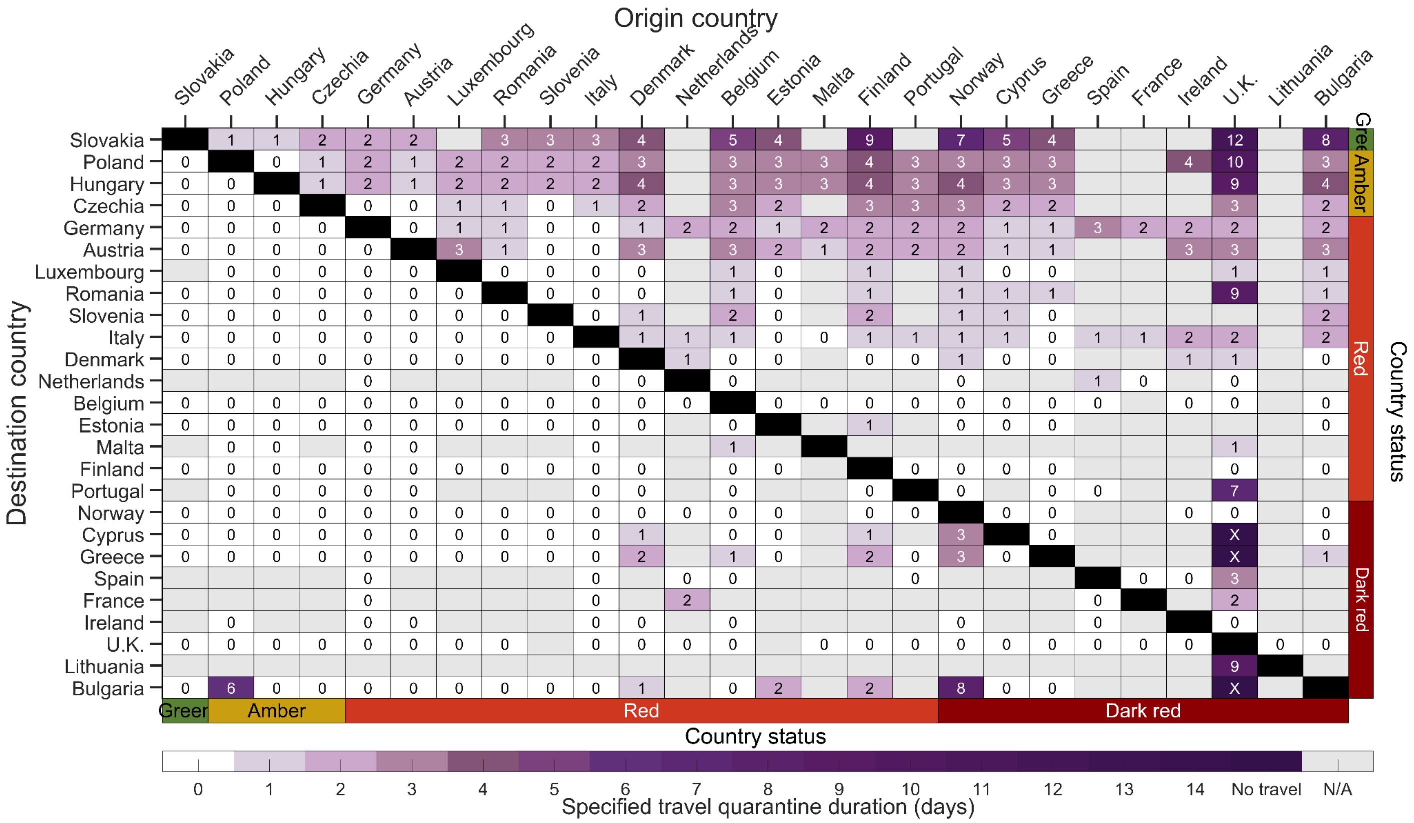


**Figure S32.** The estimated minimum duration of travel quarantine with an RT-PCR test on exit from quarantine for specified origin-destination country pairs that reduces within-country imminent infections to be equivalent to border closure based on the epidemic situation on August 8, 2021. Specifying an incubation period of 5.72 days, age-dependent vaccine effectiveness and proportion of asymptomatic infections, as well as country-specific demographics, incidence, prevalence of non-isolated infections, vaccine coverage, seroprevalence and travel flow, we determine the minimum duration of travel quarantine with an RT-PCR test on exit from quarantine (colour gradient) that should be stated by the destination country for individuals arriving from the origin country. We consider travel quarantine durations of zero-days (white) to no travel (dark purple, i.e., specified travel quarantine can exceed 14 days). Within-country travel quarantine is not evaluated in the analysis (black). Travel flow data was not available for all country pairs (grey). The countries are ranked based on their estimated incidence per 100,000 over the last two weeks (July 26 to August 8), and stratified based on the European Union country classification system: Green, < 25 cases per 100,000; Amber, 25 to 150 cases per 100,000; Red, 150 to 500 cases per 100,000; and Dark Red, > 500 cases per 100,000. For travel quarantine durations of one day or longer, the RT-PCR test was conducted 24-h before exit from quarantine. For a zero-day travel quarantine, the RT-PCR test was conducted 24-h before travel.

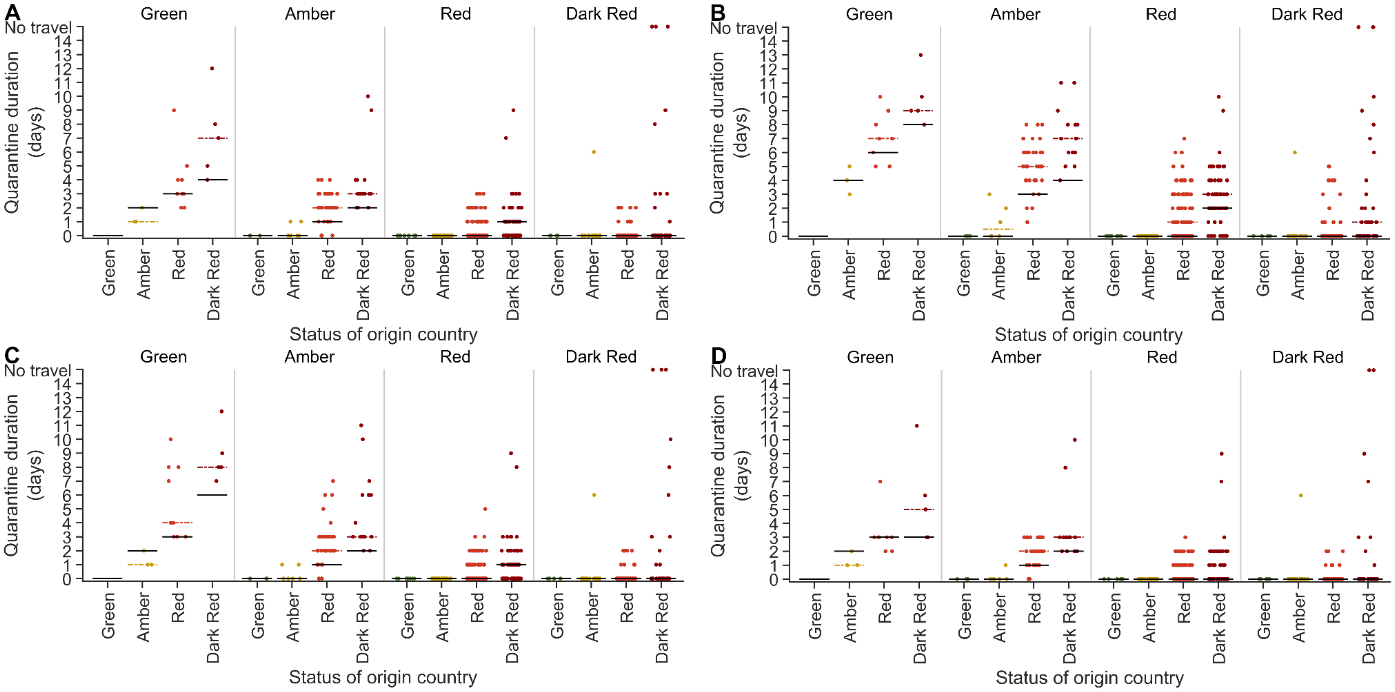


**Figure S33.** Comparison between the country-specific quarantine durations and the durations determined from the EU COVID classification system for the epidemic situation on August 8, 2021. The estimates for the minimum quarantine duration are stratified by destination country and origin country (*x* axis) for the country-pair analysis (dots; median dashed line) and our tier-based analysis (solid line) for a quarantine with **A**) a RT-PCR test on exit from quarantine, **B**) no testing conducted, **C**) rapid antigen test on exit from quarantine, and **D**) a rapid antigen test on both entry to and exit from quarantine. For travel quarantine durations of one day or longer, the RT-PCR test was conducted 24-h before exit from quarantine and with no delay in obtaining the rapid antigen test. For a zero-day travel quarantine, the RT-PCR test was conducted 24 h before travel.

#### Supplementary Tables

**Table S1.** References used for the travel flow and duration of stay for the 26 countries [a]

| Destination country | Data source for number of travellers into and duration of stay in the destination country | Year | Reference |
|---|---|---|---|
| Austria | TourMIS; Office for National Statistics (UK) | 2019 | [27,28] |
| Belgium | TourMIS; Office for National Statistics (UK) | 2019 | [27,28] |
| Bulgaria | NSI INFOSTAT; Office for National Statistics (UK) | 2019 | [28,29] |
| Czechia | TourMIS; Office for National Statistics (UK) | 2019 | [27,28] |
| Cyprus | TourMIS; Office for National Statistics (UK) | 2018 and 2019 | [27,28] |
| Denmark [b] | Statistics Denmark; Office for National Statistics (UK) | 2019 | [28,30,31] |
| Estonia | TourMIS | 2018 | [27] |
| Finland | TourMIS; Office for National Statistics (UK) | 2019 | [27,28] |
| France | Office for National Statistics (UK); Ministry of Economy for France | 2018 and 2019 | [28,32] |
| Germany | TourMIS; Office for National Statistics (UK) | 2019 | [27,28] |
| Greece | Hellenic Statistical Authority; TourMIS ; Office for National Statistics (UK) | 2019 | [27,28,33,34] |
| Hungary | TourMIS; Office for National Statistics (UK) | 2019 | [27,28] |
| Ireland | TourMIS; Failte Ireland; Office for National Statistics (UK) | 2019 | [27,28,35] |
| Italy | National Institute of Statistics, Italy; TourMIS; Office for National Statistics (UK) | 2019 | [27,28,36] |
| Lithuania | Office for National Statistics (UK) | 2019 | [28] |
| Luxembourg | Statistic Portal: Grand Duchy of Luxembourg; Office for National Statistics (UK) | 2019 | [28,37] |
| Malta | National Statistics Office - Malta; Office for National Statistics (UK) | 2019 | [28,38] |
| Netherlands | TourMIS; Office for National Statistics (UK) | 2019 | [27,28] |
| Norway [c] | Statistics Norway; TourMIS; Office for National Statistics (UK) | 2019 and 2021 | [28,39,40] |

| | | | |
|---|---|---|---|
| Poland | TourMIS; Office for National Statistics (UK) | 2019 | [27,28] |
| Portugal | TourMIS; Office for National Statistics (UK) | 2019 | [27,28] |
| Romania | National Institute of Statistics (Romania) Office for National Statistics (UK) | 2019 | [28,41] |
| | | | [28,42,43] |
| | | | [27,28] |
| Slovakia | Ministry of Transport and Construction of the Slovak Republic; Office for National Statistics (UK) | 2019 | [28,44] |
| Slovenia | TourMIS; Office for National Statistics (UK) | 2019 | [27,28] |
| Spain [d] | Office for National Statistics (UK) | 2019 | [28,45,46] |
| United Kingdom | Office for National Statistics (UK) | 2019 | [47] |

[a] For any UK related travel, data from the Office for National Statistics (UK) was the primary data source for parameterization
[b]The duration of stay in Denmark is based on the global number of arrivals and number of nights stayed. The number of arrivals to Denmark was determined by dividing the number of nights spent in any accommodation by the duration of stay.
[c]Average duration Norway assumed to be fixed for all countries and based on Jan 2021 travel
[d] The number of arrivals to the frontier for Spain were scaled by the percentage of travellers that stayed in paid accommodations. The duration of stay is based on the average duration of stay of any traveller to Spain.

**Table S2.** Parameters describing the epidemic profile of the 26 European countries as of November 21.

| Country | Population size | Cases per 100,000 [a] | Prevalence of non-isolated infections per 100,000 [b] | Daily incidence per 100,000 [c] | Recovered population | Level of vaccine-acquired immunity [d] |
|---|---|---|---|---|---|---|
| Austria | 8916185 | 4144 | 5178 | 296.0 | 21.3% | 46.2% |
| Belgium | 11419166 | 2765 | 3794 | 197.5 | 31.4% | 46.4% |
| Bulgaria | 6934625 | 2094 | 3670 | 149.6 | 72.5% | 5.8% |
| Cyprus | 1313477 | 523 | 681 | 37.3 | 14.1% | 36.2% |
| Czechia | 10643487 | 5141 | 6750 | 367.2 | 69.8% | 13.9% |
| Denmark | 5802733 | 1881 | 2357 | 134.4 | 11.7% | 61.7% |
| Estonia | 1312361 | 1248 | 2109 | 89.2 | 29.4% | 37.7% |
| Finland | 5534095 | 745 | 1050 | 53.2 | 13.5% | 59.1% |
| France | 66204315 | 391 | 528 | 28.0 | 29.6% | 47.1% |
| Germany | 84914056 | 2029 | 2572 | 145.0 | 16.5% | 49.5% |
| Greece | 10337172 | 1635 | 2216 | 116.8 | 15.7% | 49.6% |
| Hungary | 9674413 | 6117 | 8306 | 436.9 | 53.3% | 22.3% |
| Ireland | 4910357 | 2999 | 4027 | 214.2 | 19.3% | 55.3% |
| Italy | 60313170 | 571 | 725 | 40.8 | 23.3% | 51.4% |
| Lithuania | 2794223 | 2266 | 3919 | 161.8 | 66.9% | 17.9% |
| Luxembourg | 618550 | 969 | 1287 | 69.2 | 24.8% | 46.9% |
| Malta | 439221 | 410 | 516 | 29.3 | 15.8% | 66.1% |
| Netherlands | 17156788 | 2794 | 3584 | 199.6 | 25.0% | 47.8% |
| Norway | 5348847 | 1432 | 2211 | 102.3 | 24.4% | 51.0% |
| Poland | 38434445 | 3077 | 4040 | 219.8 | 50.0% | 22.9% |
| Portugal | 10651263 | 608 | 747 | 43.4 | 20.9% | 57.3% |
| Romania | 19237066 | 1108 | 2157 | 79.1 | 66.1% | 11.5% |
| Slovakia | 5437223 | 3398 | 4483 | 242.7 | 24.0% | 31.2% |
| Slovenia | 2074271 | 3864 | 5793 | 276.0 | 43.7% | 26.6% |
| Spain | 46021218 | 325 | 418 | 23.2 | 24.4% | 53.5% |

| | | | | | | |
|---|---|---|---|---|---|---|
| United Kingdom | 67220447 | 1318 | 1829 | 94.1 | 28.8% | 33.2% |

[a] Estimated cases per 100,000 is determined over a two-week period (November 8, 2021 to November 21, 2021), as specified by the European Union COVID-19 risk stratification
[b] Prevalence of non-isolated infections is a weighted calculation based on the proportion of symptomatic infections and asymptomatic infections, with symptomatic cases isolating on symptom onset. For symptomatic infections, case counts over the last six days are used to reflect the duration of the incubation period. For asymptomatic infections, case counts used over the last 26 days are used to reflect the duration of disease.
[c] The daily incidence per capita was computed as the average daily incidence per capita for the period of November 8, 2021 to November 21, 2021. The daily incidence per capita used in the calculation is per individual and not per 100,000
[e] The level of vaccine-acquired immunity is dependent on the age-demographics of the country, the proportion of the population partially and fully immunised, the age- and dose-specific vaccine effectiveness, and the vaccination coverage two weeks prior (e.g. November 7 for November 21).

**Table S3.** The percentage of infections that are attributed to the variants of concern Delta G/478K.V1 and Omicron B.1.1.529+BA [48 a]

| Country | Percentage of cases attributed to VOC Delta G/478K.V1 | Percentage of cases attributed to VOC Omicron B.1.1.529+BA |
|---|---|---|
| Austria | 21.5% | 1.7% |
| Belgium | 88.1% | 1.0% |
| Bulgaria | 100.0% | N/A |
| Cyprus | 0.0% | N/A |
| Czechia | 99.7% | 0.1% |
| Denmark | 100.0% | 0.0% |
| Estonia | 0.0% | N/A |
| Finland | 50.0% | 50.0% |
| France | 91.2% | 0.3% |
| Germany | 99.6% | 0.3% |
| Greece | 88.8% | 0.5% |
| Hungary | 0.0% | N/A |
| Republic of Ireland | 97.9% | 2.1% |
| Italy | 90.0% | 0.3% |
| Lithuania | 99.7% | N/A |
| Luxembourg | 99.9% | 0.1% |
| Malta | 100.0% | N/A |
| Netherlands | 44.9% | 1.5% |
| Norway | 86.4% | 5.4% |
| Poland | 100.0% | N/A |
| Portugal | 98.3% | 1.7% |
| Romania | 98.0% | 2.0% |
| Slovakia | 99.8% | N/A |

| | | |
|---|---|---|
| Slovenia | 99.5% | N/A |
| Spain | 94.6% | 0.9% |
| United Kingdom | 99.5% | 0.4% |

[a] These percentages are based on the proportion of cases in the past four weeks based on the data accessed December 10, 2021. N/A denotes that no measurements were available.

**Table S4.** Definitions of the terms in the inequality for daily new within-country imminent infection

| Imminent infections | Infection term | Age-class $j$ infection term | Abbreviation |
|---|---|---|---|
| Due to border closure between country A and country B | $N_T c_A R_{E_A}$ | $N_T\left(c_A \cdot \frac{w_{A,j}(1-\varphi_{A,j})}{(1-\varphi_A)}\right)\cdot R_{E_A,j}$ | $\lambda_{0,j}$ |
| From residents of country A who are not travelling and are susceptible to infection | $N_A c_A R_{E_A}$ | $N_A\left(c_A \cdot \frac{w_{A,j}(1-\varphi_{A,j})}{1-\varphi_A}\right)\cdot R_{E_A,j}$ | $\lambda_{1,j}$ |
| From previously susceptible residents of country B who are currently in country A | $n_B\, c_A\left(\frac{1-\varphi_B}{1-\varphi_A}\right)R_{E_A}$ | $n_B \cdot \left(c_A \cdot \frac{w_{B,j}(1-\varphi_{B,j})}{1-\varphi_A}\right)\cdot R_{E_A,j}$ | $\lambda_{2,j}$ |
| From infectious residents of country A who are travelling to country B that need to be removed | $n_{A,B}\rho_A R_V$ | $n_{A,B}\cdot w_{A,j}\cdot \rho_{A,j}\cdot R_{V,j}$ | $\lambda_{3,j}$ |
| From residents of country A who where infected abroad, have gone through quarantine and are returning to country A | $n_{A,B}\rho_B\left(\frac{1-\varphi_A}{1-\varphi_B}\right)R_{Q_A,V}$ | $n_{A,B}\cdot w_{A,j}\cdot \rho_{B,j}\left(\frac{1-\varphi_{A,j}}{1-\varphi_{B,j}}\right)\cdot R_{Q_A,j}$ | $\lambda_{4,j}$ |
| From infectious residents of country B who are travelling to country A and have gone through quarantine | $n_{B,A}\rho_B R_{Q_A,V}$ | $n_{B,A}\cdot w_{B,j}\cdot \rho_{B,j}\cdot R_{Q_A,j}$ | $\lambda_{5,j}$ |
| From infectious residents of country B who were infected abroad in country A and are returning to country B that need to be removed | $n_{B,A}\rho_A\left(\frac{1-\varphi_B}{1-\varphi_A}\right)R_V$ | $n_{B,A}\cdot w_{B,j}\cdot \rho_{A,j}\left(\frac{1-\varphi_{B,j}}{1-\varphi_{A,j}}\right)\cdot R_{V,j}$ | $\lambda_{6,j}$ |

**Table S5.** Age-specific parameters used in the travel quarantine analysis

| Parameter | Age class | | | | | | Reference |
|---|---|---|---|---|---|---|---|
| | 0–19 | 20–29 | 30–39 | 40–49 | 50–59 | ≥60[c] | |
| Percentage of infections asymptomatic | 81.91% | 77.59% | 77.59% | 69.46% | 69.46% | 58.48% | [49] |
| Vaccine effectiveness (First dose): Transmission [a] | 32% | 32% | 32% | 30% | 30% | 28.8% | [23] |
| Vaccine effectiveness (Second dose): Transmission [b] | 94% | 94% | 94% | 90% | 90% | 91.5% | [23] |

[a] Reduction in documented infection during the time from 0–27 days after first dose
[b] Reduction in documented infection from 7 days after second dose to the end of follow-up
[c] The age specific parameters for 60 and older were estimated based on the European demographics for 60-69, 70-79, and 80 and older.

**Table S6.** Parameter descriptions and values used to determine the minimum duration of travel quarantine

| Parameter | Definition | Value | Reference |
|---|---|---|---|
| $t_S$ | Incubation period | 5.72, 8.29, and 11.66 days | [6,11,50] |
| $t_L$ | Latent period | 2.9 days | [5] |
| $t_E$ | Duration of disease | $t_S + 20$ | [7–9] |
| $R$ | Reproduction number in the absence of self-isolation | 3 | [1] |
| $N$ | Population size (EU traffic-light analysis) | $19.4\times 10^6$ | The average population size of the 26 countries |
| $n_{AB}$ and $n_{BA}$ | Number of travellers each day (EU traffic-light analysis) | 1794 | Based the average proportion of the total population travelling for the different origin-destination country pairs |
| $r$ | Natural immunity (EU traffic-light analysis) | 0.32 | The average level of natural immunity among the 26 countries |
| $d_{AB}$ and $d_{BA}$ | Duration of stay to determine proportion abroad (EU traffic-light analysis) | 3.6 days | The average duration of stay among origin-destination country pairs |
| $\tau$ | Increase in transmission for Alpha 202012/01 GRY variant of concern | 0.82 | [51–53] |
| $\tau$ | Increase in transmission for Delta G/478K.V1 variant of concern | 1.912 | [54] |
| $C$ | Scaling constant for the RT-PCR diagnostic sensitivity curve (8.29 day incubation period) | 9.605 | Estimated |
| $z$ | Determines the shape of the RT-PCR diagnostic sensitivity curve (8.29 day incubation period) | 0.474 | Estimated |
| $K$ | Determines the timing of peak diagnostic sensitivity of the RT-PCR diagnostic sensitivity curve (8.29 | 2.202 | Calculated |

| | | | |
|---|---|---|---|
| | day incubation period) | | |
| $C$ | Scaling constant for the RT-PCR diagnostic sensitivity curve (5.72 day incubation period) | 9.023 | Estimated |
| $z$ | Determines the shape of the RT-PCR diagnostic sensitivity curve (5.72 day incubation period) | 0.746 | Estimated |
| $K$ | Determines the timing of peak diagnostic sensitivity of the RT-PCR diagnostic sensitivity curve (5.72 day incubation period) | 1.852 | Calculated |
| $C$ | Scaling constant for the RT-PCR diagnostic sensitivity curve (11.66 day incubation period) | 9.225 | Estimated |
| $z$ | Determines the shape of the RT-PCR diagnostic sensitivity curve (11.66 day incubation period) | 0.385 | Estimated |
| $K$ | Determines the timing of peak diagnostic sensitivity of the RT-PCR diagnostic sensitivity curve (11.66 day incubation period) | 2.580 | Calculated |
| $\beta_0$ | Logistic regression coefficient for the percent positive agreement curve for the rapid antigen test | 2.552 | Estimated |
| $\beta_1$ | Logistic regression coefficient for the percent positive agreement curve for the rapid antigen test | -0.379 | Estimated |

7 Xiao AT, Tong YX, Gao C, Zhu L, Zhang YJ, Zhang S. Dynamic Profile of RT-PCR Findings from 301 COVID-19 Patients in Wuhan, China: A Descriptive Study. SSRN Electronic Journal. DOI: 10.2139/ssrn.3548769.

8 CDC. Interim Guidance on Duration of Isolation and Precautions for Adults with COVID-19. 2021; published online March 16. https://www.cdc.gov/coronavirus/2019-ncov/hcp/duration-isolation.html (accessed April 13, 2021).

9 Cevik M, Tate M, Lloyd O, Maraolo AE, Schafers J, Ho A. SARS-CoV-2, SARS-CoV, and MERS-CoV viral load dynamics, duration of viral shedding, and infectiousness: a systematic review and meta-analysis. *Lancet Microbe* 2021; **2**: e13–22.

10 Qin J, You C, Lin Q, Hu T, Yu S, Zhou X-H. Estimation of incubation period distribution of COVID-19 using disease onset forward time: A novel cross-sectional and forward follow-up study. *Sci Adv* 2020; **6**: eabc1202.

11 McAloon C, Collins Á, Hunt K, *et al.* Incubation period of COVID-19: a rapid systematic review and meta-analysis of observational research. *BMJ Open* 2020; **10**: e039652.

12 Hellewell J, Russell TW, The SAFER Investigators and Field Study Team, *et al.* Estimating the effectiveness of routine asymptomatic PCR testing at different frequencies for the detection of SARS-CoV-2 infections. *BMC Medicine* 2021; **19**. DOI:10.1186/s12916-021-01982-x.

13 Ashcroft P, Lehtinen S, Angst DC, Low N, Bonhoeffer S. Quantifying the impact of quarantine duration on COVID-19 transmission. *Elife* 2021; **10**. DOI:10.7554/eLife.63704.

14 Young S, Taylor SN, Cammarata CL, *et al.* Clinical Evaluation of BD Veritor SARS-CoV-2 Point-of-Care Test Performance Compared to PCR-Based Testing and versus the Sofia 2 SARS Antigen Point-of-Care Test. *J Clin Microbiol* 2020; **59**. DOI:10.1128/JCM.02338-20.

15 [No title]. https://ec.europa.eu/eurostat/databrowser/product/page/AVIA_PAOCC__custom_1298176 (accessed Sept 20, 2021).

16 COVID-19 Vaccine Tracker. https://vaccinetracker.ecdc.europa.eu/public/extensions/COVID-19/vaccine-tracker.html#age-group-tab (accessed

Sept 14, 2021).

17 robert-koch-institut. COVID-19-Impfungen_in_Deutschland/Aktuell_Deutschland_Landkreise_COVID-19-Impfungen.csv at master · robert-koch-institut/COVID-19-Impfungen_in_Deutschland. https://github.com/robert-koch-institut/COVID-19-Impfungen_in_Deutschland (accessed Oct 26, 2021).

18 Figures on the COVID-19 vaccination programme. https://www.rivm.nl/en/covid-19-vaccination/figures-vaccination-programme (accessed Oct 26, 2021).

19 Hill RYA. Coronavirus (COVID-19) Infection Survey, antibody and vaccination data, UK. 2021; published online Sept 16. https://www.localhost/peoplepopulationandcommunity/healthandsocialcare/conditionsanddiseases/datasets/coronaviruscovid19antibodydatafortheuk (accessed Oct 26, 2021).

20 Population Pyramids of the World from 1950 to 2100. https://www.populationpyramid.net/ europe/2019/ (accessed April 20, 2021).

21 Prem K, van Zandvoort K, Klepac P, *et al.* Projecting contact matrices in 177 geographical regions: an update and comparison with empirical data for the COVID-19 era. DOI:10.1101/2020.07.22.20159772.

22 COVID-19 Vaccine Tracker. https://vaccinetracker.ecdc.europa.eu/public/extensions/COVID-19/vaccine-tracker.html#age-group-tab (accessed Sept 14, 2021).

23 Dagan N, Barda N, Kepten E, *et al.* BNT162b2 mRNA Covid-19 Vaccine in a Nationwide Mass Vaccination Setting. *N Engl J Med* 2021; published online Feb 24. DOI:10.1056/NEJMoa2101765.

24 COVID-19 Vaccine Tracker. https://vaccinetracker.ecdc.europa.eu/public/extensions/COVID-19/vaccine-tracker.html#age-group-tab (accessed Sept 14, 2021).

25 Population Pyramids of the World from 1950 to 2100. https://www.populationpyramid.net/ europe/2019/ (accessed April 20, 2021).

26 Hellewell J, Russell TW, The SAFER Investigators and Field Study Team, *et al.* Estimating the effectiveness of routine asymptomatic PCR testing at different frequencies for the detection of SARS-CoV-2 infections. *BMC Medicine* 2021; **19**,. DOI:10.1186/s12916-021-01982-x.

27 MODUL University Vienna. City Tourism Recovery Monitor (Total foreign and domestic). TourMIS. https://www.tourmis.info/index_e.html (accessed March 30, 2021).

28 Travel trends estimates: UK residents‘ visits abroad - Office for National Statistics. https://www.ons.gov.uk/peoplepopulationandcommunity/leisureandtourism/datasets/ukresidentsvisitsabroad (accessed April 15, 2021).

29 Резултат от справка. https://infostat.nsi.bg/infostat/pages/reports/result.jsf?x_2=1113 (accessed July 9, 2021).

30 StatBank Denmark: Table 'TURIST2'. https://www.statbank.dk/statbank5a/default.asp?w=1500 (accessed July 9, 2021).

31 StatBank Denmark: Table 'TURIST'. https://www.statbank.dk/statbank5a/default.asp?w=1500 (accessed July 9, 2021).

32 Cuignet L. Le 4 Pages: More than 89 million foreign tourists in France in 2018. ÉTUDES ÉCONOMIQUES, 2018 https://www.entreprises.gouv.fr/files/files/en-pratique/etudes-et-statistiques/4pages/2019-4pn88-eve-english-version.pdf (accessed April 21, 2021).

33 Greece in Figures - ELSTAT. Arrivals and nights spent in hotels, similar establishments and tourist campsites by Region and by country of residence. 2019. https://www.statistics.gr/en/greece-in-figures?p_p_id=com_liferay_portal_search_web_portlet_SearchPortlet_INSTANCE_3&p_p_lifecycle=0&p_p_state=maximized&p_p_mode=view&_com_liferay_portal_search_web_portlet_SearchPortlet_INSTANCE_3_mvcPath=%2Fview_content.jsp&_com_liferay_portal_search_web_portlet_SearchPortlet_INSTANCE_3_assetEntryId=17069628&_com_liferay_portal_search_web_portlet_SearchPortlet_INSTANCE_3_type=document (accessed April 15, 2021).

34 Greece in Figures - ELSTAT. Arrivals and nights spent in hotels, similar establishments and tourist campsites (Final Data). https://www.statistics.gr/en/greece-in-figures?p_p_id=com_liferay_portal_search_web_portlet_SearchPortlet_INSTANCE_3&p_p_lifecycle=0&p_p_state=maximized&p_p_mode=view&_com_liferay_portal_search_web_portlet_SearchPortlet_INSTANCE_3_mvcPath=%2Fview_content.jsp&_com_liferay_portal_search_web_portlet_SearchPortlet_INSTANCE_3_assetEntryId=17020463&_com_liferay_portal_search_web_portlet_SearchPortlet_INSTANCE_3_type=document (accessed April 15, 2021).

35 Key Tourism Facts 2019. Failte Ireland. 2021; published online March. https://www.failteireland.ie/FailteIreland/media/WebsiteStructure/Documents/3_Research_Insights/4_Visitor_Insights/KeyTourismFacts_2019.pdf?ext=.pdf (accessed April 21, 2021).

36 OECD. Istat Statistics. http://dati.istat.it/?lang=en (accessed April 21, 2021).

37 Trade and tourism. Statistic Portal: Grand Duchy of Luxembourg. https://statistiques.public.lu/stat/ReportFolders/ReportFolder.aspx?IF_Language=eng&MainTheme=4&FldrName=6&RFPath=19 (accessed July 9, 2021).

38 Inbound Tourism: December 2019. National Statistics Office - Malta, 2020 https://nso.gov.mt/en/News_Releases/Archived_News_Releases/Documents/2020/News2020_017.pdf (accessed July 9, 2021).

39 Norway S. 08401: Accommodation establishments total. Guest nights, by guests' country of residence (C) (closed series) 2005M01 - 2019M12. StatBank. https://www.ssb.no/en/statbank/table/08401/ (accessed April 21, 2021).

40 Statistics Norway. Accommodation: guest nights at commercial and non-commercial establishments. https://www.ssb.no/en/transport-og-reiseliv/reiseliv/statistikk/overnattingar (accessed April 21, 2021).

41 Romanian Tourism Statistical Abstract (Turismul Romaniei Breviar Statistic). National Institute of Statistics, 2020 https://insse.ro/cms/sites/default/files/field/publicatii/turismul_romaniei_2017-2019_breviar_statistic_0.pdf (accessed July 12, 2021).

42 Number of overnight stays in collective accommodation facilities. EMISS. https://www.fedstat.ru/indicator/31587 (accessed July 9, 2021).

43 Entry of foreign citizens to the Russian Federation. EMISS. https://www.fedstat.ru/indicator/38479 (accessed July 9, 2021).

44 Visitors in accommodation facilities of tourism in Slovakia in 2019. MINISTRY OF TRANSPORT AND CONSTRUCTION OF THE SLOVAK REPUBLIC.

https://www.mindop.sk/ministerstvo-1/cestovny-ruch-7/statistika/ubytovacia-statistika/rok-2019/navstevnici-v-ubytovacich-zariadeniach-cr-na-slovensku-za-rok-2019 (accessed July 9, 2021).

45 Estadística de Movimientos Turísticos en Fronteras (FRONTUR). Institute if National Statistics, 2020 https://www.ine.es/en/daco/daco42/frontur/frontur1219_en.pdf (accessed July 9, 2021).

46 Estadísticas e Informes. https://sedeaplicaciones.minetur.gob.es/Badase/BadasiUI/lstSeriesInformesPostBack.aspx (accessed July 9, 2021).

47 Travel trends estimates: overseas residents in the UK - Office for National Statistics. https://www.ons.gov.uk/peoplepopulationandcommunity/leisureandtourism/datasets/overseasresidentsvisitstotheuk (accessed April 15, 2021).

48 GISAID - hCov19 Variants. https://www.gisaid.org/hcov19-variants/ (accessed July 9, 2021).

49 Poletti P, Tirani M, Cereda D, *et al.* Association of Age With Likelihood of Developing Symptoms and Critical Disease Among Close Contacts Exposed to Patients With Confirmed SARS-CoV-2 Infection in Italy. *JAMA Netw Open* 2021; **4**: e211085–e211085.

50 Qin J, You C, Lin Q, Hu T, Yu S, Zhou X-H. Estimation of incubation period distribution of COVID-19 using disease onset forward time: A novel cross-sectional and forward follow-up study. *Sci Adv* 2020; **6**: eabc1202.

51 Davies NG, Abbott S, Barnard RC, *et al.* Estimated transmissibility and impact of SARS-CoV-2 lineage B.1.1.7 in England. *Science* 2021; **372**. DOI:10.1126/science.abg3055.

52 Volz E, Mishra S, Chand M, *et al.* Assessing transmissibility of SARS-CoV-2 lineage B.1.1.7 in England. *Nature* 2021; : 1–17.

53 Abdool Karim SS, de Oliveira T. New SARS-CoV-2 Variants - Clinical, Public Health, and Vaccine Implications. *N Engl J Med* 2021; published online March 24. DOI:10.1056/NEJMc2100362.

54 Mahase E. Delta variant: What is happening with transmission, hospital admissions, and restrictions? *BMJ* 2021; **373**. DOI:10.1136/bmj.n1513.
